# Bridging the gap: improving correspondence between low-field and high-field magnetic resonance images in young people

**DOI:** 10.1101/2024.01.05.24300892

**Authors:** Rebecca Cooper, Rebecca Hayes, Mary Corcoran, Kevin N Sheth, Thomas Campbell Arnold, Joel Stein, David C. Glahn, Maria Jalbrzikowski

## Abstract

**Background:** Portable low-field-strength magnetic resonance imaging (MRI) systems represent a promising alternative to traditional high-field-strength systems with the potential to make MR technology available at scale in low-resource settings. However, lower image quality and resolution may limit the research and clinical potential of these devices. We tested two super-resolution methods to enhance image quality in a low-field MR system and compared their correspondence with images acquired from a high-field system in a sample of young people.

**Methods:** T1- and T2-weighted structural MR images were obtained from a low-field (64mT) Hyperfine and high-field (3T) Siemens system in N = 70 individuals (mean age=20.39 years, range 9-26 years). We tested two super-resolution approaches to improve image correspondence between images acquired at high- and low-field: 1) processing via a convolutional neural network (‘SynthSR’), and 2) multi-orientation image averaging. We extracted brain region volumes, cortical thickness, and cortical surface area estimates. We used Pearson correlations to test the correspondence between these measures, and Steiger Z tests to compare the difference in correspondence between standard imaging and super-resolution approaches.

**Results:** Single pairs of T1- and T2-weighted images acquired at low field showed high correspondence to high-field-strength images for estimates of total intracranial volume, surface area cortical volume, subcortical volume, and total brain volume (*r* range=0.60-0.88). Correspondence was lower for cerebral white matter volume (*r=*0.32, *p=*.007, *q=*.009) and non-significant for mean cortical thickness (*r=*-0.05, *p=*.664, *q =*.664). Processing images with SynthSR yielded significant improvements in correspondence for total brain volume, white matter volume, total surface area, subcortical volume, cortical volume, and total intracranial volume (*r* range=0.85-0.97), with the exception of global mean cortical thickness (*r*=0.14). An alternative multi-orientation image averaging approach improved correspondence for cerebral white matter and total brain volume.

Processing with SynthSR also significantly improved correspondence across widespread regions for estimates of cortical volume, surface area and subcortical volume, as well as within isolated prefrontal and temporal regions for estimates of cortical thickness.

**Conclusions:** Applying super-resolution approaches to low-field imaging improves regional brain volume and surface area accuracy in young people. Finer-scale brain measurements, such as cortical thickness, remain challenging with the limited resolution of low-field systems.

## Introduction

Magnetic resonance imaging (MRI) has facilitated our current understanding of the processes underlying mental health and illness, and is routinely used in pediatric clinical and neuroimaging research (Figueiro Longo et al., 2022; Lee-Jayaram et al., 2020; Lerch et al., 2017). Ongoing advancements in neuroimaging technology and the development of state-of-the-art statistical techniques have helped to improve our understanding of the etiology of youth mental health disorders, and novel brain-based risk markers show promise in identifying and providing prognostic information for at-risk individuals (Andreou & Borgwardt, 2020; Ellis et al., 2020; Worthington et al., 2020). However, there are several challenges associated with conventional high-field MRI systems that limit the translation of these promising findings, prohibiting their widespread use and incorporation in community settings.

Low-field-strength MRI (LF-MRI) is a promising alternative that addresses several limitations inherent in high-field-strength systems. LF-MRI machines typically operate with magnetic fields below 0.3 Tesla (compared to high-field systems at 1.5-3T), and carry the advantages of substantially lower installation and maintenance costs, reduced power consumption, smaller space requirements, and do not require cryogenic cooling (Arnold et al., 2023; Kimberly et al., 2023). Several portable systems have been developed, and can be installed in settings with limited or unreliable power supply with minimal operator expertise. The successful deployment of LF-MRI within intensive care units (Mazurek et al., 2021, 2023; Rusche et al., 2022; Sheth et al., 2020; Yuen et al., 2022), vehicles (Deoni, Medeiros, et al., 2022), consulting offices (Guallart-Naval et al., 2022) and in low-resource settings (Chetcuti et al., 2022; Tu et al., 2023) demonstrates the increased accessibility offered by low-field technology. This improved accessibility carries the potential to reduce long-standing disparities in access to diagnostic imaging within the United States (Marin et al., 2021; Schrager et al., 2019). In addition, the lower-intensity acoustic noise and open scanner designs in LF systems provide advantages for pediatric populations, improving scanning success rates and reducing the need for child sedation (Raschle et al., 2012; Rupprecht et al., 2000). In a sample of 42 healthy children aged 6 weeks to 16 years of age, superior completion rates were achieved in a low-field (64mT) LF system compared to conventional high-field (3T) MRI system (89% compared to 75%; Deoni et al., 2021). In addition, global estimates of cortical volume showed strong correspondence between low-field- and high-field acquired images, with low-field images successfully recapitulating global gray-matter age-associations (Deoni et al., 2021). However, components of cortical volume (e.g., cortical thickness, surface area), as well as cerebral white matter and subcortical volume, show diverse trajectories of growth across development (Herting et al., 2018; Lebel & Deoni, 2018; Tamnes et al., 2010; Wierenga et al., 2014), are believed to reflect distinct biological underpinnings (Grasby et al., 2020; Pontious et al., 2008; Rakic, 1988), and have differing genetic influences (Grasby et al., 2020; Winkler et al., 2010). For LF-MRI to be feasibly used at scale in young people, we need to test the ability of this technology to accurately estimate diverse components of global and regional brain structure.

One of the major drawbacks of LF systems is a low signal-to-noise ratio (SNR), resulting in poorer image resolution and quality (Arnold et al., 2023; Kimberly et al., 2023). However, recent developments in ‘super-resolution’ approaches, defined as methods that reconstruct high-resolution images from a series of low-resolution images, may help to address these shortcomings. Multi-orientation image averaging, which involves reconstruction of several low-resolution scans taken in orthogonal slice directions (i.e. axial, sagittal and coronal), has been found to significantly improve signal-to-noise ratio within neonatal samples (Askin Incebacak et al., 2022; Sui et al., 2021). Alternative super-resolution approaches that use state-of-the-art machine-learning techniques, such as convolutional neural networks (CNNs), also improve image resolution within LF systems (Iglesias et al., 2023). Initial work in a sample of adults (N = 11, M = 49.5 ± 14.1 years, seven males) demonstrated promising results with this approach, resulting in high correspondence between low-field (64mT) and high-field (1.5 or 3T) acquisitions across the cerebrum (Iglesias et al., 2023). While these studies show promise, we must ensure that such pipelines are developmentally appropriate for the acquisition and processing of low-field images among young people. It is unclear whether current super-resolution approaches, such as multi-orientation image averaging and machine-learning-based methods, are appropriate and effective for young populations, whether the effectiveness of these pipelines are moderated by additional factors (e.g., age, motion) or whether additional processing steps are necessary.

To test our ability to use data from low-field MRI scans in young people, we collected structural MRI data from low-field 64 mT and 3T MRI scanners from a community sample of young people (final N = 70, 9-26 years). We processed all scans through a standard structural neuroimaging pipeline (i.e., Freesurfer), and extracted measures of cortical and subcortical volume, cortical thickness, and surface area. We then used these extracted measures to conduct and compare correlational analyses of two super-resolution processing strategies. First, we examined correspondence between brain measures extracted from low-field scans to brain measures extracted from 3T scans. Second, we tested whether synthesizing super-resolution MP-RAGE images from the low-field scans via a CNN approach improved the correspondence between low-field and high-field images. Third, because we collected several pairs of low-field T1- and T2-weighted scans, we examined how multi-orientation image averaging improved correspondence with measures derived from high-field scans. We tested how implementing a combination of both multi-orientation and CNN-processing approaches influenced these relationships. Finally, we examined how other factors, i.e., age and motion (Reuter et al., 2015; Satterthwaite et al., 2012; Yao et al., 2017), are related to our ability to capture high-field quality measurements with low-field scans.

## Materials and Methods

### Participants

In the current study, we recruited a community sample of 77 young people (9-26 years, N <18 years = 18) from the Boston metro area. Inclusion/exclusion of data to obtain the final sample (N = 70, mean (SD) age = 20.39 (4.7) years) is detailed in Supplemental Figure 1. Demographic information for the final sample is reported in Table 1. Exclusion criteria were a history of a brain infection, presence of a neurodegenerative disorder (e.g., Parkinson’s disorder), presence of a neurodevelopmental disorder that might interfere with completion of study procedures, endorsement of a major mental disorder other than attention deficit-hyperactivity disorder (ADHD) or a past episode of depression, or any MRI contraindications. Given the increased prevalence of ADHD in youth (Fairman et al., 2020; Safer, 2018) and our desire to produce generalizable results, we did not exclude those with a diagnosis of ADHD in this sample. Participants completed the DSM Cross-Cutting Symptom Measure for Youth (Narrow et al., 2013) and the Beck Depression Inventory (Beck et al., 1988) to assess sub-clinical symptoms of psychopathology. All participants provided written consent (if ≥18 years of age) or assent with written parental consent. Procedures were approved by the Institutional Review Board of Boston Children’s Hospital.

**Table 1.** Demographic features of the sample. BDI, Beck Depression Inventory. CCSM, DSM self-rated Cross-Cutting Symptom Measure for Youth.

|  |  | **F** | **M** | **TOTAL** |
| --- | --- | --- | --- | --- |
| Total N |  | 37 | 33 | 70 |
| Mean Age<br>(SD) |  | 19.7<br>(4.9) | 21.17<br>(4.5) | 20.39<br>(4.7) |
| Age Range |  | 9-26.7 | 9.8-26.9 | 9-26.9 |
| Mean CCSM Total Score<br>(SD) |  | 9.11<br>(7.3) | 8.82<br>(7.6) | 8.97<br>(7.4) |
| Mean BDI Total Score<br>(SD) |  | 5.15<br>(7.2) | 4.85<br>(5.4) | 5.04<br>(6.5) |
| Race N<br>(%) | American<br>Indian/Alaskan<br>Native | 1 | 0 | 1<br>(1.4%) |
|  | Asian | 11 | 11 | 22<br>(31.4%) |
|  | Black | 3 | 1 | 4<br>(5.7%) |
|  | White | 22 | 20 | 42<br>(60%) |
|  | Native<br>Hawaiian/Pacific<br>Islander | 0 | 0 | 0<br>(0%) |
|  | Interracial | 0 | 1 | 1<br>(1.4%) |
| Ethnicity N<br>(%) | Hispanic/Latino | 3 | 4 | 7<br>(10%) |
|  | Not<br>Hispanic/Latino | 34 | 29 | 63<br>(90%) |

### MRI Acquisition

We collected T1- and T2-weighted brain MRI scans for each participant using a Siemens Magnetom Prisma 3-Tesla scanner in a dedicated research MRI suite at Boston Children’s Hospital Brookline Place (total scan duration 12.61 minutes, scan resolution 0.8×0.8×0.8mm). We also used a Hyperfine Swoop 64 mT scanner (total scan duration 52.25 minutes, scan resolution 1.6×1.6×5mm) to collect two pairs of low-field T1- and T2-weighted scans using 2-dimensional techniques in sagittal, axial, and coronal orientations (see Figure 1). Scan parameters are detailed in Supplemental Table 1. An experienced radiologist reviewed all 3T scans and reported no incidental findings in this sample. We used MRIQC (Esteban et al., 2017) to verify scan quality, which provides a rating (1 [unusable] – 4 [excellent]) for each scan. We excluded all 3T MRI scans with a rating of 1 or 2 (N = 1).

**Figure 1.**
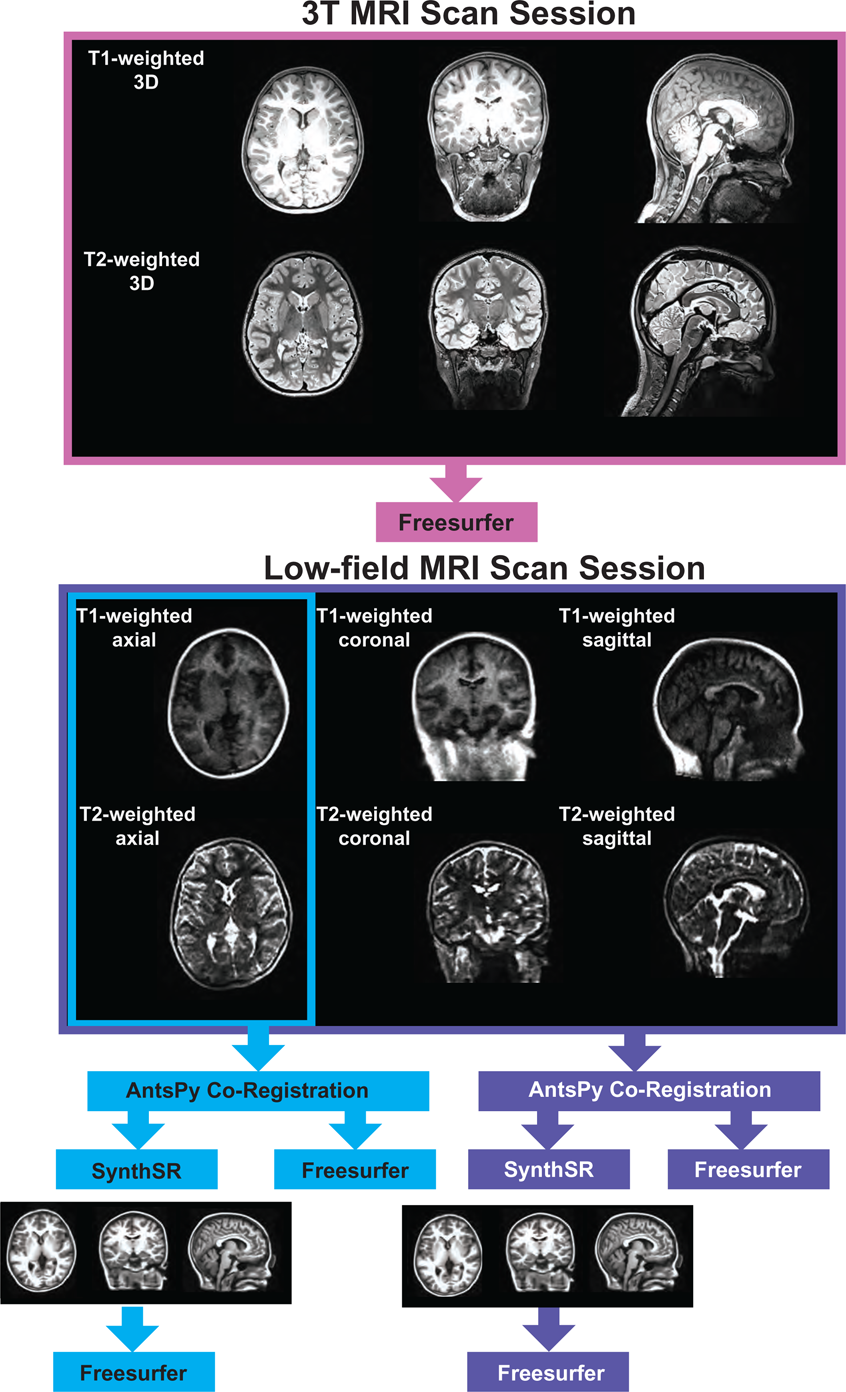
Image acquisition and processing pipeline.

### MRI Processing

We tested two super-resolution approaches to process low-field data. For the single-acquisition approach, we used the ANTsPy Python module (v 0.3.9) (Avants et al., 2008; Klein et al., 2009) to resample and coregister the axial-orientation T1- and T2-weighted low-field scans for each participant. For the multi-orientation approach, all six T1- and T2-weighted low-field MRI pairs were resampled to a 1.5mm3 voxel size, then co-registered and averaged to reconstruct a single, composite higher-resolution image with ANTsPy. We then processed the low-field images via the Hyperfine-specific version of SynthSR (v. 1.0) (Iglesias et al., 2021).

SynthSR is a convolutional neural network that predicts a 1mm isotropic three-dimensional MP-RAGE image given one or more low-resolution inputs. We then processed all low-field and 3T MR images using FreeSurfer (v. 7.3.2) (Fischl, 2012). FreeSurfer is an open-source automated segmentation software package for neuroimaging. In short, the following steps are part of the FreeSurfer processing stream: transformation of images to standard Talairach space, intensity normalization, removal of non-brain tissue, segmentation of white matter and subcortical structures, and final segmentation of cortical surfaces. Once the data was processed, we used the Desikan atlas (Desikan et al., 2006) to extract regional measures of cortical surface area, cortical thickness, and cortical volume. The Desikan atlas includes 68 regions for each measure, resulting in a total of 204 cortical measures. We obtained subcortical volume measures using FreeSurfer’s subcortical atlas (N = 38 regions). We also extracted global measures of subcortical gray matter volume, total surface area, mean cortical thickness, cerebral white matter volume, total brain volume, cortical volume, and estimated total intracranial volume from each scan. This resulted in five sets of MR scans for each participant: 1) a single-acquisition standard low-field scan; 2) a single-acquisition, low-field scan processed through SynthSR; 3) a multi-orientation standard low-field scan; 4) a multi-orientation, SynthSR-processed low-field scan; and 5) a standard 3T scan.

### Statistical Analyses

To test the association between the participants’ low-field and high-field scans we calculated Pearson correlations for each global and regional neuroimaging measure. We then used Steiger Z-tests to compare the difference in correspondence between the standard single-acquisition approach and each super-resolution method. First, we tested whether image processing via SynthSR statistically improved the fidelity of the low-field scans to their corresponding high-field images. Second, we compared correlation strengths between standard single-acquisition (one T1/T2 pair) and multi-orientation (six T1/T2 pairs) scan series. Third, we compared the standard single-acquisition approach to multi-orientation (six T1/T2 pairs) images processed with SynthSR. Fourth, we compared single-acquisition scans processed with SynthSR to multi-orientation scans processed with SynthSR. We used two additional methods to further evaluate the correspondence between low-field and 3T scans. First, we calculated the correspondence between low-field and 3T scans using two-way mixed intraclass correlation coefficients. Consistent with previous work (Lévy et al., 2018; Murata et al., 2021), we defined correspondence for both Pearson correlations and ICC values as follows: poor, r < 0.4; fair, 0.4 ≤ r < 0.6; moderate, 0.6 ≤ r < 0.75; high, r ≥ 0.75. Additionally, we used Bland-Altman plots to assess the agreement between low-field scans with the 3T estimates for each brain measure. This involved plotting the difference between the 3T and low-field estimates against the mean of the two estimates. For adequate agreement, Bland and Altman recommend that 95% of the data points should lie within ±1.96 standard deviations of the mean difference (Bland & Altman, 1999). Within each set of analyses, we used false discovery rate to correct for multiple comparisons separately in our global and regional analyses (Benjamini & Hochberg, 1995).

In secondary analyses, we examined whether potential confounding factors (participant age and motion artifacts) were associated with individual-level agreement between low- and high-field brain measures. We first calculated the absolute difference between low- and high-field estimates of each brain measure for each individual. We then converted this difference to a Z-score for each individual, with a positive Z-score indicating above average difference between high and low-field estimates (i.e., larger discrepancy between low-field and 3T measures) and a negative Z-score indicating below average difference between high- and low-field estimates (i.e., smaller discrepancy between low-field and 3T measures). We then examined the relationship between this Z-score and participant age or motion, measured as framewise displacement. We used Steiger Z-tests to examine whether these relationships significantly differed across the super-resolution approaches examined in the main analysis. Within each set of analyses, we used false discovery rate to correct for multiple comparisons separately in our global and regional analyses (Benjamini & Hochberg, 1995).

## Results

### Correspondence between standard (single-orientation) low-field- and high-field-acquired images

For global brain measures, single (axial) acquisitions of T1/T2 low-field pairs showed high correspondence with high-field images for measures of total intracranial volume (Pearson’s *r =* 0.82, *p =* 5.58e-18, *q =* 1.82e-17) and total surface area (*r =* 0.76, *p =* 4.01e-14, *q =* 1.04e-13), and moderate correspondence for cortical volume (*r =* 0.67, *p =* 3.24e-10, *q =* 7.21e-10), subcortical gray matter volume (*r =* 0.66, *p =* 3.83e-10, *q =* 8.16e-10), and total brain volume (*r =* 0.60, *p =* 4.57e-08, *q =* 9.33e-08; see Table 2, Figure 2, Figure 3A).

**Table 2.** Pearson correlation coefficients for the correspondence between low-field (64mT) and high-field (3T) MR images and comparisons across super-resolution approaches. Top row: Left column shows Pearson correlations between standard single low-field (64mT) and high-field (3T) images for each brain measure. Middle column shows Pearson correlations between SynthSR-processed low-field and high-field estimates. Right column shows results of Steiger Z-tests, assessing whether the correlations for standard and SynthSR-processed scans are statistically different. Middle row: Left column shows Pearson correlations between standard single low-field (64mT) and high-field (3T) images for each brain measure. Middle column shows Pearson correlations between standard multi-orientation low-field and high-field estimates. Right column shows results of Steiger Z-tests, assessing whether the correlations for single and multi-orientation approaches are statistically different. Bottom row: Left column shows Pearson correlations between standard single low-field (64mT) and high-field (3T) images for each brain measure. Middle column shows Pearson correlations between SynthSR-processed multi-orientation low-field and high-field estimates. Right column shows results of Steiger Z-tests, assessing whether the correlations for standard and SynthSR-processed multi-orientation approaches are statistically different. Values in bold survive correction for multiple comparisons.

| Brain measure | Standard Axial 64mT<br>Correlations with 3T |  |  | SynthSR-Processed Axial<br>64mT Correlations with<br>3T |  |  | Steiger's |  |  |
| --- | --- | --- | --- | --- | --- | --- | --- | --- | --- |
|  | *r* | *p* | *q* | *r* | *p* | *q* | *Z* | *p* | *q* |
| Total Surface Area | 0.76 | 4.01e-14 | 1.01e-13 | 0.90 | 9.01e-26 | 5.16e-25 | 3.73 | 1.94e-04 | 3.22e-04 |
| Mean Cortical Thickness | -0.05 | 0.664 | 0.697 | 0.14 | 0.241 | 0.286 | 1.43 | 0.153 | 0.186 |
| Estimated Intracranial Volume | 0.82 | 5.58e-18 | 1.67e-17 | 0.85 | 1.74e-20 | 6.10e-20 | 1.45 | 0.148 | 0.183 |
| Subcortical Gray Matter Volume | 0.66 | 3.83e-10 | 8.32e-10 | 0.88 | 4.89e-24 | 2.57e-23 | 4.57 | 4.84e-06 | 8.97e-06 |
| Cortical Volume | 0.67 | 3.24e-10 | 7.28e-10 | 0.86 | 1.94e-21 | 7.17e-21 | 4.18 | 2.90e-05 | 5.08e-05 |
| Cerebral White Matter Volume | 0.32 | 0.007 | 0.010 | 0.92 | 1.23e-28 | 7.74e-28 | 8.75 | 2.18e-18 | 6.88e-18 |
| Total Brain Volume | 0.60 | 4.57e-08 | 9.60e-08 | 0.97 | 1.15e-43 | 1.81e-42 | 9.80 | 1.15e-22 | 5.59e-22 |
| Brain measure | Standard Axial 64mT<br>Correlations with 3T |  |  | Standard Multi-Orientation 64mT<br>Correlations with 3T |  |  | Steiger's |  |  |
|  | *r* | *p* | *q* | *r* | *p* | *q* | *Z* | *p* | *q* |
| Total Surface Area | 0.76 | 4.01e-14 | 1.01e-13 | 0.58 | 1.07e-07 | 2.18e-07 | -2.52 | 0.012 | 0.017 |
| Mean Cortical Thickness | -0.05 | 0.664 | 0.697 | -0.25 | 0.038 | 0.051 | -1.54 | 0.123 | 0.155 |
| Estimated Intracranial Volume | 0.82 | 5.58e-18 | 1.67e-17 | 0.78 | 1.29e-15 | 3.39e-15 | -2.36 | 0.018 | 0.026 |
| Subcortical Gray Matter Volume | 0.66 | 3.83e-10 | 8.32e-10 | 0.71 | 5.86e-12 | 1.42e-11 | 0.94 | 0.349 | 0.393 |
| Cortical Volume | 0.67 | 3.24e-10 | 7.28e-10 | 0.54 | 1.40e-06 | 2.68e-06 | -1.76 | 0.079 | 0.101 |
| Cerebral White Matter Volume | 0.32 | 0.007 | 0.010 | 0.41 | 3.63e-04 | 5.73e-04 | 3.57 | 3.64e-04 | 5.73e-04 |
| Total Brain Volume | 0.60 | 4.57e-08 | 9.60e-08 | 0.69 | 3.30e-11 | 7.71e-11 | 3.49 | 4.84e-04 | 7.44e-04 |
| Brain measure | Standard Axial 64mT<br>Correlations with 3T |  |  | SynthSR-Processed Multi-Orientation 64mT<br>Correlations with 3T |  |  | Steiger's |  |  |
|  | *r* | *p* | *q* | *r* | *p* | *q* | *Z* | *p* | *q* |
| Total Surface Area | 0.76 | 4.01e-14 | 1.01e-13 | 0.87 | 1.42e-22 | 6.38e-22 | 2.85 | 0.004 | 0.007 |
| Mean Cortical Thickness | -0.05 | 0.664 | 0.697 | 0.12 | 0.317 | 0.363 | 1.09 | 0.275 | 0.321 |
| Estimated Intracranial Volume | 0.82 | 5.58e-18 | 1.67e-17 | 0.84 | 1.52e-19 | 5.05e-19 | 0.83 | 0.409 | 0.437 |
| Subcortical Gray<br>Matter Volume | 0.66 | 3.83e-10 | 8.32e-10 | 0.87 | 3.47e-22 | 1.46e-21 | 4.05 | 5.12e-05 | 8.71e-05 |
| Cortical Volume | 0.67 | 3.24e-10 | 7.28e-10 | 0.86 | 5.50e-22 | 2.17e-21 | 4.34 | 1.43e-05 | 2.57e-05 |
| Cerebral White Matter<br>Volume | 0.32 | 0.007 | 0.010 | 0.92 | 1.20e-28 | 7.74e-28 | 8.46 | 2.69e-17 | 7.69e-17 |
| Total Brain Volume | 0.60 | 4.57e-08 | 9.60e-08 | 0.96 | 3.64e-38 | 3.82e-37 | 8.38 | 5.52e-17 | 1.51e-16 |

**Figure 2.**
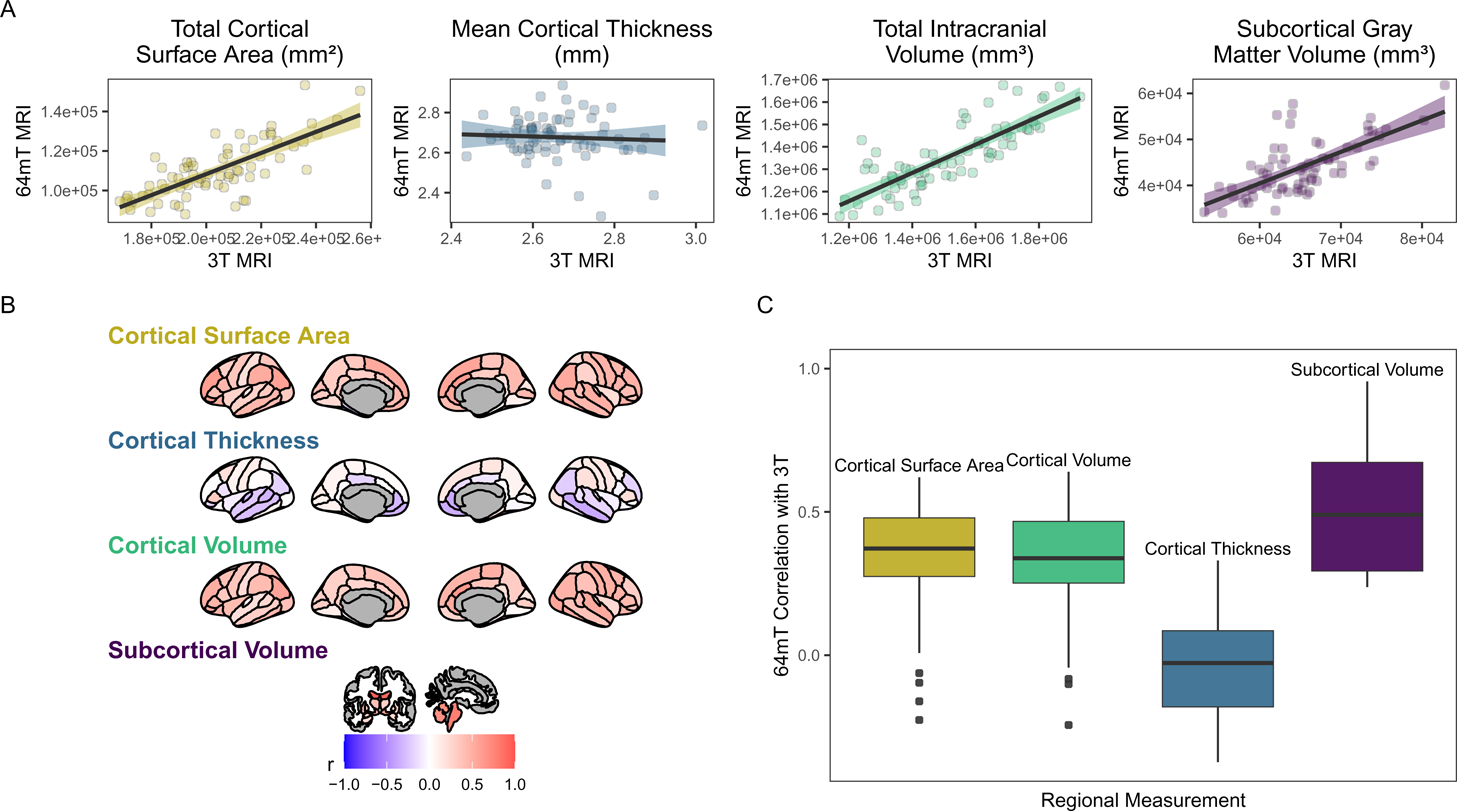
Correspondence between standard low-field (64mT) and high-field (3T) MR images. A. Scatterplots of relationships between low-field and high-field images for global brain measures. Data points represent global estimates for each individual. Shading denotes 95% confidence intervals. B. Pearson correlations for relationships between low-field and high-field images at the regional level. Shading represents mean correlation within each region across all individuals. C. Distribution of correlation strengths for relationships between standard low-field and high-field images across each brain measure. Data points represent mean correlation across each region of interest.

**Figure 3.**
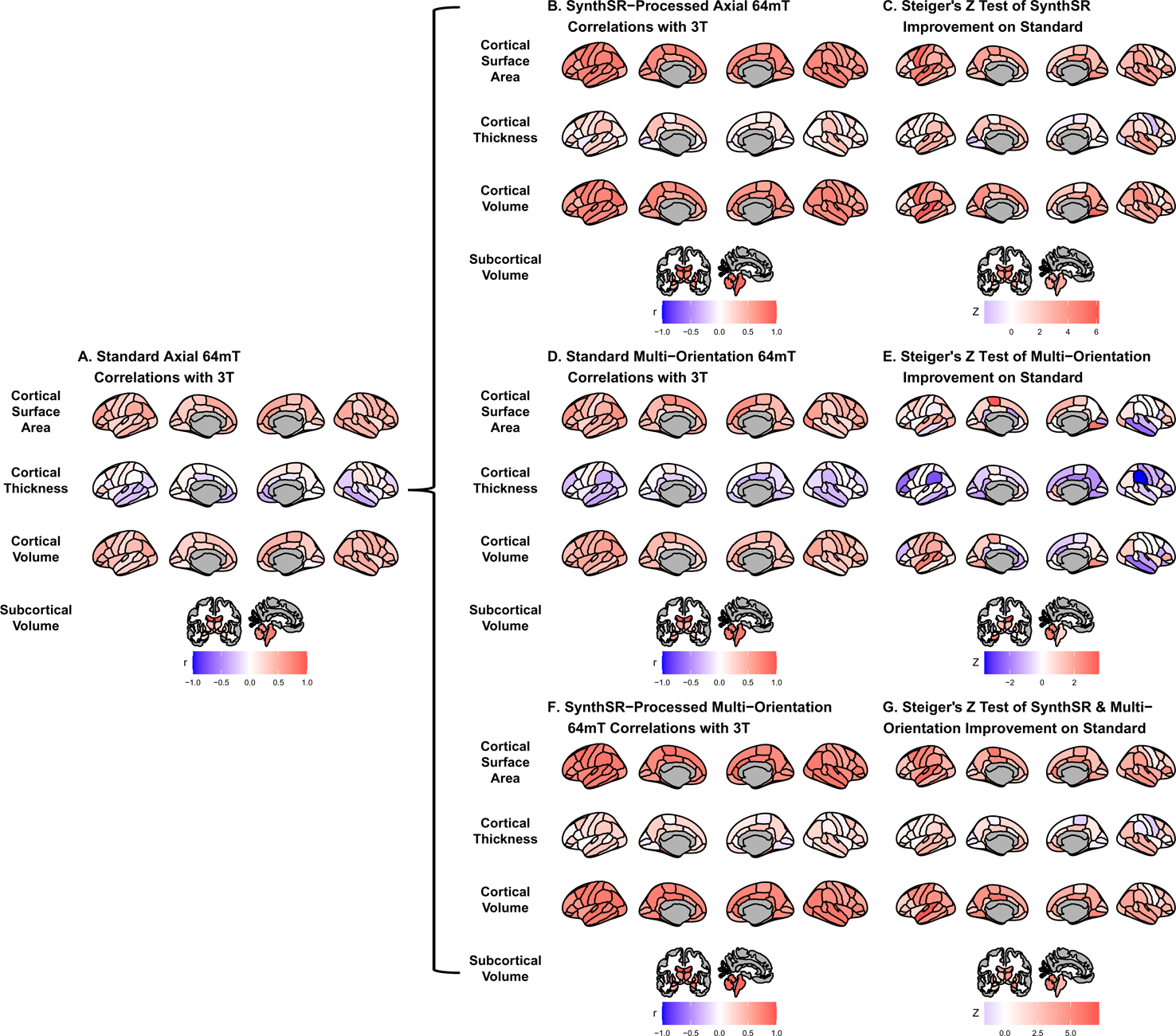
Comparison of super-resolution approaches in improving correspondence between low-field and high-field MR images. A. Correlation of standard single-acquisition low-field images with high-field images for surface area, cortical volume, cortical thickness and subcortical volume. B. Correlation of SynthSR-processed, single-acquisition low-field images with high-field images. C. Steiger Z-test values for change in correspondence between standard and SynthSR-processed data. D Correlation of multi-orientation standard low-field images with high-field images. E. Steiger Z-test values for change in correspondence between single-acquisition and multi-orientation approaches. F. Correlation of SynthSR-processed, multi-orientation low-field images with high-field images. G. Steiger Z-test values for change in correspondence between single-acquisition standard low-field images and multi-orientation, SynthSR-processed low-field images.

Correspondence was lower for cerebral white matter volume (*r =* 0.32, *p =* .007, *q =* .009) and non-significant for mean cortical thickness (*r =* -0.05, *p =* .664, *q =* .664). Similarly, intra-class correlation coefficients (ICC) for each brain measure showed high correspondence for total intracranial volume (ICC = 0.79, p = 1.11e-16, q = 3.04e-16), moderate correspondence for total surface area (ICC = 0.71, p = 1.65e-12, q = 4.15e-12) and cortical volume (ICC = 0.61, p = 1.08e-08, q = 2.34e-08), fair correspondence for subcortical gray matter volume (ICC = 0.66, p = 1.46e-10, q = 3.40e-10) and total brain volume (ICC = 0.56, p = 1.43e-07, q = 2.90e-07), poor correspondence for cerebral white matter volume (ICC = 0.24, p = .024, q = .036) and non-significant for mean cortical thickness (ICC = -0.05, p = .668, q = .701; see Table 3). When we examined the individual data distributions (Figure 4), we found that standard single-acquisition low-field images typically underestimated high-field values for each brain measure. Mean differences (low-field subtracting 3T) for surface area were -45.8% (standard deviation (SD) 4.4), cortical thickness +0.85% (5.5), cortical volume - 46.4% (5.3), subcortical volume -32.5% (7.0), cerebral white matter volume -33.8% (29.4), total brain volume - 37.0% (11.4) and intracranial volume -10.1% (6.83). Bland-Altman analyses for agreement between low- and high-field scans (see Supplemental Figure 2) showed positive bias for total surface area (bias 9.35e04mm^2^, 95% limits of agreement 6.78e04-11.93e04), total brain volume (4.44e05mm^3^ [1.62e5-7.25e5]), cortical volume (2.37e5mm^3^ [1.59e5-3.22e5]), intracranial volume (1.57e05mm^3^ [-0.55e5-3.70e5]), subcortical volume (2.12e04mm^3^ [1.16e4-3.08e4]) and cerebral white matter volume (1.53e05mm^3^ [-1.11e5-4.18e5]), indicating the low-field acquisition underestimated high-field data. Bias for cortical thickness was not significantly different from zero (−0.02mm [-0.31-0.28]), indicating that, on average, low-field estimates neither over- or under-estimated the high-field data.

**Table 3.**
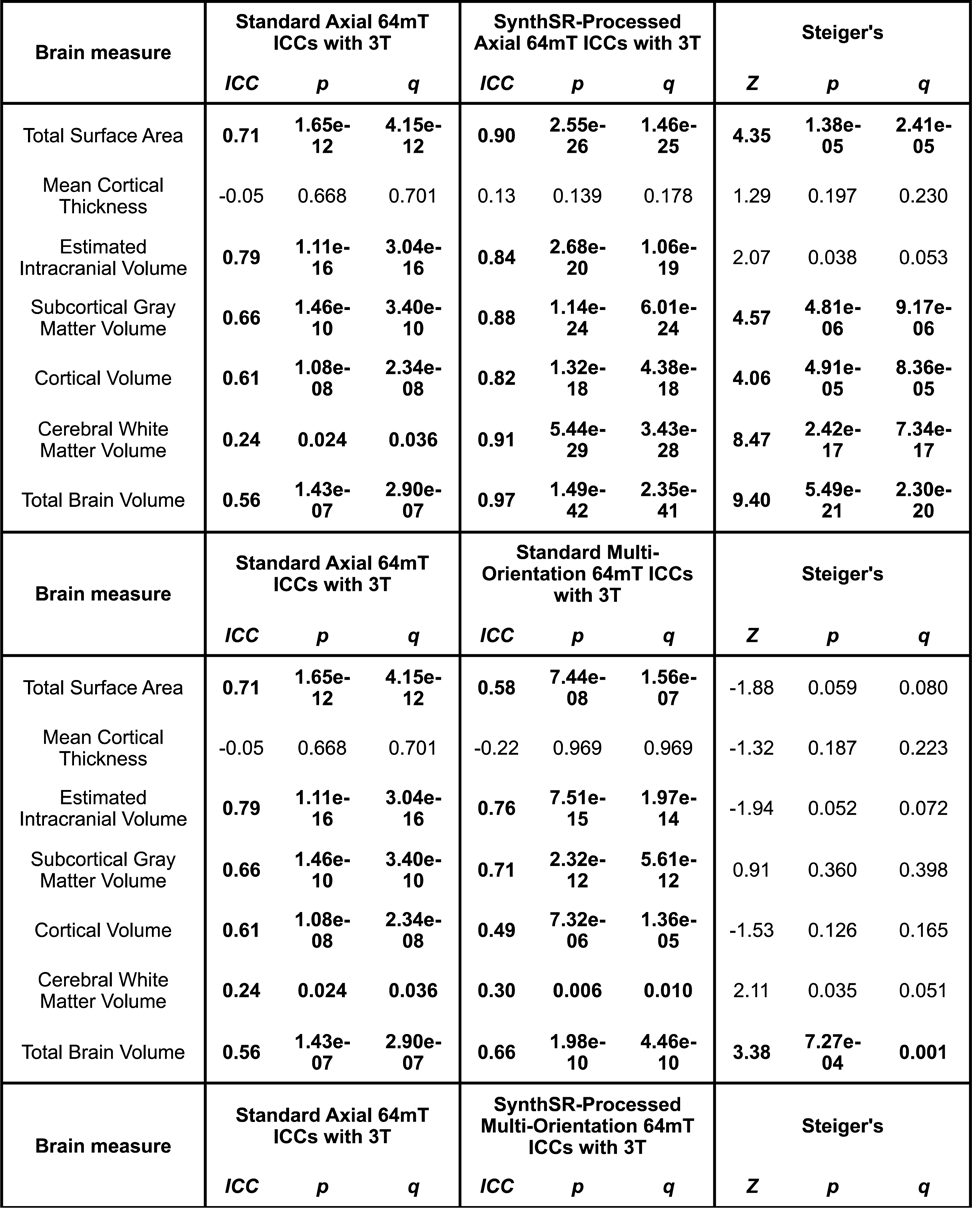
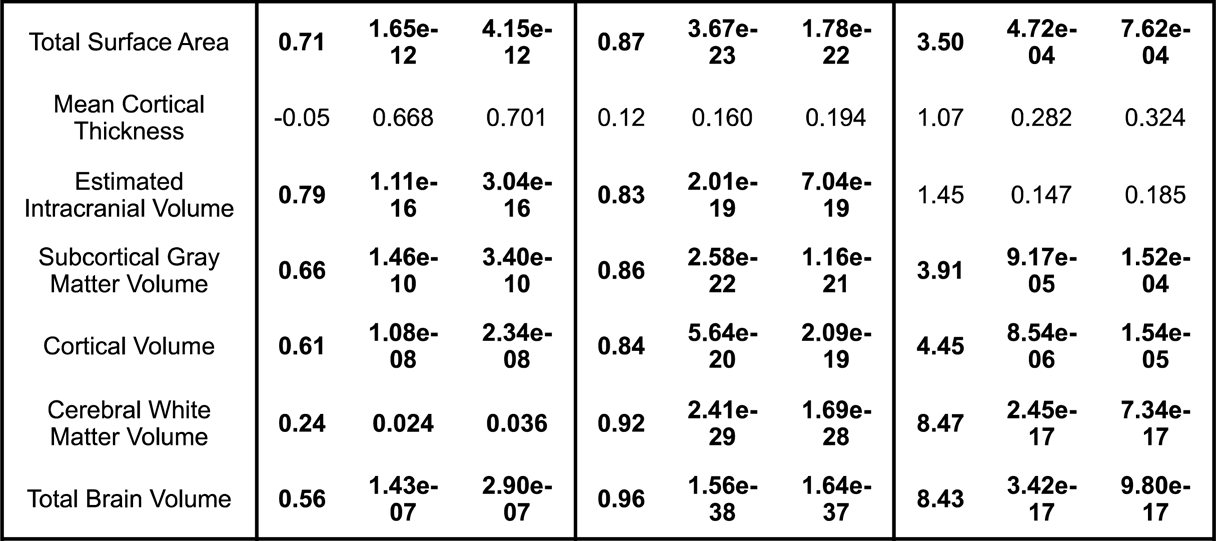
Intraclass correlation coefficients (ICCs) for the correspondence between low-field (64mT) and high-field (3T) MR images and comparisons across super-resolution approaches. Top row: Left column shows ICC values between standard single low-field (64mT) and high-field (3T) images for each brain measure. Middle column shows ICCs between SynthSR-processed low-field and high-field estimates. Right column shows results of Steiger Z-tests, assessing whether the ICCs for standard and SynthSR-processed scans are statistically different. Middle row: Left column shows ICCs between standard single low-field (64mT) and high-field (3T) images for each brain measure. Middle column shows ICCs between standard multi-orientation low-field and high-field estimates. Right column shows results of Steiger Z-tests, assessing whether the ICCs for single and multi-orientation approaches are statistically different. Bottom row: Left column shows ICCs between standard single low-field (64mT) and high-field (3T) images for each brain measure. Middle column shows ICCs between SynthSR-processed multi-orientation low-field and high-field estimates. Right column shows results of Steiger Z-tests, assessing whether the ICCs for standard and SynthSR-processed multi-orientation approaches are statistically different. Values in bold survive correction for multiple comparisons.

| Brain measure | Standard Axial 64mT ICCs with 3T |  |  | SynthSR-Processed Axial 64mT ICCs with 3T |  |  | Steiger's |  |  |
| --- | --- | --- | --- | --- | --- | --- | --- | --- | --- |
|  | ICC | p | q | ICC | p | q | Z | p | q |
| Total Surface Area | 0.71 | 1.65e-12 | 4.15e-12 | 0.90 | 2.55e-26 | 1.46e-25 | 4.35 | 1.38e-05 | 2.41e-05 |
| Mean Cortical Thickness | -0.05 | 0.668 | 0.701 | 0.13 | 0.139 | 0.178 | 1.29 | 0.197 | 0.230 |
| Estimated Intracranial Volume | 0.79 | 1.11e-16 | 3.04e-16 | 0.84 | 2.68e-20 | 1.06e-19 | 2.07 | 0.038 | 0.053 |
| Subcortical Gray Matter Volume | 0.66 | 1.46e-10 | 3.40e-10 | 0.88 | 1.14e-24 | 6.01e-24 | 4.57 | 4.81e-06 | 9.17e-06 |
| Cortical Volume | 0.61 | 1.08e-08 | 2.34e-08 | 0.82 | 1.32e-18 | 4.38e-18 | 4.06 | 4.91e-05 | 8.36e-05 |
| Cerebral White Matter Volume | 0.24 | 0.024 | 0.036 | 0.91 | 5.44e-29 | 3.43e-28 | 8.47 | 2.42e-17 | 7.34e-17 |
| Total Brain Volume | 0.56 | 1.43e-07 | 2.90e-07 | 0.97 | 1.49e-42 | 2.35e-41 | 9.40 | 5.49e-21 | 2.30e-20 |
| Brain measure | Standard Axial 64mT ICCs with 3T |  |  | Standard Multi-Orientation 64mT ICCs with 3T |  |  | Steiger's |  |  |
|  | ICC | p | q | ICC | p | q | Z | p | q |
| Total Surface Area | 0.71 | 1.65e-12 | 4.15e-12 | 0.58 | 7.44e-08 | 1.56e-07 | -1.88 | 0.059 | 0.080 |
| Mean Cortical Thickness | -0.05 | 0.668 | 0.701 | -0.22 | 0.969 | 0.969 | -1.32 | 0.187 | 0.223 |
| Estimated Intracranial Volume | 0.79 | 1.11e-16 | 3.04e-16 | 0.76 | 7.51e-15 | 1.97e-14 | -1.94 | 0.052 | 0.072 |
| Subcortical Gray Matter Volume | 0.66 | 1.46e-10 | 3.40e-10 | 0.71 | 2.32e-12 | 5.61e-12 | 0.91 | 0.360 | 0.398 |
| Cortical Volume | 0.61 | 1.08e-08 | 2.34e-08 | 0.49 | 7.32e-06 | 1.36e-05 | -1.53 | 0.126 | 0.165 |
| Cerebral White Matter Volume | 0.24 | 0.024 | 0.036 | 0.30 | 0.006 | 0.010 | 2.11 | 0.035 | 0.051 |
| Total Brain Volume | 0.56 | 1.43e-07 | 2.90e-07 | 0.66 | 1.98e-10 | 4.46e-10 | 3.38 | 7.27e-04 | 0.001 |
| Brain measure | Standard Axial 64mT ICCs with 3T |  |  | SynthSR-Processed Multi-Orientation 64mT ICCs with 3T |  |  | Steiger's |  |  |
|  | ICC | p | q | ICC | p | q | Z | p | q |
| Total Surface Area | **0.71** | **1.65e-12** | **4.15e-12** | **0.87** | **3.67e-23** | **1.78e-22** | **3.50** | **4.72e-04** | **7.62e-04** |
| Mean Cortical Thickness | -0.05 | 0.668 | 0.701 | 0.12 | 0.160 | 0.194 | 1.07 | 0.282 | 0.324 |
| Estimated Intracranial Volume | **0.79** | **1.11e-16** | **3.04e-16** | **0.83** | **2.01e-19** | **7.04e-19** | 1.45 | 0.147 | 0.185 |
| Subcortical Gray Matter Volume | **0.66** | **1.46e-10** | **3.40e-10** | **0.86** | **2.58e-22** | **1.16e-21** | **3.91** | **9.17e-05** | **1.52e-04** |
| Cortical Volume | **0.61** | **1.08e-08** | **2.34e-08** | **0.84** | **5.64e-20** | **2.09e-19** | **4.45** | **8.54e-06** | **1.54e-05** |
| Cerebral White Matter Volume | **0.24** | **0.024** | **0.036** | **0.92** | **2.41e-29** | **1.69e-28** | **8.47** | **2.45e-17** | **7.34e-17** |
| Total Brain Volume | **0.56** | **1.43e-07** | **2.90e-07** | **0.96** | **1.56e-38** | **1.64e-37** | **8.43** | **3.42e-17** | **9.80e-17** |

**Figure 4.**
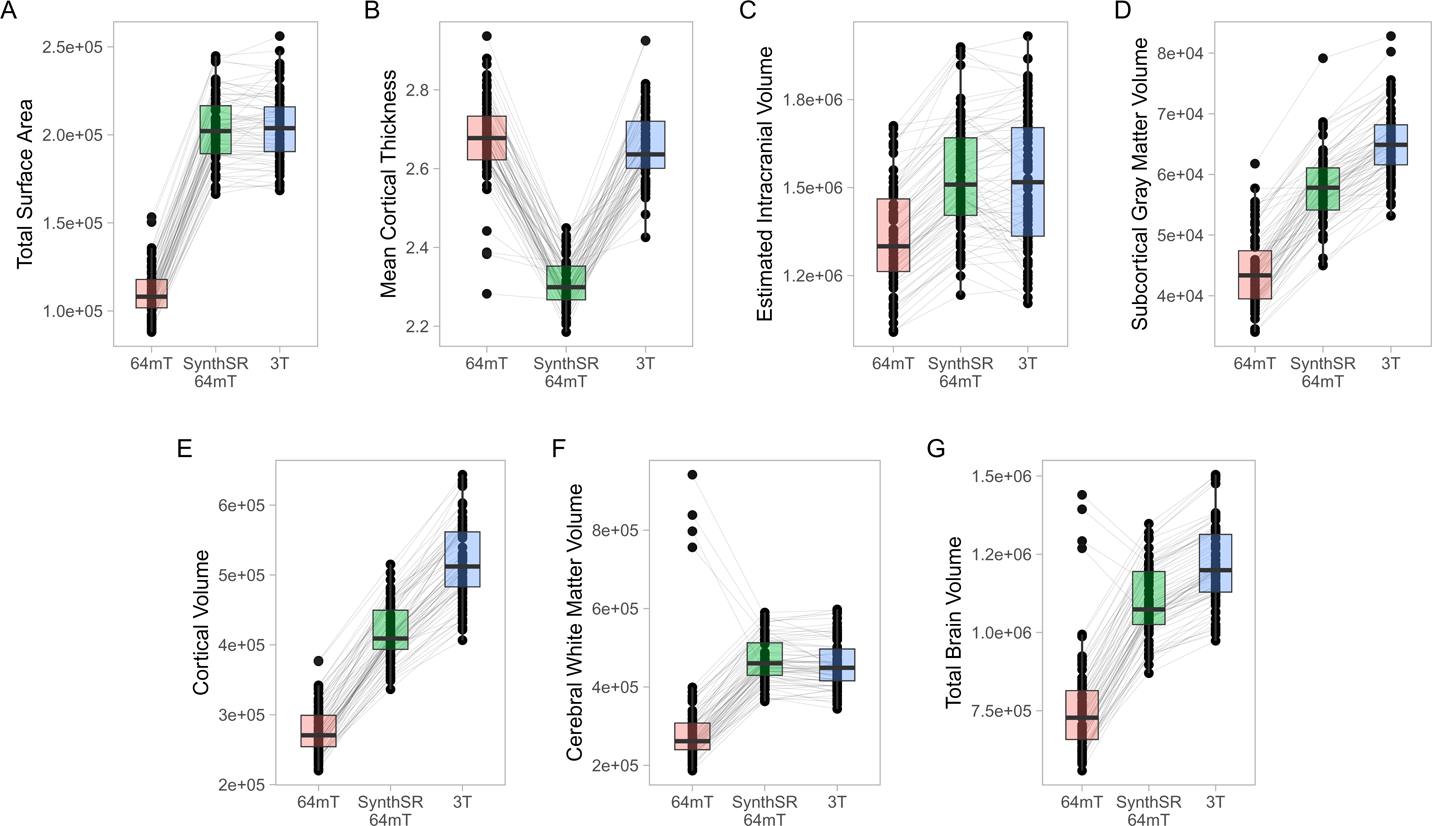
Comparison of individual-level global measurements across standard 64mT axial scans, SynthSR-processed 64mT axial scans, and traditional 3T scans. Boxplots show individual estimates for global brain measures (A-G) derived from standard axial 64mT scans (64mT), SynthSR-processed axial 64mT scans (SynthSR 64mT) and 3T scans (3T). Gray lines connect the same individuals across the three approaches.

At the regional level, we found moderate correspondence across widespread cortical regions for surface area (mean Pearson’s *r* across all 68 regions = 0.35, range = -0.23-0.62; mean ICC = 0.34, range = -0.16-0.59) and cortical volume (mean *r* across all 68 regions = 0.32, range = -0.24-0.64; mean ICC = 0.31, range -0.21-0.58), whereas estimates for cortical thickness were either poor, null or negative (mean *r* across all 68 regions = - 0.04, range = -0.37-0.33; mean ICC = -0.03, range = -0.31-0.29, Figure 2, Supplemental Tables 2 and 3). In particular, the brainstem *(r =* 0.79, *p* = 3.29e-16, *q* = 7.21e-14, ICC = 0.78, p = 2.74e-16, q = 4.29e-15) and lateral ventricle (right, *r* = 0.92, *p* = 3.67e-29, *q* = 6.67e-27; ICC = 0.92, p = 1.53e-29, q = 2.79e-27; left, *r* = 0.96, *p* = 9.47e-38, *q* = 2.58e-35; ICC = 0.95, p = 2.89e-36, q = 7.86e-34) low-field estimates showed high fidelity to measures from high-field scans (Figure 2, Figure 3A). For surface area and cortical volume, the strongest positive correlations were observed in prefrontal (area, left rostral middle frontal, *r* = 0.61, *p* = 2.61e-08, *q* = 1.31e-07; ICC = 0.56, p = 2.02e-07, q = 9.38e-07), occipital (volume, right lateral occipital, *r* = 0.57, *p* = 3.13e-07, *q* = 1.36e-06; ICC = 0.56, p = 1.46e-07, q = 6.91e-07) and right anterior cingulate regions (area, *r* = 0.62, *p* = 9.78e-09, *q* = 5.24e-08; ICC =0.49, p = 8.19e-06, q = 2.97e-05; volume, *r* = 0.64, *p* = 2.45e-09, *q* = 1.39e-08; ICC = 0.58, p = 6.08e-08, q = 2.99e-07; see Supplemental Tables 2 and 3).

### Application of SynthSR significantly improves correspondence between neuroimaging measures extracted from single orientation low-field scans and high-field scans

When we applied SynthSR to single T1/T2 pairs of low-field-acquired images, we found significantly improved correspondence with high-field images for most global brain measures, with the exception of mean cortical thickness and intracranial volume (see Table 2 and Table 3). We observed statistically significant improvements for global estimates of cerebral white matter volume (standard image Pearson’s *r* = 0.32, SynthSR-processed image *r* = 0.92; Steiger’s *z* = 8.75, *p =* 2.18e-18, *q =* 6.88e-18), total brain volume (standard *r* = 0.60, SynthSR-processed *r* = 0.97; *z =* 9.80, *p =* 1.15e-22, *q =* 5.59e-22), cortical volume (standard *r* = 0.67, SynthSR-processed *r* = 0.86; *z =* 4.18, *p =* 2.90e-05, *q =* 5.08e-05), subcortical gray matter volume (standard *r* = 0.66, SynthSR-processed *r* = 0.88; *z =* 4.57, *p =* 4.84e-06, *q =* 8.97e-06) and total surface area (standard *r* = 0.76, SynthSR-processed *r* = 0.90; *z =* 3.73, *p =* 1.94e-04, *q =* 3.22e-04). With the exception of cortical thickness, the ICC also showed significant improvements in correspondence, with ICC coefficients for SynthSR-processed images in the range of 0.84-0.97 (Table 3). When we examined the individual data distributions (Figure 4), we found that processing low-field scans with SynthSR improved correspondence by increasing global brain estimates to more closely approximate the high-field data. Mean differences (SynthSR subtracting 3T) for estimates of surface area were -0.4% (SD 4.1), cortical volume - 19.4% (4.4), subcortical volume -11.4% (4.2), cerebral white matter volume +1.8% (5.6), total brain volume - 9.4% (2.3) and estimated intracranial volume +1.3% (7.2). However, for cortical thickness, SynthSR decreased global estimates, resulting in poorer correspondence with high-field values (mean difference for standard low-field 0.85% [5.4], mean difference for SynthSR-processed low-field -12.9% [3.7]). Bland-Altman plots of agreement (see Supplemental Figure 3) showed positive bias for total brain volume (1.14e05mm^3^, 95% limits of agreement [0.52e5-1.75e5]), cortical volume (1.02e05mm^3^ [0.44e05-1.59e05]), subcortical volume (2.12e04mm^3^ [1.94e3-12.98e3]), and cortical thickness (0.35mm [0.14-0.56]), indicating that SynthSR-processed low-field images still underestimated 3T data. There was no or minimal bias for total surface area (977mm^2^, 95% limits of agreement -1.65e04-1.85e04]), intracranial volume (−1.29e04mm^3^ [-2.09e05-1.83e05]), and cerebral white matter volume (−7.03e03mm^3^ [-55.96e03-41.89e03]). At the regional level, processing with SynthSR significantly improved cortical thickness correspondence (via Pearson correlations) within temporal (*z* range = 2.18-4.08; Figure 3C, Supplemental Table 2) and prefrontal regions (*z* range = 2.78-2.85), with similar improvements when assessed via ICC (temporal z range 2.65-4.02; prefrontal z range 2.44-2.68; Supplemental Table 3). In addition, using SynthSR yielded widespread improvements in image correspondence (assessed via Pearson correlations) for regional estimates of cortical surface area (mean *z* = 2.55, range = -0.85-5.70), cortical volume (mean *z* = 2.51, range = -0.55-6.18) and subcortical volume (mean *z* = 2.50, range = -1.67-5.15), with the strongest effects observed in the left superior temporal lobe (area, z = 5.45; volume, z = 6.18) and precentral gyri supplementary motor areas (area, *z* = 5.70; volume, *z* = 5.42; Figure 3C, Supplemental Table 2; corresponding ICC values can be found in Supplemental Table 3).

### A multi-orientation approach achieves modest improvements on a single-orientation approach for some neuroimaging measures

When we used a multi-orientation (N = six pairs of T1/T2 multi-orientation scans) image averaging approach, we found modest and significantly improved correspondence to high-field scans for global estimates of cerebral white matter volume as assessed via Pearson’s correlation (*z =* 3.57, *p =* 3.64e-04, *q =* 5.73e-04) and total brain volume (*z =* 3.49, *p =* 4.84e-04, *q =* 7.44e-04; Table 2). In contrast, we found a negative relationship for mean cortical thickness, with higher cortical thickness values in the 3T corresponding to lower total surface area (*z =* -2.52, *p =* .012, *q =* .016) and estimated intracranial volume (*z =* -2.36, *p =* .018, *q =* .024; Table 2) values in the low-field data. We did not see a significant improvement in global estimates of cortical thickness, subcortical and cortical volume when we increased the number of scan acquisitions. Examination of the data distributions for cerebral white matter and total brain volume (Supplemental Figure 4) revealed that the multi-orientation approach increased the correspondence for all scans in a relatively uniform manner. In contrast, there were highly varied outcomes for the remaining brain measures (i.e., surface area, cortical thickness, subcortical and cortical gray matter volume), whereby some scans improved while others decreased in correspondence to the high-field estimates.

At the regional level, when we used a multi-orientation approach, correspondence (via Pearson correlations) became more negative for cortical thickness in several brain regions, including the left rostral middle frontal (*z =* -2.30, *p =* .021, *q =* .040) and bilateral supramarginal gyri (left, *z =* -2.69, *p =* .007, *q =* .015; right, *z =* -3.57, *p =* 3.63e-04, *q =* 9.60e-04; Figure 3D, Supplemental Table 4; corresponding ICC values can be found in Supplemental Table 5). Correlations also became significantly more negative in the right middle temporal gyrus for estimates of cortical surface area (*z =* -2.26, *p =* .024, *q =* .044) and cortical volume (*z =* - 2.23, *p =* .026, *q =* .047).

### A combination of SynthSR and multi-orientation approaches achieved significant improvements in image correspondence over a standard single-pair acquisition

When we combined the multi-orientation (N = six T1/T2 multi-orientation scan pairs) and SynthSR processing approaches, we found significantly improved correspondence compared to a standard single-pair acquisition for almost all brain measures (Table 2 and Table 3). Following this combined approach, almost all global measures showed excellent correspondence (Pearson *r*’s > 0.85, ICC values > 0.84) with high-field images, with the exception of mean cortical thickness, of which correspondence was not significantly improved following additional processing (Pearson’s r = 0.12, p = 0.317, q = 0.363; ICC = 0.12, p = 0.160, q = 0.194).

We found that the combined approach resulted in uniform increases in global estimates across all individuals, with cortical thickness as the exception (Supplemental Figure 5). At a regional level, the combined approach improved correspondence for surface area and volume across widespread cortical and subcortical regions, with strongest effects observed in the temporal lobe and supplementary motor areas (Figure 3G).

### Multi-orientation scans processed with SynthSR improved image correspondence over single-acquisition scans processed with SynthSR in localized regions of the cortex

Finally, we compared the difference in correspondence between single-acquisition (N = one T1/T2 pair) scans processed with SynthSR and multi-orientation (N = six T1/T2 pairs) scans processed with SynthSR. We found that this approach decreased the correspondence for estimates of global surface area (*z* = -2.16, *p* = .031, *q* = .042), but did not significantly influence other global measures (Supplemental Table 6). At a regional level, increasing the number of scans acquired and processed with SynthSR improved the correspondence of regions primarily within the cingulate cortex (Supplemental Figure 6).

### Correlations with age and motion

We tested whether age or motion were associated with individual-level differences between low- and high-field estimates of each brain measure. After correction for multiple comparisons, there were no statistically significant relationships between the individual-level differences in high- and low-field data and participant motion (See Supplemental Table 7). Age was negatively correlated with discrepancies in estimates of cortical thickness following processing with SynthSR (Pearson’s *r* = -0.59, *p* = 8.36e-08, *q* = 2.73e-06; Supplemental Table 8), and was also negatively correlated with discrepancies in estimates of cortical volume across all processing approaches. This indicates that the younger the participant, the greater the discrepancy between low-field and the 3T estimates. Further, we observed a negative correlation between age and discrepancies in estimates of surface area in the standard acquisition (Pearson’s *r* = -0.59, *p* = 8.36e-08, *q* = 2.73e-06), which was not present in images processed with SynthSR.

## Discussion

In this study, we evaluated the correspondence between MR images acquired from low- and high-field scanners in a community sample of young people. We found that pairs of T1- and T2-weighted images obtained at low-field (64mT) showed high fidelity to images acquired at high-field (3T) for most global and regional brain measures, most notably measures of surface area, cortical volume, and subcortical volume. When we implemented a novel super-resolution method via a CNN (i.e., SynthSR, Iglesias et al., 2021), we found significantly improved image correspondence across almost all brain measures. When we increased the number of low-field scan acquisitions and orientations, we observed improvements in low-field image correspondence for estimates of cerebral white matter volume and total brain volume. The discrepancy between low-field and 3T measurements was not related to motion during the low-field scan session. These results demonstrate the potential of low-field imaging in young people and provide a foundation for future work to further improve low-field image quality and fidelity.

We found that standard single-acquisition low-field scans showed high correspondence with high-field-acquired scans for estimates of intracranial volume (*r* = 0.82) and surface area (*r* = 0.76), and moderate correspondence for estimates of subcortical, cortical gray and total brain volume (*r* = 0.60–0.67). These findings support previous work in children and adolescents demonstrating high fidelity between low-field and high-field scans for estimates of global gray matter, white matter, and total intracranial volume (Deoni et al., 2021). Further, we extend previous work by characterizing the regional variation in LF-3T correspondence in cortical surface area, cortical volume, and subcortical volume, identifying regions with the greatest fidelity to high-field scans. Regions with the strongest correlations between low- and high-field tended to be larger and are typically high-contrast (i.e., the brainstem and ventricles), consistent with previous work demonstrating greater image fidelity for low-field data within larger brain regions (Iglesias et al., 2023). Within cortical regions, the greater correspondence within temporal, prefrontal, and cingulate lobes are important developmentally as these regions show protracted maturational trajectories (Amlien et al., 2016; Baum et al., 2020) and are commonly implicated in mental health disorders (Jones et al., 2017). The considerably lower cost and increased accessibility offered by low-field technology, coupled with demonstrated correspondence with high-field scans, provide a compelling argument for implementation of low-field technology at scale among young people.

We found that processing scans via SynthSR significantly improved image correspondence for global and regional estimates of gray and white matter volume, subcortical volume, surface area and total brain volume in young people. Regions that showed the greatest improvements included temporal and prefrontal regions, parts of the brain which have a developmental peak last, as opposed to sensory and motor cortices, which reach maturation at earlier points in development (Larsen et al., 2023; Sydnor et al., 2023, 2023). Our findings build on and extend recent work in adults demonstrating high correspondence between low- and high-field images following processing with SynthSR (Iglesias et al., 2021). While no incidental findings were reported in this study, future work should seek to investigate how SynthSR and other super-resolution methods might help to improve the ability to identify these findings in clinical and non-clinical samples.

We found that individual-level differences in estimates between low-field and high-field data were negatively correlated with age for cortical thickness and cortical volume several brain measures, indicating that with both standard and SynthSR-processed low-field estimates data were more accurate in older individuals for these measures. This might be partly explained by the existing reference library used to build SynthSR, which was built on an adult sample (Iglesias et al., 2021). Alternatively, age-associated patterns in gray-white matter contrast may contribute to the ability to differentiate the pial surface and white matter boundary at differing ages (Norbom et al., 2019, 2021). However, on the whole, despite being derived from adult training data, SynthSR still performed quite well in improving image correspondence in our sample, which included youth from middle childhood, adolescents, and young adults. These results demonstrate the validity of using SynthSR in estimating global and regional cortical surface area, cortical volume, and subcortical volume measures in younger populations.

We found that implementing a multi-orientation scan series yielded modest improvements in image correspondence for estimates of cerebral white matter and total brain volume. However, these improvements were not observed in other brain measures. There was decreased correspondence for estimates of surface area and intracranial volume, and null effects for cortical thickness. While our approach did not show the same degree of improvement displayed in previous work (Askin Incebacak et al., 2022; Sui et al., 2021), we note that these studies were acquired with a high-field (3T) magnet, in contrast to the low-field magnet used in our study.

It is also possible that we could employ alternative co-registration procedures to improve this correspondence, as we only implemented one type of co-registration in this study (i.e., an affine transformation registered to the axial image). In future, we may also achieve improved low-field and 3T measure correspondence if we employ alternative multi-orientation approaches. These approaches might include, for example, combining a multi-orientation approach with variable echo times (Deoni, O’Muircheartaigh, et al., 2022), or obtaining multiple scans in a single orientation (Arnold et al., 2022). The variable results we observed across brain measures may suggest that specific super-resolution approaches might be more effective for specific brain tissues — further research is necessary to investigate this possibility.

In this sample, neither super-resolution approach improved the correspondence of low-field estimates for total mean cortical thickness. However, at the regional level, processing with SynthSR yielded significant improvements in correspondence within the prefrontal and temporal lobes; for example, correspondence increased from *r* = -0.18 to *r* = 0.43 within the right temporal lobe following processing with SynthSR.As cortical thickness is measured at the sub-millimeter level, changes in voxel size can greatly affect those measurements. As a result, estimation of cortical thickness remains challenging at low field, whereby images are acquired with larger voxel size and slice thickness than traditional high-field-strength systems (Iglesias et al., 2021). In addition, the lower contrast-to-noise ratio of low-field images increases the difficulty for software to accurately estimate gray and white matter boundaries, which are required for accurate estimation of cortical thickness (Fischl, 2012). In order to address these challenges, further refinement of acquisition and processing pipelines, or deployment of alternative super-resolution approaches, will be necessary to successfully recapitulate estimates of cortical thickness in low-field-strength systems. We are rigorously investigating additional methods at this time. Improvements to the measurement of cortical thickness with low-field MRI are important for implementation of low-field MR systems among youth and young adults, given the marked changes in cortical thickness that occur during this developmental period (Tamnes et al., 2010; Vijayakumar et al., 2016) and its relevance to several psychiatric disorders (Cheon et al., 2022; Hettwer et al., 2022). One approach may be to leverage the advantages of both low- and high-strength MRI systems by employing a sequential staging approach, using low-field systems at scale for initial identification of high-risk cases, and reserving high-field MR for later classification of true-positive from false-positive findings (Koutsouleris et al., 2021). Alternatively, one promising approach may be to use measurement-in-error statistical modeling. In other fields of medicine, measurement-in-error statistical modeling is often used to develop low-cost, convenient measures for risk assessment (Collins et al., 1990; Cook et al., 1998; MacMahon et al., 1990). Firstly, a functional relationship is established between a ‘gold standard’ measure (e.g., 3T-acquired image) and a noisier, low-cost, convenient measure (e.g., LF-acquired image) for risk assessment (Carroll et al., 2006).This model is then transported to an external sample where a proxy estimate of the gold standard measure is obtained using only the noisier measure. We plan to test the feasibility of this approach once we have collected sufficient data in an independent sample of youth.

Characterization of the diverse neurodevelopmental trajectories in mental health and illness is integral to understanding the etiology of mental disorders in youth. Despite the advent of ‘big data’ and the emergence of several large neuroimaging datasets, most samples only include individuals who can travel to fixed MR scanners within urban medical and research centers (Garcini et al., 2022; Shen et al., 2021), commonly termed “samples of convenience.” Systemic and structural barriers prohibiting widespread MR access limit the diversity and generalizability of neuroimaging samples, which may act to reinforce systemic biases in inference and interpretation (Garcini et al., 2022). Low-field technology offers an opportunity to improve the accessibility of MR technology to underserved populations, thereby addressing systemic biases in healthcare access and in representation within neuroscience (Garcini et al., 2022; Shen et al., 2021). Also, for neuroimaging to have practical prognostic and diagnostic utility, we must be able to obtain MRI scans in a variety of settings, not only in urban research centers. Incorporating community settings into risk assessment programs is crucial to identify at-risk individuals earlier, reach individuals traditionally underserved by medical research centers, and progress towards the goal of universal screening and prevention (Garner et al., 2012; Murray et al., 2021; Schneider et al., 2020). This approach will also satisfy service users’ preference for community settings and easily implemented assessments (Schneider et al., 2020). Finally, in young people in particular, increasing diversity in neuroscience is essential to improving our ability to understand inter-individual differences in brain development, identify emerging neurodevelopmental and psychiatric disorders, and discover biomarkers that are generalizable to diverse populations (Burkhard et al., 2021; Garcini et al., 2022).

We must acknowledge the limitations of this study. This study was conducted in healthy children, adolescents, and young adults; further work is necessary to test the generalizability of these findings to clinical populations. Further, we assumed high-field acquired images as the “ground truth”, although the accuracy of high-field MR images in estimating structural brain measures has not been established. While we used pairs of T1- and T2-weighted images, previous low-field studies have used only T2-weighted scans (Deoni et al., 2021); thus, additional work is necessary to quantitatively compare and evaluate these contrasting approaches. We did not apply distortion correction or concomitant field correction for either low-field or high-field scans; employing these correction strategies may help to further improve image quality, as has been demonstrated elsewhere (Thaler et al., 2023). In addition, our selected acquisition parameters and image processing and super-resolution software were not specifically developed for pediatric populations; developmentally-specific software has yet to be developed, and may further improve image fidelity and quality.

In summary, we found that brain structural images acquired in a sample of young people in a portable low-field MR system showed high fidelity to high-field MR images for measures of brain volume and surface area. Additionally, we found that greater correspondence to high-field images could be achieved for cerebral white matter volume and subcortical volume following processing via a CNN developed for low-field images. In contrast, using a multi-orientation image averaging approach resulted in modest improvements in image correspondence for measures of white matter volume and total brain volume, but resulted in lower correspondence for surface area and intracranial volume. Finally, we found that using a combined multi-orientation and CNN-processing approach significantly improved image correspondence when compared to standard single-acquisition scans, but negligible improvements in correspondence above single-acquisition, CNN-processed scans. Taken together, our results indicate that using single pairs of T1- and T2-weighted images, combined with super-resolution of images via SynthSR, yielded the greatest improvements in correspondence between low-field and high-field MR images. Future work should seek to evaluate different combinations of acquisition and processing approaches to further improve images acquired with portable low-field systems.

## Data Availability

All data produced in the present study are available upon reasonable request to the authors.

## Acknowledgements

The authors would like to thank Beatriz Horta and Rashmi Sahasrabudhe for their contributions in reading and providing feedback on drafts of the manuscript. The authors would also like to thank Emma Waite, who was involved in the recruitment of participants and data collection procedures.

High-field MRI scans were obtained at Boston Children’s Hospital at 2 Brookline Place. Funding for this study was supported by the Office Of The Director, National Institutes Of Health of the National Institutes of Health under Award Number S10OD025111. The content is solely the responsibility of the authors and does not necessarily represent the official views of the National Institutes of Health.

## References

1. Amlien, I. K., Fjell, A. M., Tamnes, C. K., Grydeland, H., Krogsrud, S. K., Chaplin, T. A., Rosa, M. G. P., & Walhovd, K. B. (2016). Organizing Principles of Human Cortical Development--Thickness and Area from 4 to 30 Years: Insights from Comparative Primate Neuroanatomy. *Cerebral Cortex (New York*, N.Y.: 1991), *26*(1), 257–267. 10.1093/cercor/bhu214

2. Andreou, C., & Borgwardt, S. (2020). Structural and functional imaging markers for susceptibility to psychosis. Molecular Psychiatry, 25(11), 2773–2785. 10.1038/s41380-020-0679-7

3. Arnold, T. C., Freeman, C. W., Litt, B., & Stein, J. M. (2023). Low-field MRI: Clinical promise and challenges. Journal of Magnetic Resonance Imaging: JMRI, 57(1), 25–44. 10.1002/jmri.28408

4. Arnold, T. C., Tu, D., Okar, S. V., Nair, G., By, S., Kawatra, K. D., Robert-Fitzgerald, T. E., Desiderio, L. M., Schindler, M. K., Shinohara, R. T., Reich, D. S., & Stein, J. M. (2022). Sensitivity of portable low-field magnetic resonance imaging for multiple sclerosis lesions. NeuroImage. Clinical, 35, 103101. 10.1016/j.nicl.2022.103101

5. Askin Incebacak, N. C., Sui, Y., Gui Levy, L., Merlini, L., Sa de Almeida, J., Courvoisier, S., Wallace, T. E., Klauser, A., Afacan, O., Warfield, S. K., Hüppi, P., & Lazeyras, F. (2022). Super-resolution reconstruction of T2-weighted thick-slice neonatal brain MRI scans. Journal of Neuroimaging: Official Journal of the American Society of Neuroimaging, 32(1), 68–79. 10.1111/jon.12929

6. Avants, B. B., Epstein, C. L., Grossman, M., & Gee, J. C. (2008). Symmetric diffeomorphic image registration with cross-correlation: Evaluating automated labeling of elderly and neurodegenerative brain. Medical Image Analysis, 12(1), 26–41. 10.1016/j.media.2007.06.004

7. Baum, G. L., Cui, Z., Roalf, D. R., Ciric, R., Betzel, R. F., Larsen, B., Cieslak, M., Cook, P. A., Xia, C. H., Moore, T. M., Ruparel, K., Oathes, D. J., Alexander-Bloch, A. F., Shinohara, R. T., Raznahan, A., Gur, R. E., Gur, R. C., Bassett, D. S., & Satterthwaite, T. D. (2020). Development of structure-function coupling in human brain networks during youth. Proceedings of the National Academy of Sciences of the United States of America, 117(1), 771–778. 10.1073/pnas.1912034117

8. Beck, A. T., Steer, R. A., & Carbin, M. G. (1988). Psychometric properties of the Beck Depression Inventory: Twenty-five years of evaluation. Clinical Psychology Review, 8(1), 77–100. 10.1016/0272-7358(88)90050-5

9. Benjamini, Y., & Hochberg, Y. (1995). Controlling the False Discovery Rate: A Practical and Powerful Approach to Multiple Testing. Journal of the Royal Statistical Society. Series B (Methodological), 57(1), 289–300.

10. Bland, J. M., & Altman, D. G. (1999). Measuring agreement in method comparison studies. Statistical Methods in Medical Research, 8(2), 135–160. 10.1177/096228029900800204

11. Burkhard, C., Cicek, S., Barzilay, R., Radhakrishnan, R., & Guloksuz, S. (2021). Need for Ethnic and Population Diversity in Psychosis Research. Schizophrenia Bulletin, 47(4), 889–895. 10.1093/schbul/sbab048

12. Carroll, R. J., Ruppert, D., Stefanski, L. A., & Crainiceanu, C. M. (2006). Measurement Error in Nonlinear Models: A Modern Perspective, Second Edition (2nd ed.). Chapman and Hall/CRC. 10.1201/9781420010138

13. Cheon, E.-J., Bearden, C. E., Sun, D., Ching, C. R. K., Andreassen, O. A., Schmaal, L., Veltman, D. J., Thomopoulos, S. I., Kochunov, P., Jahanshad, N., Thompson, P. M., Turner, J. A., & van Erp, T. G. M. (2022). Cross disorder comparisons of brain structure in schizophrenia, bipolar disorder, major depressive disorder, and 22q11.2 deletion syndrome: A review of ENIGMA findings. Psychiatry and Clinical Neurosciences, 76(5), 140–161. 10.1111/pcn.13337

14. Chetcuti, K., Chilingulo, C., Goyal, M. S., Vidal, L., O’Brien, N. F., Postels, D. G., Seydel, K. B., & Taylor, T. E. (2022). Implementation of a Low-Field Portable MRI Scanner in a Resource-Constrained Environment: Our Experience in Malawi. AJNR: American Journal of Neuroradiology, 43(5), 670–674. 10.3174/ajnr.A7494

15. Collins, R., Peto, R., MacMahon, S., Hebert, P., Fiebach, N. H., Eberlein, K. A., Godwin, J., Qizilbash, N., Taylor, J. O., & Hennekens, C. H. (1990). Blood pressure, stroke, and coronary heart disease. Part 2, Short-term reductions in blood pressure: Overview of randomised drug trials in their epidemiological context. Lancet (London, England), 335(8693), 827–838. 10.1016/0140-6736(90)90944-z

16. Cook, N. R., Kumanyika, S. K., & Cutler, J. A. (1998). Effect of change in sodium excretion on change in blood pressure corrected for measurement error. The Trials of Hypertension Prevention, Phase I. American Journal of Epidemiology, 148(5), 431–444. 10.1093/oxfordjournals.aje.a009668

17. Deoni, S. C. L., Bruchhage, M. M. K., Beauchemin, J., Volpe, A., D’Sa, V., Huentelman, M., & Williams, S. C. R. (2021). Accessible pediatric neuroimaging using a low field strength MRI scanner. NeuroImage, 238, 118273. 10.1016/j.neuroimage.2021.118273

18. Deoni, S. C. L., Medeiros, P., Deoni, A. T., Burton, P., Beauchemin, J., D’Sa, V., Boskamp, E., By, S., McNulty, C., Mileski, W., Welch, B. E., & Huentelman, M. (2022). Development of a mobile low-field MRI scanner. Scientific Reports, 12(1), Article 1. 10.1038/s41598-022-09760-2

19. Deoni, S. C. L., O’Muircheartaigh, J., Ljungberg, E., Huentelman, M., & Williams, S. C. R. (2022). Simultaneous high-resolution T2 -weighted imaging and quantitative T2 mapping at low magnetic field strengths using a multiple TE and multi-orientation acquisition approach. Magnetic Resonance in Medicine, 88(3), 1273–1281. 10.1002/mrm.29273

20. Desikan, R. S., Ségonne, F., Fischl, B., Quinn, B. T., Dickerson, B. C., Blacker, D., Buckner, R. L., Dale, A. M., Maguire, R. P., Hyman, B. T., Albert, M. S., & Killiany, R. J. (2006). An automated labeling system for subdividing the human cerebral cortex on MRI scans into gyral based regions of interest. NeuroImage, 31(3), 968–980. 10.1016/j.neuroimage.2006.01.021

21. Ellis, J. K., Walker, E. F., & Goldsmith, D. R. (2020). Selective Review of Neuroimaging Findings in Youth at Clinical High Risk for Psychosis: On the Path to Biomarkers for Conversion. Frontiers in Psychiatry, 11. https://www.frontiersin.org/articles/10.3389/fpsyt.2020.567534

22. Esteban, O., Birman, D., Schaer, M., Koyejo, O. O., Poldrack, R. A., & Gorgolewski, K. J. (2017). MRIQC: Advancing the automatic prediction of image quality in MRI from unseen sites. PloS One, 12(9), e0184661. 10.1371/journal.pone.0184661

23. Fairman, K. A., Peckham, A. M., & Sclar, D. A. (2020). Diagnosis and Treatment of ADHD in the United States: Update by Gender and Race. Journal of Attention Disorders, 24(1), 10–19. 10.1177/1087054716688534

24. Figueiro Longo, M. G., Jaimes, C., Machado, F., Delgado, J., & Gee, M. S. (2022). Pediatric Emergency MRI. Magnetic Resonance Imaging Clinics of North America, 30(3), 533–552. 10.1016/j.mric.2022.05.004

25. Fischl, B. (2012). FreeSurfer. NeuroImage, 62(2), 774–781. 10.1016/j.neuroimage.2012.01.021

26. Garcini, L. M., Arredondo, M. M., Berry, O., Church, J. A., Fryberg, S., Thomason, M. E., & McLaughlin, K. A. (2022). Increasing diversity in developmental cognitive neuroscience: A roadmap for increasing representation in pediatric neuroimaging research. Developmental Cognitive Neuroscience, 58, 101167. 10.1016/j.dcn.2022.101167

27. Garner, A. S., Shonkoff, J. P., Committee on Psychosocial Aspects of Child and Family Health, Committee on Early Childhood, Adoption, and Dependent Care, & Section on Developmental and Behavioral Pediatrics. (2012). Early childhood adversity, toxic stress, and the role of the pediatrician: Translating developmental science into lifelong health. Pediatrics, 129(1), e224–231. 10.1542/peds.2011-2662

28. Grasby, K. L., Jahanshad, N., Painter, J. N., Colodro-Conde, L., Bralten, J., Hibar, D. P., Lind, P. A., Pizzagalli, F., Ching, C. R. K., McMahon, M. A. B., Shatokhina, N., Zsembik, L. C. P., Thomopoulos, S. I., Zhu, A. H., Strike, L. T., Agartz, I., Alhusaini, S., Almeida, M. A. A., Alnæs, D., … Enhancing NeuroImaging Genetics through Meta-Analysis Consortium (ENIGMA)—Genetics working group. (2020). The genetic architecture of the human cerebral cortex. Science (New York, N.Y.), 367(6484). 10.1126/science.aay6690

29. Guallart-Naval, T., Algarín, J. M., Pellicer-Guridi, R., Galve, F., Vives-Gilabert, Y., Bosch, R., Pallás, E., González, J. M., Rigla, J. P., Martínez, P., Lloris, F. J., Borreguero, J., Marcos-Perucho, Á., Negnevitsky, V., Martí-Bonmatí, L., Ríos, A., Benlloch, J. M., & Alonso, J. (2022). Portable magnetic resonance imaging of patients indoors, outdoors and at home. Scientific Reports, 12(1), Article 1. 10.1038/s41598-022-17472-w

30. Herting, M. M., Johnson, C., Mills, K. L., Vijayakumar, N., Dennison, M., Liu, C., Goddings, A.-L., Dahl, R. E., Sowell, E. R., Whittle, S., Allen, N. B., & Tamnes, C. K. (2018). Development of subcortical volumes across adolescence in males and females: A multisample study of longitudinal changes. NeuroImage, 172, 194–205. 10.1016/j.neuroimage.2018.01.020

31. Hettwer, M. D., Larivière, S., Park, B. Y., van den Heuvel, O. A., Schmaal, L., Andreassen, O. A., Ching, C. R. K., Hoogman, M., Buitelaar, J., van Rooij, D., Veltman, D. J., Stein, D. J., Franke, B., van Erp, T. G. M., ENIGMA ADHD Working Group, ENIGMA Autism Working Group, ENIGMA Bipolar Disorder Working Group, ENIGMA Major Depression Working Group, ENIGMA OCD Working Group, … Valk, S. L. (2022). Coordinated cortical thickness alterations across six neurodevelopmental and psychiatric disorders. Nature Communications, 13(1), 6851. 10.1038/s41467-022-34367-6

32. Iglesias, J. E., Billot, B., Balbastre, Y., Tabari, A., Conklin, J., Gilberto González, R., Alexander, D. C., Golland, P., Edlow, B. L., Fischl, B., & Alzheimer’s Disease Neuroimaging Initiative. (2021). Joint super-resolution and synthesis of 1 mm isotropic MP-RAGE volumes from clinical MRI exams with scans of different orientation, resolution and contrast. NeuroImage, 237, 118206. 10.1016/j.neuroimage.2021.118206

33. Iglesias, J. E., Schleicher, R., Laguna, S., Billot, B., Schaefer, P., McKaig, B., Goldstein, J. N., Sheth, K. N., Rosen, M. S., & Kimberly, W. T. (2023). Quantitative Brain Morphometry of Portable Low-Field-Strength MRI Using Super-Resolution Machine Learning. Radiology, 306(3), e220522. 10.1148/radiol.220522

34. Jones, S. A., Morales, A. M., Lavine, J. B., & Nagel, B. J. (2017). Convergent neurobiological predictors of emergent psychopathology during adolescence. Birth Defects Research, 109(20), 1613–1622. 10.1002/bdr2.1176

35. Kimberly, W. T., Sorby-Adams, A. J., Webb, A. G., Wu, E. X., Beekman, R., Bowry, R., Schiff, S. J., de Havenon, A., Shen, F. X., Sze, G., Schaefer, P., Iglesias, J. E., Rosen, M. S., & Sheth, K. N. (2023). Brain imaging with portable low-field MRI. Nature Reviews Bioengineering, 1(9), 617–630. 10.1038/s44222-023-00086-w

36. Klein, A., Andersson, J., Ardekani, B. A., Ashburner, J., Avants, B., Chiang, M.-C., Christensen, G. E., Collins, D. L., Gee, J., Hellier, P., Song, J. H., Jenkinson, M., Lepage, C., Rueckert, D., Thompson, P., Vercauteren, T., Woods, R. P., Mann, J. J., & Parsey, R. V. (2009). Evaluation of 14 nonlinear deformation algorithms applied to human brain MRI registration. NeuroImage, 46(3), 786–802. 10.1016/j.neuroimage.2008.12.037

37. Koutsouleris, N., Dwyer, D. B., Degenhardt, F., Maj, C., Urquijo-Castro, M. F., Sanfelici, R., Popovic, D., Oeztuerk, O., Haas, S. S., Weiske, J., Ruef, A., Kambeitz-Ilankovic, L., Antonucci, L. A., Neufang, S., Schmidt-Kraepelin, C., Ruhrmann, S., Penzel, N., Kambeitz, J., Haidl, T. K., … PRONIA Consortium. (2021). Multimodal Machine Learning Workflows for Prediction of Psychosis in Patients With Clinical High-Risk Syndromes and Recent-Onset Depression. JAMA Psychiatry, 78(2), 195–209. 10.1001/jamapsychiatry.2020.3604

38. Larsen, B., Sydnor, V. J., Keller, A. S., Yeo, B. T. T., & Satterthwaite, T. D. (2023). A critical period plasticity framework for the sensorimotor–association axis of cortical neurodevelopment. Trends in Neurosciences, 46(10), 847–862. 10.1016/j.tins.2023.07.007

39. Lebel, C., & Deoni, S. (2018). The development of brain white matter microstructure. NeuroImage, 182, 207–218. 10.1016/j.neuroimage.2017.12.097

40. Lee-Jayaram, J. J., Goerner, L. N., & Yamamoto, L. G. (2020). Magnetic Resonance Imaging of the Brain in the Pediatric Emergency Department. Pediatric Emergency Care, 36(12), 586–590. 10.1097/PEC.0000000000002286

41. Lerch, J. P., van der Kouwe, A. J. W., Raznahan, A., Paus, T., Johansen-Berg, H., Miller, K. L., Smith, S. M., Fischl, B., & Sotiropoulos, S. N. (2017). Studying neuroanatomy using MRI. Nature Neuroscience, 20(3), Article 3. 10.1038/nn.4501

42. Lévy, S., Guertin, M.-C., Khatibi, A., Mezer, A., Martinu, K., Chen, J.-I., Stikov, N., Rainville, P., & Cohen-Adad, J. (2018). Test-retest reliability of myelin imaging in the human spinal cord: Measurement errors versus region- and aging-induced variations. PLOS ONE, 13(1), e0189944. 10.1371/journal.pone.0189944

43. MacMahon, S., Peto, R., Cutler, J., Collins, R., Sorlie, P., Neaton, J., Abbott, R., Godwin, J., Dyer, A., & Stamler, J. (1990). Blood pressure, stroke, and coronary heart disease. Part 1, Prolonged differences in blood pressure: Prospective observational studies corrected for the regression dilution bias. Lancet (London, England), 335(8692), 765–774. 10.1016/0140-6736(90)90878-9

44. Marin, J. R., Rodean, J., Hall, M., Alpern, E. R., Aronson, P. L., Chaudhari, P. P., Cohen, E., Freedman, S. B., Morse, R. B., Peltz, A., Samuels-Kalow, M., Shah, S. S., Simon, H. K., & Neuman, M. I. (2021). Racial and Ethnic Differences in Emergency Department Diagnostic Imaging at US Children’s Hospitals, 2016-2019. JAMA Network Open, 4(1), e2033710. 10.1001/jamanetworkopen.2020.33710

45. Mazurek, M. H., Cahn, B. A., Yuen, M. M., Prabhat, A. M., Chavva, I. R., Shah, J. T., Crawford, A. L., Welch, E. B., Rothberg, J., Sacolick, L., Poole, M., Wira, C., Matouk, C. C., Ward, A., Timario, N., Leasure, A., Beekman, R., Peng, T. J., Witsch, J., … Sheth, K. N. (2021). Portable, bedside, low-field magnetic resonance imaging for evaluation of intracerebral hemorrhage. Nature Communications, 12(1), 5119. 10.1038/s41467-021-25441-6

46. Mazurek, M. H., Parasuram, N. R., Peng, T. J., Beekman, R., Yadlapalli, V., Sorby-Adams, A. J., Lalwani, D., Zabinska, J., Gilmore, E. J., Petersen, N. H., Falcone, G. J., Sujijantarat, N., Matouk, C., Payabvash, S., Sze, G., Schiff, S. J., Iglesias, J. E., Rosen, M. S., de Havenon, A., … Sheth, K. N. (2023). Detection of Intracerebral Hemorrhage Using Low-Field, Portable Magnetic Resonance Imaging in Patients With Stroke. Stroke. 10.1161/STROKEAHA.123.043146

47. Murata, S., Hagiwara, A., Kaga, H., Someya, Y., Nemoto, K., Goto, M., Kamagata, K., Irie, R., Hori, M., Andica, C., Wada, A., Kumamaru, K. K., Shimoji, K., Otsuka, Y., Hoshito, H., Tamura, Y., Kawamori, R., Watada, H., & Aoki, S. (2021). Comparison of Brain Volume Measurements Made with 0.3- and 3-T MR Imaging. Magnetic Resonance in Medical Sciences, 21(3), 517–524. 10.2463/mrms.tn.2020-0034

48. Murray, R. M., David, A. S., & Ajnakina, O. (2021). Prevention of psychosis: Moving on from the at-risk mental state to universal primary prevention. Psychological Medicine, 51(2), 223–227. 10.1017/S003329172000313X

49. Narrow, W. E., Clarke, D. E., Kuramoto, S. J., Kraemer, H. C., Kupfer, D. J., Greiner, L., & Regier, D. A. (2013). DSM-5 field trials in the United States and Canada, Part III: Development and reliability testing of a cross-cutting symptom assessment for DSM-5. The American Journal of Psychiatry, 170(1), 71–82. 10.1176/appi.ajp.2012.12071000

50. Norbom, L. B., Doan, N. T., Alnæs, D., Kaufmann, T., Moberget, T., Rokicki, J., Andreassen, O. A., Westlye, L. T., & Tamnes, C. K. (2019). Probing Brain Developmental Patterns of Myelination and Associations With Psychopathology in Youths Using Gray/White Matter Contrast. Biological Psychiatry, 85(5), 389–398. 10.1016/j.biopsych.2018.09.027

51. Norbom, L. B., Ferschmann, L., Parker, N., Agartz, I., Andreassen, O. A., Paus, T., Westlye, L. T., & Tamnes, C. K. (2021). New insights into the dynamic development of the cerebral cortex in childhood and adolescence: Integrating macro- and microstructural MRI findings. Progress in Neurobiology, 204, 102109. 10.1016/j.pneurobio.2021.102109

52. Pontious, A., Kowalczyk, T., Englund, C., & Hevner, R. F. (2008). Role of intermediate progenitor cells in cerebral cortex development. Developmental Neuroscience, 30(1–3), 24–32. 10.1159/000109848

53. Rakic, P. (1988). Specification of cerebral cortical areas. Science (New York, N.Y.), 241(4862), 170–176. 10.1126/science.3291116

54. Raschle, N., Zuk, J., Ortiz-Mantilla, S., Sliva, D. D., Franceschi, A., Grant, P. E., Benasich, A. A., & Gaab, N. (2012). Pediatric neuroimaging in early childhood and infancy: Challenges and practical guidelines. Annals of the New York Academy of Sciences, 1252, 43–50. 10.1111/j.1749-6632.2012.06457.x

55. Reuter, M., Tisdall, M. D., Qureshi, A., Buckner, R. L., Van Der Kouwe, A. J. W., & Fischl, B. (2015). Head motion during MRI acquisition reduces gray matter volume and thickness estimates. NeuroImage, 107, 107–115. 10.1016/j.neuroimage.2014.12.006

56. Rupprecht, T., Kuth, R., Bowing, B., Gerling, S., Wagner, M., & Rascher, W. (2000). Sedation and monitoring of paediatric patients undergoing open low-field MRI. Acta Paediatrica (Oslo, Norway: 1992), 89(9), 1077–1081. 10.1080/713794566

57. Rusche, T., Vosshenrich, J., Winkel, D. J., Donners, R., Segeroth, M., Bach, M., Merkle, E. M., & Breit, H.-C. (2022). More Space, Less Noise-New-generation Low-Field Magnetic Resonance Imaging Systems Can Improve Patient Comfort: A Prospective 0.55T–1.5T-Scanner Comparison. *Journal of Clinical Medicine*, *11*(22), 6705. 10.3390/jcm11226705

58. Safer, D. J. (2018). Is ADHD Really Increasing in Youth? Journal of Attention Disorders, 22(2), 107–115. 10.1177/1087054715586571

59. Satterthwaite, T. D., Wolf, D. H., Loughead, J., Ruparel, K., Elliott, M. A., Hakonarson, H., Gur, R. C., & Gur, R. E. (2012). Impact of in-scanner head motion on multiple measures of functional connectivity: Relevance for studies of neurodevelopment in youth. NeuroImage, 60(1), 623–632. 10.1016/j.neuroimage.2011.12.063

60. Schneider, M., Mehari, K., & Langhinrichsen-Rohling, J. (2020). What Caregivers Want: Preferences for Behavioral Health Screening Implementation Procedures in Pediatric Primary Care. Journal of Clinical Psychology in Medical Settings. 10.1007/s10880-020-09745-1

61. Schrager, J. D., Patzer, R. E., Kim, J. J., Pitts, S. R., Chokshi, F. H., Phillips, J. S., & Zhang, X. (2019). Racial and Ethnic Differences in Diagnostic Imaging Utilization During Adult Emergency Department Visits in the United States, 2005 to 2014. Journal of the American College of Radiology: JACR, 16(8), 1036–1045. 10.1016/j.jacr.2019.03.002

62. Shen, F. X., Wolf, S. M., Bhavnani, S., Deoni, S., Elison, J. T., Fair, D., Garwood, M., Gee, M. S., Geethanath, S., Kay, K., Lim, K. O., Lockwood Estrin, G., Luciana, M., Peloquin, D., Rommelfanger, K., Schiess, N., Siddiqui, K., Torres, E., & Vaughan, J. T. (2021). Emerging ethical issues raised by highly portable MRI research in remote and resource-limited international settings. NeuroImage, 238, 118210. 10.1016/j.neuroimage.2021.118210

63. Sheth, K. N., Mazurek, M. H., Yuen, M. M., Cahn, B. A., Shah, J. T., Ward, A., Kim, J. A., Gilmore, E. J., Falcone, G. J., Petersen, N., Gobeske, K. T., Kaddouh, F., Hwang, D. Y., Schindler, J., Sansing, L., Matouk, C., Rothberg, J., Sze, G., Siner, J., … Kimberly, W. T. (2020). Assessment of Brain Injury Using Portable, Low-Field Magnetic Resonance Imaging at the Bedside of Critically Ill Patients. JAMA Neurology. 10.1001/jamaneurol.2020.3263

64. Sui, Y., Afacan, O., Gholipour, A., & Warfield, S. K. (2021). Fast and High-Resolution Neonatal Brain MRI Through Super-Resolution Reconstruction From Acquisitions With Variable Slice Selection Direction. Frontiers in Neuroscience, 15, 636268. 10.3389/fnins.2021.636268

65. Sydnor, V. J., Larsen, B., Seidlitz, J., Adebimpe, A., Alexander-Bloch, A. F., Bassett, D. S., Bertolero, M. A., Cieslak, M., Covitz, S., Fan, Y., Gur, R. E., Gur, R. C., Mackey, A. P., Moore, T. M., Roalf, D. R., Shinohara, R. T., & Satterthwaite, T. D. (2023). Intrinsic activity development unfolds along a sensorimotor–association cortical axis in youth. Nature Neuroscience, 26(4), Article 4. 10.1038/s41593-023-01282-y

66. Tamnes, C. K., Ostby, Y., Fjell, A. M., Westlye, L. T., Due-Tønnessen, P., & Walhovd, K. B. (2010). Brain maturation in adolescence and young adulthood: Regional age-related changes in cortical thickness and white matter volume and microstructure. *Cerebral Cortex (New York*, N.Y.: 1991), *20*(3), 534–548. 10.1093/cercor/bhp118

67. Thaler, C., Sedlacik, J., Forkert, N. D., Stellmann, J.-P., Schön, G., Fiehler, J., & Gellißen, S. (2023). Effect of geometric distortion correction on thickness and volume measurements of cortical parcellations in 3D T1w gradient echo sequences. PLOS ONE, 18(4), e0284440. 10.1371/journal.pone.0284440

68. Tu, D., Goyal, M. S., Dworkin, J. D., Kampondeni, S., Vidal, L., Biondo-Savin, E., Juvvadi, S., Raghavan, P., Nicholas, J., Chetcuti, K., Clark, K., Robert-Fitzgerald, T., Satterthwaite, T. D., Yushkevich, P., Davatzikos, C., Erus, G., Tustison, N. J., Postels, D. G., Taylor, T. E., … Shinohara, R. T. (2023). Automated analysis of low-field brain MRI in cerebral malaria. Biometrics, 79(3), 2417–2429. 10.1111/biom.13708

69. Vijayakumar, N., Allen, N. B., Youssef, G., Dennison, M., Yücel, M., Simmons, J. G., & Whittle, S. (2016). Brain development during adolescence: A mixed-longitudinal investigation of cortical thickness, surface area, and volume. Human Brain Mapping, 37(6), 2027–2038. 10.1002/hbm.23154

70. Wierenga, L. M., Langen, M., Oranje, B., & Durston, S. (2014). Unique developmental trajectories of cortical thickness and surface area. NeuroImage, 87, 120–126. 10.1016/j.neuroimage.2013.11.010

71. Winkler, A. M., Kochunov, P., Blangero, J., Almasy, L., Zilles, K., Fox, P. T., Duggirala, R., & Glahn, D. C. (2010). Cortical thickness or grey matter volume? The importance of selecting the phenotype for imaging genetics studies. NeuroImage, 53(3), 1135–1146. 10.1016/j.neuroimage.2009.12.028

72. Worthington, M. A., Cao, H., & Cannon, T. D. (2020). Discovery and Validation of Prediction Algorithms for Psychosis in Youths at Clinical High Risk. Biological Psychiatry. Cognitive Neuroscience and Neuroimaging, 5(8), 738–747. 10.1016/j.bpsc.2019.10.006

73. Yao, N., Winkler, A. M., Barrett, J., Book, G. A., Beetham, T., Horseman, R., Leach, O., Hodgson, K., Knowles, E. E., Mathias, S., Stevens, M. C., Assaf, M., van Erp, T. G. M., Pearlson, G. D., & Glahn, D. C. (2017). Inferring pathobiology from structural MRI in schizophrenia and bipolar disorder: Modeling head motion and neuroanatomical specificity. Human Brain Mapping, 38(8), 3757–3770. 10.1002/hbm.23612

74. Yuen, M. M., Prabhat, A. M., Mazurek, M. H., Chavva, I. R., Crawford, A., Cahn, B. A., Beekman, R., Kim, J. A., Gobeske, K. T., Petersen, N. H., Falcone, G. J., Gilmore, E. J., Hwang, D. Y., Jasne, A. S., Amin, H., Sharma, R., Matouk, C., Ward, A., Schindler, J., … Sheth, K. N. (2022). Portable, low-field magnetic resonance imaging enables highly accessible and dynamic bedside evaluation of ischemic stroke. Science Advances, 8(16), eabm3952. 10.1126/sciadv.abm3952

