## Supplement for "Bridging the gap: improving correspondence between low-field and high-field magnetic resonance images in young people"

### Supplementary Content

#### Supplemental Tables

**STable 1.** Scan Parameters for the 64 mT and 3T MRI scans obtained in this study
**STable 2.** Pearson correlations of regional measurements from standard versus SynthSR-processed axial 64mT scans with 3T scans
**STable 3.** Intra-class correlations of regional measurements from standard versus SynthSR-processed axial 64mT scans with 3T scans
**STable 4.** Pearson correlations of regional measurements from standard axial versus standard multi-orientation 64mT scans with 3T scans
**STable 5.** Intra-class correlations of regional measurements from standard versus SynthSR-processed axial 64mT scans with 3T scans
**STable 6.** Correlations of global measurements from SynthSR-processed axial 64mT scans versus SynthSR-processed repeated 64mT scans with 3T scans
**STable 7.** Correlations of individual-level differences between low- and high-field scans with motion during low-field scans (framewise displacement).
**STable 8.** Correlations of individual-level differences between low- and high-field scans with participant age.

#### Supplemental Figures

**SFigure 1.** Flow chart of participant inclusion and exclusion
**SFigure 2.** Bland-Altman plots of individual-level global measurements from standard 64mT axial and 3T scans
**SFigure 3.** Bland-Altman plots of individual-level global measurements from SynthSR-processed 64mT axial and 3T scans
**SFigure 4.** Comparison of individual-level global measurements across standard 64mT axial scans, standard 64mT repeated multi-orientation scans, and traditional 3T scans
**SFigure 5.** Comparison of individual-level global measurements across standard 64mT axial scans, SynthSR-processed 64mT repeated multi-orientation scans, and traditional 3T scans
**SFigure 6.** Comparison of super-resolution approaches in improving correspondence between low-field and high-field-acquired MR images

***Supplemental Tables***

**STable 1.** Scan Parameters for the 64 mT and 3T MRI scans obtained in this study.

| Scan Type | Scanner Manufacturer | Scanner Model | Repetition Time (s) | Echo Time (s) | Inversion Time (s) | Flip Angle (deg) | Percent Phase FOV | Slice Thickness (mm) | Slice Resolution (mm) | Total Acquisition Time (s) |
|---|---|---|---|---|---|---|---|---|---|---|
| T1 64mT Axial | Hyperfine | Swoop | 1.5 | 0.00596 | 0.3 | 90 | 100.00 | 5.0 | 1.6 x 1.6 | 339 |
| T1 64mT Coronal | Hyperfine | Swoop | 1.5 | 0.00559 | 0.3 | 90 | 100.00 | 5.0 | 1.6 x 1.6 | 332 |
| T1 64mT Sagittal | Hyperfine | Swoop | 1.5 | 0.00588 | 0.3 | 90 | 100.00 | 5.0 | 1.6 x 1.6 | 339 |
| T1 3T | Siemens | Prisma | 2.5 | 0.00207 | 1 | 8 | 93.75 | 0.8 | 0.8 x 0.8 | 394 |
| T2 64mT Axial | Hyperfine | Swoop | 2.0 | 0.18240 | • | 90 | 100.00 | 5.0 | 1.6 x 1.6 | 173 |
| T2 64mT Coronal | Hyperfine | Swoop | 2.0 | 0.21760 | • | 90 | 65.00 | 5.0 | 1.6 x 1.6 | 141 |
| T2 64mT Sagittal | Hyperfine | Swoop | 2.0 | 0.23000 | • | 90 | 65.00 | 5.0 | 1.6 x 1.6 | 119 |
| T2 3T | Siemens | Prisma | 3.2 | 0.56400 | • | 120 | 93.75 | 0.8 | 0.8 x 0.8 | 357 |

*Note: Four participants received slightly modified T1-weighted 64mT scans (repetition time = 0.88 s, inversion time = 0.354 s). Excluding these participants from the analyses did not significantly change the results.*

**STable 2.** Pearson correlations of regional measurements from standard versus SynthSR-processed axial 64mT scans with 3T scans. Differences between correlation strengths were tested using Steiger's Z. A positive Z-value indicates that SynthSR-processed regions were more strongly correlated to 3T scans than standard regions. Analyses that are statistically significant after correction for multiple comparisons are in bold.

| Measurement | Standard Axial 64mT Correlations with 3T | | | SynthSR−Processed Axial 64mT Correlations with 3T | | | Steiger | | |
|---|---|---|---|---|---|---|---|---|---|
| | *r* | *p* | *q* | *r* | *p* | *q* | *z* | *p* | *q* |
| left banks of superior temporal sulcus thickness | **-0.32** | **0.007** | **0.015** | -0.07 | 0.589 | 0.675 | 1.81 | 0.070 | 0.113 |
| left caudal anterior cingulate thickness | 0.01 | 0.914 | 0.939 | 0.25 | 0.037 | 0.064 | 1.59 | 0.113 | 0.170 |
| left caudal middle frontal thickness | 0.20 | 0.090 | 0.140 | **0.35** | **0.003** | **0.007** | 1.08 | 0.278 | 0.367 |
| left cuneus thickness | 0.13 | 0.276 | 0.365 | 0.14 | 0.260 | 0.348 | 0.03 | 0.977 | 0.984 |
| left entorhinal thickness | -0.02 | 0.896 | 0.926 | 5.0e-03 | 0.967 | 0.977 | 0.13 | 0.898 | 0.928 |
| left fusiform thickness | -0.17 | 0.168 | 0.239 | 0.17 | 0.166 | 0.237 | 1.95 | 0.051 | 0.086 |
| left inferior parietal thickness | -0.19 | 0.123 | 0.183 | 0.16 | 0.173 | 0.245 | **2.70** | **0.007** | **0.014** |
| left inferior temporal thickness | 0.09 | 0.481 | 0.574 | 0.18 | 0.133 | 0.195 | 0.56 | 0.578 | 0.666 |
| left isthmus cingulate thickness | -0.03 | 0.829 | 0.878 | 0.08 | 0.536 | 0.628 | 0.55 | 0.580 | 0.667 |
| left lateral occipital thickness | 0.01 | 0.929 | 0.949 | 0.07 | 0.568 | 0.657 | 0.39 | 0.700 | 0.768 |
| left lateral orbitofrontal thickness | -0.04 | 0.771 | 0.830 | 0.19 | 0.110 | 0.166 | 1.56 | 0.118 | 0.177 |
| left lingual thickness | 0.09 | 0.449 | 0.542 | -0.06 | 0.614 | 0.695 | -0.83 | 0.405 | 0.503 |
| left medial orbitofrontal thickness | **-0.34** | **0.004** | **0.009** | 0.18 | 0.130 | 0.192 | **2.85** | **0.004** | **0.010** |
| left middle temporal thickness | **-0.27** | **0.022** | **0.041** | **0.30** | **0.013** | **0.025** | **3.24** | **0.001** | **0.003** |
| left parahippocampal thickness | 0.03 | 0.776 | 0.836 | **0.36** | **0.002** | **0.005** | 2.12 | 0.034 | 0.060 |
| left paracentral thickness | 0.06 | 0.642 | 0.720 | 0.04 | 0.761 | 0.821 | -0.12 | 0.903 | 0.930 |
| left pars triangularis thickness | **0.33** | **0.005** | **0.011** | **0.33** | **0.005** | **0.010** | 0.02 | 0.982 | 0.987 |
| left pars opercularis thickness | -0.13 | 0.298 | 0.389 | -0.02 | 0.844 | 0.888 | 0.74 | 0.460 | 0.554 |
| left pars orbitalis thickness | -0.21 | 0.081 | 0.129 | -2.9e-05 | 1.000 | 1.000 | 1.32 | 0.185 | 0.259 |
| left pericalcarine thickness | 0.09 | 0.447 | 0.542 | -0.20 | 0.089 | 0.138 | -1.90 | 0.058 | 0.095 |
| left postcentral thickness | 0.13 | 0.299 | 0.390 | 0.24 | 0.045 | 0.077 | 0.77 | 0.442 | 0.536 |
| left posterior cingulate thickness | -0.18 | 0.133 | 0.196 | 0.24 | 0.045 | 0.077 | **2.46** | **0.014** | **0.027** |
| left precentral thickness | 0.09 | 0.484 | 0.577 | 0.19 | 0.121 | 0.180 | 0.71 | 0.479 | 0.572 |
| left precuneus thickness | 0.07 | 0.550 | 0.642 | **0.40** | **5.74e-04** | **0.001** | **2.30** | **0.022** | **0.040** |
| left rostral anterior cingulate thickness | **-0.30** | **0.010** | **0.021** | -0.02 | 0.891 | 0.921 | 2.04 | 0.042 | 0.072 |
| left rostral middle frontal thickness | -0.01 | 0.915 | 0.939 | 0.09 | 0.467 | 0.561 | 0.81 | 0.419 | 0.515 |
| left superior frontal thickness | 0.02 | 0.843 | 0.888 | **0.36** | **0.002** | **0.005** | **2.78** | **0.005** | **0.012** |
| left superior parietal thickness | 0.04 | 0.722 | 0.787 | 0.11 | 0.363 | 0.461 | 0.43 | 0.670 | 0.742 |
| left superior temporal thickness | -0.24 | 0.049 | 0.082 | **0.31** | **0.009** | **0.019** | **3.10** | **0.002** | **0.005** |

| | | | | | | | | | |
|---|---|---|---|---|---|---|---|---|---|
| left supramarginal thickness | 0.02 | 0.859 | 0.898 | **0.41** | **4.39e-04** | **0.001** | **2.44** | **0.015** | **0.028** |
| left frontal pole thickness | -0.23 | 0.058 | 0.095 | **-0.35** | **0.003** | **0.007** | -0.75 | 0.453 | 0.547 |
| left temporal pole thickness | -0.24 | 0.048 | 0.081 | 0.14 | 0.257 | 0.345 | 2.18 | 0.029 | 0.052 |
| left transverse temporal thickness | **-0.30** | **0.012** | **0.023** | **0.27** | **0.024** | **0.043** | **3.46** | **5.49e-04** | **0.001** |
| left insula thickness | -0.07 | 0.539 | 0.631 | -0.10 | 0.406 | 0.503 | -0.18 | 0.861 | 0.899 |
| right banks of superior temporal sulcus thickness | -0.13 | 0.290 | 0.381 | 0.10 | 0.392 | 0.491 | 1.71 | 0.087 | 0.136 |
| right caudal anterior cingulate thickness | -0.16 | 0.174 | 0.247 | 0.01 | 0.931 | 0.949 | 0.97 | 0.333 | 0.427 |
| right caudal middle frontal thickness | 0.05 | 0.656 | 0.732 | **0.29** | **0.015** | **0.029** | 1.46 | 0.145 | 0.210 |
| right cuneus thickness | 0.10 | 0.428 | 0.523 | 0.14 | 0.231 | 0.315 | 0.28 | 0.780 | 0.839 |
| right entorhinal thickness | -0.23 | 0.057 | 0.094 | -0.05 | 0.661 | 0.736 | 1.01 | 0.313 | 0.405 |
| right fusiform thickness | -0.21 | 0.088 | 0.137 | **-0.35** | **0.003** | **0.007** | -0.93 | 0.350 | 0.447 |
| right inferior parietal thickness | -0.21 | 0.077 | 0.123 | 0.06 | 0.649 | 0.725 | 2.02 | 0.043 | 0.074 |
| right inferior temporal thickness | -0.14 | 0.256 | 0.344 | -2.7e-03 | 0.982 | 0.987 | 0.88 | 0.376 | 0.475 |
| right isthmus cingulate thickness | -0.02 | 0.871 | 0.907 | 0.06 | 0.608 | 0.690 | 0.49 | 0.626 | 0.706 |
| right lateral occipital thickness | 0.18 | 0.128 | 0.189 | 0.06 | 0.632 | 0.712 | -1.02 | 0.309 | 0.400 |
| right lateral orbitofrontal thickness | -0.05 | 0.710 | 0.776 | 0.09 | 0.474 | 0.568 | 0.77 | 0.441 | 0.536 |
| right lingual thickness | -0.04 | 0.756 | 0.817 | 0.06 | 0.630 | 0.710 | 0.56 | 0.574 | 0.662 |
| right medial orbitofrontal thickness | **-0.35** | **0.003** | **0.006** | -0.05 | 0.686 | 0.756 | 2.07 | 0.039 | 0.068 |
| right middle temporal thickness | **-0.37** | **0.002** | **0.004** | 0.17 | 0.164 | 0.234 | **3.37** | **7.46e-04** | **0.002** |
| right parahippocampal thickness | 0.21 | 0.077 | 0.122 | **0.40** | **6.56e-04** | **0.002** | 1.13 | 0.259 | 0.347 |
| right paracentral thickness | 0.17 | 0.171 | 0.243 | 0.09 | 0.459 | 0.553 | -0.51 | 0.608 | 0.690 |
| right pars triangularis thickness | 0.21 | 0.087 | 0.136 | 0.20 | 0.105 | 0.159 | -0.08 | 0.938 | 0.955 |
| right pars opercularis thickness | -0.05 | 0.683 | 0.754 | **0.28** | **0.021** | **0.039** | 2.01 | 0.045 | 0.076 |
| right pars orbitalis thickness | 3.2e-03 | 0.979 | 0.986 | 0.25 | 0.036 | 0.063 | 1.83 | 0.067 | 0.108 |
| right pericalcarine thickness | -0.03 | 0.808 | 0.861 | -7.4e-03 | 0.951 | 0.965 | 0.15 | 0.883 | 0.916 |
| right postcentral thickness | 0.15 | 0.229 | 0.313 | **0.27** | **0.026** | **0.047** | 0.90 | 0.366 | 0.463 |
| right posterior cingulate thickness | -0.11 | 0.362 | 0.460 | 0.24 | 0.043 | 0.074 | **2.39** | **0.017** | **0.032** |
| right precentral thickness | 0.16 | 0.187 | 0.261 | -0.06 | 0.619 | 0.699 | -1.71 | 0.087 | 0.136 |
| right precuneus thickness | 0.07 | 0.539 | 0.631 | 0.20 | 0.093 | 0.143 | 0.91 | 0.361 | 0.459 |
| right rostral anterior cingulate thickness | **-0.37** | **0.001** | **0.003** | -0.13 | 0.278 | 0.367 | 1.60 | 0.111 | 0.167 |
| right rostral middle frontal thickness | -0.19 | 0.113 | 0.170 | 0.03 | 0.825 | 0.874 | 1.46 | 0.145 | 0.211 |
| right superior frontal thickness | 0.16 | 0.193 | 0.268 | 0.11 | 0.374 | 0.473 | -0.36 | 0.722 | 0.787 |
| right superior parietal thickness | 0.12 | 0.314 | 0.405 | 0.08 | 0.525 | 0.618 | -0.34 | 0.735 | 0.800 |
| right superior temporal thickness | -0.18 | 0.144 | 0.209 | **0.43** | **2.45e-04** | **6.69e-04** | **4.08** | **4.42e-05** | **1.35e-04** |
| right supramarginal thickness | 0.19 | 0.106 | 0.161 | **0.39** | **7.92e-04** | **0.002** | 1.58 | 0.114 | 0.172 |

| | | | | | | | | | |
|---|---|---|---|---|---|---|---|---|---|
| right frontal pole thickness | -0.10 | 0.406 | 0.503 | -0.14 | 0.263 | 0.351 | -0.21 | 0.836 | 0.882 |
| right temporal pole thickness | 0.17 | 0.168 | 0.239 | 0.22 | 0.062 | 0.102 | 0.37 | 0.711 | 0.776 |
| right transverse temporal thickness | -0.03 | 0.814 | 0.866 | 0.06 | 0.594 | 0.679 | 0.59 | 0.554 | 0.646 |
| right insula thickness | -0.07 | 0.589 | 0.674 | 0.17 | 0.170 | 0.241 | 1.35 | 0.178 | 0.251 |
| left banks of superior temporal sulcus area | **0.34** | **0.003** | **0.008** | **0.73** | **7.58e-13** | **6.82e-12** | **3.95** | **7.77e-05** | **2.28e-04** |
| left caudal anterior cingulate area | **0.35** | **0.003** | **0.007** | **0.48** | **2.46e-05** | **7.81e-05** | 1.62 | 0.106 | 0.161 |
| left caudal middle frontal area | **0.31** | **0.009** | **0.018** | **0.73** | **5.25e-13** | **4.80e-12** | **4.12** | **3.78e-05** | **1.17e-04** |
| left cuneus area | **0.41** | **4.66e-04** | **0.001** | **0.64** | **1.98e-09** | **1.14e-08** | 1.96 | 0.050 | 0.084 |
| left entorhinal area | -0.06 | 0.607 | 0.690 | **0.41** | **4.01e-04** | **0.001** | **2.69** | **0.007** | **0.015** |
| left fusiform area | **0.31** | **0.009** | **0.018** | **0.65** | **1.61e-09** | **9.45e-09** | **2.54** | **0.011** | **0.022** |
| left inferior parietal area | **0.59** | **1.04e-07** | **4.85e-07** | **0.84** | **1.27e-19** | **3.75e-18** | **3.63** | **2.79e-04** | **7.52e-04** |
| left inferior temporal area | **0.43** | **1.98e-04** | **5.47e-04** | **0.74** | **2.58e-13** | **2.48e-12** | **3.07** | **0.002** | **0.005** |
| left isthmus cingulate area | **0.34** | **0.004** | **0.009** | **0.51** | **5.65e-06** | **2.02e-05** | 1.28 | 0.202 | 0.279 |
| left lateral occipital area | **0.33** | **0.005** | **0.011** | **0.52** | **4.68e-06** | **1.70e-05** | 1.38 | 0.167 | 0.237 |
| left lateral orbitofrontal area | **0.51** | **6.58e-06** | **2.33e-05** | **0.67** | **1.51e-10** | **1.01e-09** | 1.82 | 0.068 | 0.111 |
| left lingual area | 0.24 | 0.046 | 0.078 | **0.59** | **9.68e-08** | **4.53e-07** | **2.52** | **0.012** | **0.023** |
| left medial orbitofrontal area | **0.49** | **1.68e-05** | **5.53e-05** | **0.51** | **6.85e-06** | **2.41e-05** | 0.19 | 0.853 | 0.895 |
| left middle temporal area | **0.46** | **6.52e-05** | **1.93e-04** | **0.75** | **1.03e-13** | **1.04e-12** | **3.09** | **0.002** | **0.005** |
| left parahippocampal area | -0.23 | 0.060 | 0.098 | 0.24 | 0.048 | 0.081 | **2.59** | **0.009** | **0.019** |
| left paracentral area | **0.34** | **0.003** | **0.008** | **0.76** | **1.59e-14** | **1.77e-13** | **3.95** | **7.94e-05** | **2.32e-04** |
| left pars triangularis area | **0.40** | **6.91e-04** | **0.002** | **0.65** | **1.11e-09** | **6.68e-09** | **2.55** | **0.011** | **0.022** |
| left pars opercularis area | **0.45** | **8.10e-05** | **2.37e-04** | **0.49** | **1.66e-05** | **5.46e-05** | 0.31 | 0.760 | 0.820 |
| left pars orbitalis area | **0.60** | **4.50e-08** | **2.22e-07** | **0.63** | **4.27e-09** | **2.37e-08** | 0.38 | 0.704 | 0.771 |
| left pericalcarine area | **0.33** | **0.005** | **0.011** | **0.65** | **1.12e-09** | **6.73e-09** | **2.81** | **0.005** | **0.011** |
| left postcentral area | 0.21 | 0.088 | 0.137 | **0.70** | **1.57e-11** | **1.19e-10** | **4.25** | **2.12e-05** | **6.80e-05** |
| left posterior cingulate area | **0.30** | **0.011** | **0.022** | **0.57** | **2.02e-07** | **9.01e-07** | **2.45** | **0.014** | **0.027** |
| left precentral area | **0.36** | **0.002** | **0.006** | **0.83** | **3.20e-19** | **8.40e-18** | **5.70** | **1.22e-08** | **6.43e-08** |
| left precuneus area | **0.32** | **0.007** | **0.015** | **0.72** | **1.47e-12** | **1.29e-11** | **3.55** | **3.88e-04** | **0.001** |
| left rostral anterior cingulate area | **0.44** | **1.35e-04** | **3.84e-04** | **0.60** | **2.96e-08** | **1.48e-07** | 1.97 | 0.048 | 0.082 |
| left rostral middle frontal area | **0.61** | **2.61e-08** | **1.31e-07** | **0.73** | **5.50e-13** | **4.99e-12** | 2.09 | 0.037 | 0.064 |
| left superior frontal area | **0.53** | **1.85e-06** | **7.08e-06** | **0.77** | **3.45e-15** | **4.30e-14** | **3.46** | **5.38e-04** | **0.001** |
| left superior parietal area | **0.37** | **0.002** | **0.004** | **0.67** | **2.72e-10** | **1.77e-09** | **2.60** | **0.009** | **0.019** |
| left superior temporal area | **0.29** | **0.015** | **0.029** | **0.79** | **3.02e-16** | **4.64e-15** | **5.45** | **5.08e-08** | **2.47e-07** |
| left supramarginal area | **0.52** | **3.70e-06** | **1.36e-05** | **0.78** | **1.67e-15** | **2.27e-14** | **3.16** | **0.002** | **0.004** |

| | | | | | | | | | |
|---|---|---|---|---|---|---|---|---|---|
| left frontal pole area | 0.25 | 0.038 | 0.066 | **0.27** | **0.023** | **0.043** | 0.14 | 0.886 | 0.918 |
| left temporal pole area | -0.10 | 0.428 | 0.523 | 0.14 | 0.253 | 0.341 | 1.34 | 0.179 | 0.252 |
| left transverse temporal area | 0.16 | 0.192 | 0.267 | **0.73** | **4.45e-13** | **4.13e-12** | **4.64** | **3.56e-06** | **1.31e-05** |
| left insula area | **0.28** | **0.018** | **0.033** | **0.77** | **8.40e-15** | **9.89e-14** | **4.57** | **4.94e-06** | **1.78e-05** |
| right banks of superior temporal sulcus area | **0.41** | **3.90e-04** | **0.001** | **0.69** | **5.38e-11** | **3.70e-10** | **2.60** | **0.009** | **0.019** |
| right caudal anterior cingulate area | **0.62** | **9.78e-09** | **5.24e-08** | **0.71** | **6.36e-12** | **5.04e-11** | 1.32 | 0.186 | 0.260 |
| right caudal middle frontal area | **0.44** | **1.14e-04** | **3.28e-04** | **0.60** | **4.47e-08** | **2.21e-07** | 1.59 | 0.112 | 0.169 |
| right cuneus area | 0.18 | 0.141 | 0.206 | **0.51** | **5.56e-06** | **1.99e-05** | **2.81** | **0.005** | **0.011** |
| right entorhinal area | -0.16 | 0.183 | 0.257 | 0.15 | 0.210 | 0.288 | 1.98 | 0.048 | 0.081 |
| right fusiform area | **0.44** | **1.17e-04** | **3.36e-04** | **0.77** | **3.84e-15** | **4.76e-14** | **3.48** | **4.96e-04** | **0.001** |
| right inferior parietal area | **0.53** | **2.26e-06** | **8.58e-06** | **0.74** | **3.22e-13** | **3.06e-12** | **2.57** | **0.010** | **0.020** |
| right inferior temporal area | **0.55** | **8.35e-07** | **3.39e-06** | **0.77** | **5.89e-15** | **7.05e-14** | **2.76** | **0.006** | **0.012** |
| right isthmus cingulate area | 0.19 | 0.122 | 0.181 | **0.72** | **1.59e-12** | **1.38e-11** | **4.74** | **2.09e-06** | **7.97e-06** |
| right lateral occipital area | **0.54** | **1.44e-06** | **5.62e-06** | **0.65** | **1.26e-09** | **7.49e-09** | 1.13 | 0.257 | 0.344 |
| right lateral orbitofrontal area | **0.51** | **7.31e-06** | **2.56e-05** | **0.54** | **1.19e-06** | **4.72e-06** | 0.35 | 0.725 | 0.790 |
| right lingual area | 0.10 | 0.409 | 0.505 | **0.60** | **5.27e-08** | **2.56e-07** | **4.33** | **1.46e-05** | **4.86e-05** |
| right medial orbitofrontal area | **0.43** | **2.03e-04** | **5.58e-04** | **0.45** | **8.81e-05** | **2.57e-04** | 0.17 | 0.868 | 0.904 |
| right middle temporal area | **0.45** | **9.29e-05** | **2.69e-04** | **0.76** | **3.22e-14** | **3.45e-13** | **3.59** | **3.34e-04** | **8.93e-04** |
| right parahippocampal area | 0.03 | 0.814 | 0.866 | **0.28** | **0.018** | **0.035** | 1.53 | 0.126 | 0.187 |
| right paracentral area | **0.36** | **0.002** | **0.005** | **0.62** | **1.45e-08** | **7.60e-08** | **2.44** | **0.015** | **0.028** |
| right pars triangularis area | **0.42** | **3.09e-04** | **8.29e-04** | **0.75** | **6.47e-14** | **6.74e-13** | **3.97** | **7.26e-05** | **2.14e-04** |
| right pars opercularis area | **0.48** | **2.23e-05** | **7.13e-05** | **0.55** | **1.03e-06** | **4.11e-06** | 0.58 | 0.561 | 0.651 |
| right pars orbitalis area | **0.37** | **0.001** | **0.003** | **0.64** | **2.70e-09** | **1.52e-08** | **2.75** | **0.006** | **0.013** |
| right pericalcarine area | **0.44** | **1.53e-04** | **4.31e-04** | **0.68** | **8.42e-11** | **5.69e-10** | **2.63** | **0.009** | **0.017** |
| right postcentral area | **0.44** | **1.27e-04** | **3.64e-04** | **0.69** | **3.44e-11** | **2.43e-10** | **2.84** | **0.004** | **0.010** |
| right posterior cingulate area | **0.32** | **0.007** | **0.014** | **0.70** | **2.11e-11** | **1.57e-10** | **4.18** | **2.88e-05** | **9.02e-05** |
| right precentral area | **0.48** | **2.62e-05** | **8.27e-05** | **0.63** | **6.76e-09** | **3.69e-08** | 1.51 | 0.132 | 0.193 |
| right precuneus area | **0.43** | **1.77e-04** | **4.92e-04** | **0.70** | **1.61e-11** | **1.21e-10** | **2.58** | **0.010** | **0.020** |
| right rostral anterior cingulate area | **0.57** | **2.66e-07** | **1.17e-06** | **0.78** | **1.14e-15** | **1.57e-14** | **3.20** | **0.001** | **0.003** |
| right rostral middle frontal area | **0.49** | **1.97e-05** | **6.38e-05** | **0.64** | **1.86e-09** | **1.07e-08** | 1.78 | 0.075 | 0.120 |
| right superior frontal area | **0.56** | **4.54e-07** | **1.92e-06** | **0.69** | **3.90e-11** | **2.74e-10** | 1.82 | 0.069 | 0.111 |
| right superior parietal area | 0.22 | 0.062 | 0.102 | **0.61** | **1.72e-08** | **8.90e-08** | **3.21** | **0.001** | **0.003** |
| right superior temporal area | **0.50** | **8.61e-06** | **2.98e-05** | **0.79** | **2.42e-16** | **3.83e-15** | **3.72** | **1.99e-04** | **5.48e-04** |
| right supramarginal area | **0.28** | **0.021** | **0.039** | **0.66** | **5.42e-10** | **3.39e-09** | **3.66** | **2.49e-04** | **6.80e-04** |
| right frontal pole area | **0.26** | **0.027** | **0.049** | 0.14 | 0.252 | 0.340 | -0.85 | 0.396 | 0.495 |

| | | | | | | | | | |
|---|---|---|---|---|---|---|---|---|---|
| right temporal pole area | 7.4e-03 | 0.951 | 0.965 | 0.07 | 0.578 | 0.666 | 0.36 | 0.717 | 0.783 |
| right transverse temporal area | 0.26 | 0.032 | 0.057 | **0.59** | **6.46e-08** | **3.08e-07** | **2.71** | **0.007** | **0.014** |
| right insula area | 0.22 | 0.066 | 0.107 | **0.67** | **2.15e-10** | **1.41e-09** | **3.55** | **3.84e-04** | **0.001** |
| left banks of superior temporal sulcus volume | 0.26 | 0.033 | 0.059 | **0.67** | **2.73e-10** | **1.77e-09** | **4.31** | **1.60e-05** | **5.28e-05** |
| left caudal anterior cingulate volume | **0.30** | **0.011** | **0.021** | **0.58** | **1.43e-07** | **6.51e-07** | **3.48** | **5.03e-04** | **0.001** |
| left caudal middle frontal volume | **0.40** | **6.72e-04** | **0.002** | **0.73** | **1.24e-12** | **1.08e-11** | **3.29** | **9.99e-04** | **0.002** |
| left cuneus volume | **0.31** | **0.010** | **0.020** | **0.66** | **6.62e-10** | **4.10e-09** | **2.90** | **0.004** | **0.008** |
| left entorhinal volume | -0.10 | 0.405 | 0.503 | **0.39** | **9.33e-04** | **0.002** | **2.74** | **0.006** | **0.013** |
| left fusiform volume | **0.31** | **0.009** | **0.018** | **0.66** | **5.61e-10** | **3.50e-09** | **2.71** | **0.007** | **0.014** |
| left inferior parietal volume | **0.57** | **3.03e-07** | **1.31e-06** | **0.85** | **7.18e-21** | **2.95e-19** | **4.18** | **2.89e-05** | **9.05e-05** |
| left inferior temporal volume | **0.46** | **6.67e-05** | **1.97e-04** | **0.78** | **2.76e-15** | **3.49e-14** | **3.50** | **4.70e-04** | **0.001** |
| left isthmus cingulate volume | **0.31** | **0.010** | **0.020** | **0.52** | **4.48e-06** | **1.64e-05** | 1.52 | 0.130 | 0.191 |
| left lateral occipital volume | **0.39** | **7.49e-04** | **0.002** | **0.55** | **9.78e-07** | **3.94e-06** | 1.31 | 0.189 | 0.263 |
| left lateral orbitofrontal volume | **0.48** | **2.98e-05** | **9.31e-05** | **0.69** | **5.83e-11** | **3.96e-10** | 2.18 | 0.029 | 0.052 |
| left lingual volume | 0.12 | 0.312 | 0.403 | **0.65** | **8.48e-10** | **5.14e-09** | **4.19** | **2.84e-05** | **8.93e-05** |
| left medial orbitofrontal volume | **0.30** | **0.012** | **0.024** | **0.41** | **3.75e-04** | **9.91e-04** | 0.94 | 0.348 | 0.445 |
| left middle temporal volume | **0.31** | **0.010** | **0.020** | **0.71** | **6.56e-12** | **5.18e-11** | **3.73** | **1.92e-04** | **5.31e-04** |
| left parahippocampal volume | -0.04 | 0.721 | 0.787 | 0.19 | 0.109 | 0.165 | 1.40 | 0.163 | 0.233 |
| left paracentral volume | **0.32** | **0.006** | **0.013** | **0.65** | **1.22e-09** | **7.26e-09** | **2.75** | **0.006** | **0.013** |
| left pars triangularis volume | **0.47** | **4.51e-05** | **1.37e-04** | **0.65** | **8.33e-10** | **5.07e-09** | **2.26** | **0.024** | **0.044** |
| left pars opercularis volume | **0.35** | **0.003** | **0.006** | **0.43** | **1.92e-04** | **5.31e-04** | 0.63 | 0.528 | 0.620 |
| left pars orbitalis volume | **0.54** | **1.27e-06** | **4.99e-06** | **0.58** | **1.13e-07** | **5.22e-07** | 0.42 | 0.673 | 0.744 |
| left pericalcarine volume | **0.32** | **0.007** | **0.014** | **0.56** | **3.72e-07** | **1.60e-06** | 2.04 | 0.041 | 0.071 |
| left postcentral volume | 0.23 | 0.053 | 0.088 | **0.68** | **7.81e-11** | **5.30e-10** | **3.79** | **1.50e-04** | **4.24e-04** |
| left posterior cingulate volume | **0.27** | **0.023** | **0.042** | **0.55** | **7.17e-07** | **2.94e-06** | **2.49** | **0.013** | **0.025** |
| left precentral volume | **0.36** | **0.002** | **0.006** | **0.79** | **3.11e-16** | **4.74e-15** | **5.42** | **6.11e-08** | **2.93e-07** |
| left precuneus volume | **0.29** | **0.015** | **0.029** | **0.75** | **6.46e-14** | **6.74e-13** | **4.29** | **1.76e-05** | **5.76e-05** |
| left rostral anterior cingulate volume | **0.39** | **8.49e-04** | **0.002** | **0.52** | **4.98e-06** | **1.79e-05** | 1.31 | 0.189 | 0.263 |
| left rostral middle frontal volume | **0.51** | **7.43e-06** | **2.59e-05** | **0.60** | **5.01e-08** | **2.45e-07** | 1.08 | 0.280 | 0.370 |
| left superior frontal volume | **0.38** | **0.001** | **0.003** | **0.70** | **1.22e-11** | **9.42e-11** | **4.15** | **3.36e-05** | **1.04e-04** |
| left superior parietal volume | **0.47** | **4.29e-05** | **1.31e-04** | **0.64** | **1.76e-09** | **1.03e-08** | 1.57 | 0.117 | 0.174 |
| left superior temporal volume | 0.21 | 0.087 | 0.136 | **0.80** | **1.05e-16** | **1.74e-15** | **6.18** | **6.46e-10** | **4.01e-09** |
| left supramarginal volume | **0.42** | **2.70e-04** | **7.31e-04** | **0.75** | **4.94e-14** | **5.22e-13** | **3.66** | **2.51e-04** | **6.85e-04** |
| left frontal pole volume | -0.04 | 0.754 | 0.816 | -0.11 | 0.371 | 0.469 | -0.48 | 0.632 | 0.712 |

| | | | | | | | | | |
|---|---|---|---|---|---|---|---|---|---|
| left temporal pole volume | -0.08 | 0.495 | 0.589 | 0.08 | 0.528 | 0.620 | 0.85 | 0.394 | 0.492 |
| left transverse temporal volume | 0.11 | 0.380 | 0.479 | **0.68** | **8.82e-11** | **5.95e-10** | **4.39** | **1.14e-05** | **3.89e-05** |
| left insula volume | **0.42** | **2.65e-04** | **7.20e-04** | **0.73** | **4.72e-13** | **4.35e-12** | **3.31** | **9.48e-04** | **0.002** |
| right banks of superior temporal sulcus volume | **0.43** | **1.89e-04** | **5.23e-04** | **0.61** | **1.84e-08** | **9.50e-08** | 1.64 | 0.101 | 0.155 |
| right caudal anterior cingulate volume | **0.64** | **2.45e-09** | **1.39e-08** | **0.74** | **4.18e-13** | **3.89e-12** | 1.39 | 0.166 | 0.237 |
| right caudal middle frontal volume | **0.47** | **4.19e-05** | **1.28e-04** | **0.58** | **1.40e-07** | **6.37e-07** | 1.11 | 0.268 | 0.356 |
| right cuneus volume | 0.16 | 0.179 | 0.252 | **0.51** | **7.31e-06** | **2.56e-05** | **2.80** | **0.005** | **0.011** |
| right entorhinal volume | -0.24 | 0.042 | 0.072 | 0.22 | 0.067 | 0.108 | **2.88** | **0.004** | **0.009** |
| right fusiform volume | **0.32** | **0.007** | **0.014** | **0.72** | **1.56e-12** | **1.36e-11** | **3.63** | **2.88e-04** | **7.76e-04** |
| right inferior parietal volume | **0.48** | **2.84e-05** | **8.93e-05** | **0.73** | **8.85e-13** | **7.87e-12** | **3.20** | **0.001** | **0.003** |
| right inferior temporal volume | **0.53** | **2.75e-06** | **1.03e-05** | **0.76** | **1.43e-14** | **1.62e-13** | **2.86** | **0.004** | **0.009** |
| right isthmus cingulate volume | 0.19 | 0.112 | 0.169 | **0.69** | **3.15e-11** | **2.24e-10** | **4.42** | **9.72e-06** | **3.34e-05** |
| right lateral occipital volume | **0.57** | **3.13e-07** | **1.36e-06** | **0.62** | **1.03e-08** | **5.46e-08** | 0.61 | 0.542 | 0.634 |
| right lateral orbitofrontal volume | **0.49** | **1.86e-05** | **6.06e-05** | **0.60** | **4.26e-08** | **2.11e-07** | 1.13 | 0.257 | 0.344 |
| right lingual volume | 0.04 | 0.753 | 0.815 | **0.64** | **1.78e-09** | **1.04e-08** | **5.43** | **5.61e-08** | **2.70e-07** |
| right medial orbitofrontal volume | **0.28** | **0.020** | **0.038** | **0.32** | **0.008** | **0.016** | 0.29 | 0.774 | 0.834 |
| right middle temporal volume | **0.38** | **0.001** | **0.003** | **0.69** | **3.21e-11** | **2.28e-10** | **3.28** | **0.001** | **0.003** |
| right parahippocampal volume | 0.12 | 0.309 | 0.400 | 0.03 | 0.787 | 0.844 | -0.55 | 0.583 | 0.670 |
| right paracentral volume | **0.49** | **1.46e-05** | **4.86e-05** | **0.55** | **8.02e-07** | **3.27e-06** | 0.58 | 0.564 | 0.654 |
| right pars triangularis volume | **0.44** | **1.41e-04** | **3.98e-04** | **0.64** | **2.07e-09** | **1.19e-08** | **2.29** | **0.022** | **0.041** |
| right pars opercularis volume | **0.48** | **3.10e-05** | **9.63e-05** | **0.55** | **7.12e-07** | **2.93e-06** | 0.74 | 0.457 | 0.551 |
| right pars orbitalis volume | **0.38** | **0.001** | **0.003** | **0.59** | **5.76e-08** | **2.77e-07** | **2.33** | **0.020** | **0.037** |
| right pericalcarine volume | **0.32** | **0.006** | **0.013** | **0.61** | **1.63e-08** | **8.45e-08** | **2.73** | **0.006** | **0.013** |
| right postcentral volume | **0.50** | **1.04e-05** | **3.53e-05** | **0.71** | **4.10e-12** | **3.34e-11** | **2.64** | **0.008** | **0.017** |
| right posterior cingulate volume | **0.37** | **0.002** | **0.004** | **0.67** | **2.15e-10** | **1.41e-09** | **3.31** | **9.38e-04** | **0.002** |
| right precentral volume | **0.50** | **1.12e-05** | **3.80e-05** | **0.61** | **1.49e-08** | **7.78e-08** | 1.34 | 0.180 | 0.252 |
| right precuneus volume | **0.31** | **0.008** | **0.017** | **0.70** | **2.44e-11** | **1.77e-10** | **3.60** | **3.18e-04** | **8.53e-04** |
| right rostral anterior cingulate volume | **0.53** | **2.12e-06** | **8.07e-06** | **0.71** | **5.60e-12** | **4.47e-11** | 2.11 | 0.035 | 0.061 |
| right rostral middle frontal volume | **0.31** | **0.008** | **0.016** | **0.54** | **1.69e-06** | **6.49e-06** | **2.54** | **0.011** | **0.022** |
| right superior frontal volume | **0.47** | **3.63e-05** | **1.12e-04** | **0.65** | **1.05e-09** | **6.34e-09** | **2.31** | **0.021** | **0.039** |
| right superior parietal volume | **0.28** | **0.020** | **0.037** | **0.59** | **8.63e-08** | **4.06e-07** | **2.71** | **0.007** | **0.014** |
| right superior temporal volume | **0.53** | **2.03e-06** | **7.76e-06** | **0.79** | **8.11e-16** | **1.13e-14** | **3.42** | **6.23e-04** | **0.002** |
| right supramarginal volume | **0.39** | **9.04e-04** | **0.002** | **0.76** | **1.37e-14** | **1.56e-13** | **4.62** | **3.78e-06** | **1.39e-05** |
| right frontal pole volume | 0.03 | 0.791 | 0.847 | -0.05 | 0.658 | 0.734 | -0.53 | 0.595 | 0.679 |
| right temporal pole volume | 0.11 | 0.365 | 0.463 | 0.08 | 0.511 | 0.604 | -0.18 | 0.861 | 0.899 |

| | | | | | | | | | |
|---|---|---|---|---|---|---|---|---|---|
| right transverse temporal volume | 0.23 | 0.055 | 0.091 | **0.51** | **5.43e-06** | **1.95e-05** | **2.33** | **0.020** | **0.037** |
| right insula volume | 0.23 | 0.052 | 0.087 | **0.72** | **2.38e-12** | **2.00e-11** | **4.06** | **4.91e-05** | **1.49e-04** |
| left lateral ventricle subcortical volume | **0.96** | **9.47e-38** | **2.58e-35** | **0.98** | **9.25e-52** | **5.04e-49** | **4.43** | **9.34e-06** | **3.22e-05** |
| left inf lat vent subcortical volume | **0.61** | **1.48e-08** | **7.71e-08** | **0.57** | **2.08e-07** | **9.23e-07** | -0.40 | 0.686 | 0.756 |
| left cerebellum white matter subcortical volume | **0.50** | **9.03e-06** | **3.12e-05** | **0.39** | **8.08e-04** | **0.002** | -0.86 | 0.391 | 0.491 |
| left cerebellum cortex subcortical volume | **0.74** | **3.75e-13** | **3.53e-12** | **0.88** | **1.06e-23** | **7.41e-22** | **3.44** | **5.75e-04** | **0.001** |
| left thalamus proper subcortical volume | **0.41** | **5.04e-04** | **0.001** | **0.79** | **2.92e-16** | **4.50e-15** | **4.01** | **5.96e-05** | **1.77e-04** |
| left caudate subcortical volume | **0.49** | **1.43e-05** | **4.79e-05** | **0.71** | **5.75e-12** | **4.57e-11** | **2.91** | **0.004** | **0.008** |
| left putamen subcortical volume | **0.38** | **0.001** | **0.003** | **0.57** | **2.89e-07** | **1.26e-06** | 1.94 | 0.052 | 0.087 |
| left pallidum subcortical volume | **0.27** | **0.025** | **0.046** | **0.51** | **6.62e-06** | **2.34e-05** | 1.94 | 0.053 | 0.088 |
| 3rd ventricle subcortical volume | **0.72** | **1.65e-12** | **1.43e-11** | **0.90** | **7.19e-26** | **6.80e-24** | **4.21** | **2.50e-05** | **7.92e-05** |
| 4th ventricle subcortical volume | **0.62** | **7.64e-09** | **4.15e-08** | **0.85** | **1.33e-20** | **5.07e-19** | **3.79** | **1.53e-04** | **4.31e-04** |
| brain stem subcortical volume | **0.79** | **3.29e-16** | **4.97e-15** | **0.90** | **7.33e-27** | **7.60e-25** | **3.34** | **8.32e-04** | **0.002** |
| left hippocampus subcortical volume | 0.25 | 0.038 | 0.067 | **0.51** | **6.57e-06** | **2.33e-05** | 1.90 | 0.058 | 0.095 |
| left amygdala subcortical volume | **0.34** | **0.004** | **0.009** | **0.48** | **2.92e-05** | **9.12e-05** | 1.18 | 0.239 | 0.325 |
| csf subcortical volume | **0.59** | **7.99e-08** | **3.78e-07** | **0.77** | **5.44e-15** | **6.59e-14** | **3.48** | **4.98e-04** | **0.001** |
| left accumbens area subcortical volume | 0.01 | 0.932 | 0.950 | 0.24 | 0.044 | 0.075 | 1.39 | 0.165 | 0.236 |
| left ventral diencephalon subcortical volume | **0.49** | **1.68e-05** | **5.52e-05** | **0.69** | **5.19e-11** | **3.58e-10** | 2.17 | 0.030 | 0.053 |
| left vessel subcortical volume | 0.02 | 0.853 | 0.895 | **0.40** | **6.24e-04** | **0.002** | 2.19 | 0.028 | 0.051 |
| left choroid plexus subcortical volume | **0.52** | **3.10e-06** | **1.16e-05** | **0.74** | **3.07e-13** | **2.93e-12** | **3.43** | **5.96e-04** | **0.002** |
| right lateral ventricle subcortical volume | **0.92** | **3.67e-29** | **6.67e-27** | **0.98** | **3.13e-47** | **1.37e-44** | **5.15** | **2.64e-07** | **1.16e-06** |
| right inf lat vent subcortical volume | **0.57** | **3.16e-07** | **1.37e-06** | **0.56** | **3.57e-07** | **1.54e-06** | -0.02 | 0.986 | 0.990 |
| cerebellum white matter subcortical volume | **0.67** | **1.51e-10** | **1.01e-09** | **0.52** | **3.30e-06** | **1.22e-05** | -1.67 | 0.096 | 0.148 |
| cerebellum cortex subcortical volume | **0.69** | **4.46e-11** | **3.11e-10** | **0.86** | **8.15e-22** | **4.44e-20** | **3.37** | **7.64e-04** | **0.002** |
| right thalamus proper subcortical volume | **0.30** | **0.013** | **0.025** | **0.75** | **1.25e-13** | **1.24e-12** | **4.44** | **8.96e-06** | **3.10e-05** |
| right caudate subcortical volume | 0.25 | 0.040 | 0.070 | **0.48** | **2.28e-05** | **7.27e-05** | **2.47** | **0.013** | **0.026** |
| right putamen subcortical volume | **0.29** | **0.014** | **0.027** | **0.71** | **6.78e-12** | **5.33e-11** | **3.65** | **2.65e-04** | **7.21e-04** |
| right pallidum subcortical volume | 0.24 | 0.043 | 0.074 | **0.62** | **1.12e-08** | **5.94e-08** | **2.87** | **0.004** | **0.009** |
| right hippocampus subcortical volume | 0.18 | 0.128 | 0.189 | **0.56** | **4.18e-07** | **1.78e-06** | **3.00** | **0.003** | **0.006** |
| right amygdala subcortical volume | 0.24 | 0.048 | 0.081 | **0.61** | **2.04e-08** | **1.04e-07** | **3.26** | **0.001** | **0.003** |
| right accumbens area subcortical volume | 0.19 | 0.122 | 0.181 | **0.41** | **4.20e-04** | **0.001** | 1.44 | 0.149 | 0.215 |
| right ventral diencephalon subcortical volume | **0.59** | **1.05e-07** | **4.86e-07** | **0.77** | **1.16e-14** | **1.34e-13** | **2.47** | **0.013** | **0.026** |
| right vessel subcortical volume | -0.05 | 0.666 | 0.740 | 0.11 | 0.359 | 0.456 | 0.98 | 0.328 | 0.422 |
| right choroid plexus subcortical volume | **0.50** | **1.01e-05** | **3.45e-05** | **0.49** | **2.02e-05** | **6.52e-05** | -0.21 | 0.833 | 0.880 |

| | | | | | | | | | |
|---|---|---|---|---|---|---|---|---|---|
| optic chiasm subcortical volume | 0.02 | 0.891 | 0.921 | 0.24 | 0.048 | 0.081 | 1.31 | 0.190 | 0.264 |
| corpus callosum posterior subcortical volume | **0.44** | **1.58e-04** | **4.42e-04** | **0.77** | **9.33e-15** | **1.09e-13** | **4.28** | **1.86e-05** | **6.06e-05** |
| corpus callosum mid posterior subcortical volume | 0.17 | 0.152 | 0.219 | **0.55** | **7.23e-07** | **2.96e-06** | **3.87** | **1.10e-04** | **3.16e-04** |
| corpus callosum central subcortical volume | **0.31** | **0.009** | **0.018** | **0.52** | **4.90e-06** | **1.78e-05** | 1.97 | 0.049 | 0.082 |
| corpus callosum mid anterior subcortical volume | **0.34** | **0.004** | **0.010** | **0.56** | **4.51e-07** | **1.92e-06** | **2.63** | **0.008** | **0.017** |
| corpus callosum anterior subcortical volume | **0.47** | **4.15e-05** | **1.27e-04** | **0.84** | **4.46e-20** | **1.45e-18** | **5.15** | **2.66e-07** | **1.17e-06** |

**STable 3.** Intra-class correlations (ICCs) of regional measurements from standard versus SynthSR-processed axial 64mT scans with 3T scans. Differences between ICC strengths were tested using Steiger's Z. A positive Z-value indicates that SynthSR-processed regions were more strongly correlated to 3T scans than standard regions. Analyses that are statistically significant after correction for multiple comparisons are in bold.

| **Measurement** | **Standard Axial 64mT ICC with 3T** | | | **SynthSR−Processed Axial 64mT ICC with 3T** | | | **Steiger** | | |
|---|---|---|---|---|---|---|---|---|---|
| | ***ICC*** | ***p*** | ***q*** | ***ICC*** | ***p*** | ***q*** | ***z*** | ***p*** | ***q*** |
| left banks of superior temporal sulcus thickness | -0.20 | 0.952 | 0.977 | -0.06 | 0.684 | 0.774 | 0.96 | 0.336 | 0.443 |
| left caudal anterior cingulate thickness | 0.01 | 0.462 | 0.572 | 0.23 | 0.028 | 0.051 | 1.44 | 0.149 | 0.224 |
| left caudal middle frontal thickness | 0.17 | 0.079 | 0.130 | **0.35** | **0.001** | **0.003** | 1.29 | 0.199 | 0.286 |
| left cuneus thickness | 0.09 | 0.239 | 0.336 | 0.14 | 0.129 | 0.200 | 0.32 | 0.748 | 0.827 |
| left entorhinal thickness | -5.2e-03 | 0.517 | 0.621 | 4.4e-03 | 0.485 | 0.592 | 0.06 | 0.954 | 0.977 |
| left fusiform thickness | -0.12 | 0.847 | 0.907 | 0.16 | 0.092 | 0.149 | 1.65 | 0.100 | 0.160 |
| left inferior parietal thickness | -0.15 | 0.898 | 0.941 | 0.16 | 0.085 | 0.139 | **2.29** | **0.022** | **0.041** |
| left inferior temporal thickness | 0.07 | 0.293 | 0.396 | 0.16 | 0.091 | 0.147 | 0.55 | 0.584 | 0.683 |
| left isthmus cingulate thickness | -0.02 | 0.577 | 0.678 | 0.08 | 0.267 | 0.368 | 0.54 | 0.589 | 0.687 |
| left lateral occipital thickness | 7.3e-03 | 0.476 | 0.584 | 0.06 | 0.300 | 0.402 | 0.36 | 0.718 | 0.803 |
| left lateral orbitofrontal thickness | -0.03 | 0.609 | 0.705 | 0.19 | 0.056 | 0.095 | 1.53 | 0.126 | 0.196 |
| left lingual thickness | 0.07 | 0.274 | 0.376 | -0.06 | 0.684 | 0.774 | -0.71 | 0.477 | 0.585 |
| left medial orbitofrontal thickness | -0.29 | 0.992 | 0.996 | 0.17 | 0.073 | 0.122 | **2.51** | **0.012** | **0.024** |
| left middle temporal thickness | -0.19 | 0.940 | 0.972 | **0.29** | **0.006** | **0.013** | **2.76** | **0.006** | **0.012** |
| left parahippocampal thickness | 0.01 | 0.454 | 0.565 | **0.35** | **0.001** | **0.003** | 2.11 | 0.035 | 0.062 |
| left paracentral thickness | 0.05 | 0.346 | 0.454 | 0.04 | 0.380 | 0.491 | -0.07 | 0.946 | 0.974 |
| left pars triangularis thickness | **0.29** | **0.007** | **0.016** | **0.33** | **0.002** | **0.005** | 0.32 | 0.748 | 0.827 |
| left pars opercularis thickness | -0.12 | 0.842 | 0.903 | -0.02 | 0.578 | 0.678 | 0.70 | 0.485 | 0.591 |
| left pars orbitalis thickness | -0.19 | 0.943 | 0.973 | -2.8e-05 | 0.500 | 0.604 | 1.19 | 0.235 | 0.332 |
| left pericalcarine thickness | 0.07 | 0.274 | 0.376 | -0.20 | 0.953 | 0.977 | -1.72 | 0.085 | 0.139 |
| left postcentral thickness | 0.12 | 0.151 | 0.226 | 0.22 | 0.034 | 0.061 | 0.62 | 0.538 | 0.639 |
| left posterior cingulate thickness | -0.13 | 0.863 | 0.913 | **0.23** | **0.026** | **0.047** | 2.12 | 0.034 | 0.061 |
| left precentral thickness | 0.08 | 0.244 | 0.342 | 0.19 | 0.061 | 0.103 | 0.70 | 0.485 | 0.591 |
| left precuneus thickness | 0.06 | 0.305 | 0.408 | **0.40** | **2.87e-04** | **7.85e-04** | **2.28** | **0.023** | **0.042** |
| left rostral anterior cingulate thickness | -0.24 | 0.978 | 0.991 | -0.02 | 0.552 | 0.655 | 1.54 | 0.124 | 0.192 |
| left rostral middle frontal thickness | -0.01 | 0.542 | 0.643 | 0.09 | 0.232 | 0.327 | 0.79 | 0.428 | 0.539 |
| left superior frontal thickness | 0.02 | 0.421 | 0.534 | **0.36** | **0.001** | **0.003** | **2.68** | **0.007** | **0.015** |
| left superior parietal thickness | 0.04 | 0.373 | 0.485 | 0.11 | 0.184 | 0.268 | 0.43 | 0.666 | 0.757 |
| left superior temporal thickness | -0.20 | 0.952 | 0.977 | **0.31** | **0.004** | **0.010** | **2.90** | **0.004** | **0.008** |
| left supramarginal thickness | 0.02 | 0.438 | 0.550 | **0.41** | **2.07e-04** | **5.82e-04** | **2.44** | **0.015** | **0.029** |

| | | | | | | | | | |
|---|---|---|---|---|---|---|---|---|---|
| left frontal pole thickness | -0.22 | 0.968 | 0.986 | -0.33 | 0.998 | 0.998 | -0.71 | 0.478 | 0.586 |
| left temporal pole thickness | -0.10 | 0.799 | 0.869 | 0.13 | 0.136 | 0.208 | 1.35 | 0.177 | 0.259 |
| left transverse temporal thickness | -0.25 | 0.980 | 0.993 | **0.27** | **0.012** | **0.024** | **3.08** | **0.002** | **0.005** |
| left insula thickness | -0.06 | 0.699 | 0.785 | -0.09 | 0.775 | 0.851 | -0.19 | 0.852 | 0.908 |
| right banks of superior temporal sulcus thickness | -0.10 | 0.789 | 0.862 | 0.10 | 0.196 | 0.282 | 1.39 | 0.163 | 0.242 |
| right caudal anterior cingulate thickness | -0.13 | 0.859 | 0.912 | 8.8e-03 | 0.471 | 0.581 | 0.76 | 0.444 | 0.556 |
| right caudal middle frontal thickness | 0.05 | 0.344 | 0.452 | **0.29** | **0.007** | **0.015** | 1.49 | 0.137 | 0.209 |
| right cuneus thickness | 0.07 | 0.277 | 0.379 | 0.14 | 0.122 | 0.190 | 0.39 | 0.694 | 0.782 |
| right entorhinal thickness | -0.10 | 0.803 | 0.872 | -0.05 | 0.665 | 0.757 | 0.29 | 0.768 | 0.846 |
| right fusiform thickness | -0.14 | 0.882 | 0.925 | -0.32 | 0.997 | 0.997 | -1.14 | 0.256 | 0.355 |
| right inferior parietal thickness | -0.17 | 0.923 | 0.960 | 0.06 | 0.323 | 0.428 | 1.60 | 0.109 | 0.173 |
| right inferior temporal thickness | -0.10 | 0.801 | 0.871 | -2.7e-03 | 0.509 | 0.614 | 0.63 | 0.528 | 0.630 |
| right isthmus cingulate thickness | -0.01 | 0.537 | 0.639 | 0.06 | 0.305 | 0.408 | 0.43 | 0.667 | 0.758 |
| right lateral occipital thickness | 0.13 | 0.139 | 0.212 | 0.06 | 0.323 | 0.428 | -0.57 | 0.567 | 0.671 |
| right lateral orbitofrontal thickness | -0.04 | 0.637 | 0.731 | 0.09 | 0.237 | 0.334 | 0.75 | 0.454 | 0.565 |
| right lingual thickness | -0.03 | 0.610 | 0.705 | 0.06 | 0.324 | 0.428 | 0.52 | 0.603 | 0.700 |
| right medial orbitofrontal thickness | -0.31 | 0.996 | 0.997 | -0.05 | 0.651 | 0.745 | 1.78 | 0.075 | 0.124 |
| right middle temporal thickness | -0.27 | 0.988 | 0.995 | 0.17 | 0.082 | 0.134 | **2.65** | **0.008** | **0.017** |
| right parahippocampal thickness | 0.08 | 0.252 | 0.350 | **0.38** | **5.29e-04** | **0.001** | 1.81 | 0.071 | 0.119 |
| right paracentral thickness | 0.14 | 0.125 | 0.194 | 0.09 | 0.232 | 0.327 | -0.33 | 0.745 | 0.825 |
| right pars triangularis thickness | 0.19 | 0.059 | 0.100 | 0.20 | 0.051 | 0.089 | 0.06 | 0.954 | 0.977 |
| right pars opercularis thickness | -0.04 | 0.638 | 0.732 | **0.27** | **0.011** | **0.022** | 1.94 | 0.053 | 0.091 |
| right pars orbitalis thickness | 2.8e-03 | 0.491 | 0.597 | **0.24** | **0.020** | **0.038** | 1.76 | 0.078 | 0.129 |
| right pericalcarine thickness | -0.02 | 0.572 | 0.674 | -7.4e-03 | 0.524 | 0.627 | 0.09 | 0.926 | 0.962 |
| right postcentral thickness | 0.15 | 0.113 | 0.177 | **0.24** | **0.020** | **0.039** | 0.71 | 0.475 | 0.584 |
| right posterior cingulate thickness | -0.07 | 0.731 | 0.815 | 0.23 | 0.028 | 0.051 | 1.99 | 0.047 | 0.082 |
| right precentral thickness | 0.16 | 0.092 | 0.148 | -0.05 | 0.673 | 0.763 | -1.60 | 0.110 | 0.174 |
| right precuneus thickness | 0.06 | 0.299 | 0.402 | 0.20 | 0.047 | 0.082 | 0.93 | 0.354 | 0.463 |
| right rostral anterior cingulate thickness | -0.30 | 0.994 | 0.997 | -0.12 | 0.835 | 0.897 | 1.17 | 0.243 | 0.341 |
| right rostral middle frontal thickness | -0.18 | 0.932 | 0.966 | 0.03 | 0.413 | 0.526 | 1.36 | 0.173 | 0.255 |
| right superior frontal thickness | 0.16 | 0.098 | 0.157 | 0.11 | 0.186 | 0.269 | -0.34 | 0.734 | 0.817 |
| right superior parietal thickness | 0.10 | 0.200 | 0.288 | 0.08 | 0.265 | 0.366 | -0.18 | 0.858 | 0.911 |
| right superior temporal thickness | -0.17 | 0.922 | 0.959 | **0.43** | **1.10e-04** | **3.22e-04** | **4.02** | **5.82e-05** | **1.79e-04** |
| right supramarginal thickness | 0.18 | 0.062 | 0.105 | **0.39** | **3.63e-04** | **9.73e-04** | 1.63 | 0.104 | 0.165 |

| | | | | | | | | | |
|---|---|---|---|---|---|---|---|---|---|
| right frontal pole thickness | -0.09 | 0.780 | 0.856 | -0.12 | 0.848 | 0.907 | -0.18 | 0.856 | 0.911 |
| right temporal pole thickness | 0.10 | 0.213 | 0.304 | 0.22 | 0.031 | 0.056 | 0.79 | 0.430 | 0.541 |
| right transverse temporal thickness | -0.02 | 0.572 | 0.674 | 0.06 | 0.296 | 0.399 | 0.54 | 0.590 | 0.688 |
| right insula thickness | -0.06 | 0.678 | 0.768 | 0.15 | 0.104 | 0.166 | 1.20 | 0.230 | 0.325 |
| left banks of superior temporal sulcus area | **0.34** | **0.002** | **0.004** | **0.73** | **3.00e-13** | **2.91e-12** | **3.94** | **8.19e-05** | **2.46e-04** |
| left caudal anterior cingulate area | **0.28** | **0.008** | **0.017** | **0.44** | **7.06e-05** | **2.14e-04** | 1.78 | 0.075 | 0.124 |
| left caudal middle frontal area | **0.29** | **0.007** | **0.015** | **0.73** | **1.89e-13** | **1.88e-12** | **4.19** | **2.76e-05** | **9.10e-05** |
| left cuneus area | **0.41** | **2.15e-04** | **6.02e-04** | **0.62** | **2.91e-09** | **1.69e-08** | 1.78 | 0.075 | 0.124 |
| left entorhinal area | -0.06 | 0.692 | 0.781 | **0.30** | **0.006** | **0.013** | 2.00 | 0.046 | 0.081 |
| left fusiform area | **0.31** | **0.005** | **0.010** | **0.64** | **1.02e-09** | **6.21e-09** | **2.50** | **0.012** | **0.025** |
| left inferior parietal area | **0.57** | **1.03e-07** | **4.99e-07** | **0.84** | **3.69e-20** | **1.22e-18** | **3.75** | **1.79e-04** | **5.09e-04** |
| left inferior temporal area | **0.42** | **1.25e-04** | **3.64e-04** | **0.72** | **5.88e-13** | **5.57e-12** | **2.87** | **0.004** | **0.009** |
| left isthmus cingulate area | **0.33** | **0.002** | **0.005** | **0.50** | **3.76e-06** | **1.43e-05** | 1.24 | 0.214 | 0.306 |
| left lateral occipital area | **0.32** | **0.003** | **0.007** | **0.52** | **1.99e-06** | **7.90e-06** | 1.44 | 0.151 | 0.226 |
| left lateral orbitofrontal area | **0.51** | **2.90e-06** | **1.13e-05** | **0.67** | **9.41e-11** | **6.54e-10** | 1.74 | 0.082 | 0.134 |
| left lingual area | **0.24** | **0.023** | **0.042** | **0.58** | **4.25e-08** | **2.13e-07** | **2.51** | **0.012** | **0.024** |
| left medial orbitofrontal area | **0.48** | **1.09e-05** | **3.84e-05** | **0.50** | **3.73e-06** | **1.42e-05** | 0.22 | 0.824 | 0.889 |
| left middle temporal area | **0.45** | **4.19e-05** | **1.32e-04** | **0.72** | **8.58e-13** | **7.88e-12** | **2.70** | **0.007** | **0.015** |
| left parahippocampal area | -0.16 | 0.906 | 0.947 | 0.21 | 0.039 | 0.069 | 2.06 | 0.039 | 0.070 |
| left paracentral area | **0.34** | **0.002** | **0.004** | **0.75** | **3.25e-14** | **3.61e-13** | **3.73** | **1.93e-04** | **5.47e-04** |
| left pars triangularis area | **0.40** | **3.23e-04** | **8.75e-04** | **0.65** | **4.52e-10** | **2.87e-09** | **2.54** | **0.011** | **0.022** |
| left pars opercularis area | **0.45** | **4.43e-05** | **1.39e-04** | **0.48** | **1.10e-05** | **3.87e-05** | 0.27 | 0.789 | 0.862 |
| left pars orbitalis area | **0.59** | **4.06e-08** | **2.04e-07** | **0.63** | **1.97e-09** | **1.18e-08** | 0.48 | 0.631 | 0.726 |
| left pericalcarine area | **0.33** | **0.002** | **0.006** | **0.65** | **5.25e-10** | **3.31e-09** | **2.78** | **0.005** | **0.012** |
| left postcentral area | 0.20 | 0.044 | 0.078 | **0.70** | **5.61e-12** | **4.67e-11** | **4.25** | **2.09e-05** | **7.02e-05** |
| left posterior cingulate area | **0.30** | **0.006** | **0.013** | **0.57** | **8.80e-08** | **4.26e-07** | **2.48** | **0.013** | **0.026** |
| left precentral area | **0.35** | **0.001** | **0.003** | **0.82** | **6.55e-19** | **1.68e-17** | **5.50** | **3.76e-08** | **1.90e-07** |
| left precuneus area | **0.31** | **0.004** | **0.010** | **0.72** | **6.43e-13** | **6.07e-12** | **3.58** | **3.48e-04** | **9.36e-04** |
| left rostral anterior cingulate area | **0.33** | **0.003** | **0.006** | **0.60** | **1.25e-08** | **6.71e-08** | **2.70** | **0.007** | **0.014** |
| left rostral middle frontal area | **0.56** | **2.02e-07** | **9.38e-07** | **0.73** | **1.92e-13** | **1.91e-12** | **2.52** | **0.012** | **0.024** |
| left superior frontal area | **0.52** | **1.89e-06** | **7.54e-06** | **0.76** | **5.76e-15** | **6.97e-14** | **3.22** | **0.001** | **0.003** |
| left superior parietal area | **0.37** | **7.83e-04** | **0.002** | **0.67** | **1.04e-10** | **7.15e-10** | **2.61** | **0.009** | **0.018** |
| left superior temporal area | **0.29** | **0.008** | **0.016** | **0.78** | **6.94e-16** | **1.00e-14** | **5.14** | **2.79e-07** | **1.26e-06** |
| left supramarginal area | **0.50** | **3.88e-06** | **1.47e-05** | **0.78** | **7.28e-16** | **1.04e-14** | **3.28** | **0.001** | **0.003** |
| left frontal pole area | **0.24** | **0.020** | **0.038** | **0.24** | **0.020** | **0.038** | 5.8e-03 | 0.995 | 0.997 |

| | | | | | | | | | |
|---|---|---|---|---|---|---|---|---|---|
| left temporal pole area | -0.06 | 0.693 | 0.781 | 0.12 | 0.151 | 0.226 | 1.06 | 0.289 | 0.391 |
| left transverse temporal area | 0.13 | 0.145 | 0.219 | **0.72** | **9.76e-13** | **8.85e-12** | **4.52** | **6.20e-06** | **2.30e-05** |
| left insula area | **0.27** | **0.012** | **0.024** | **0.77** | **3.37e-15** | **4.37e-14** | **4.63** | **3.73e-06** | **1.42e-05** |
| right banks of superior temporal sulcus area | **0.39** | **3.81e-04** | **0.001** | **0.68** | **3.30e-11** | **2.38e-10** | **2.65** | **0.008** | **0.017** |
| right caudal anterior cingulate area | **0.49** | **8.19e-06** | **2.97e-05** | **0.71** | **2.68e-12** | **2.30e-11** | **2.51** | **0.012** | **0.024** |
| right caudal middle frontal area | **0.40** | **2.76e-04** | **7.58e-04** | **0.60** | **2.11e-08** | **1.11e-07** | 1.93 | 0.054 | 0.093 |
| right cuneus area | 0.17 | 0.073 | 0.122 | **0.51** | **2.92e-06** | **1.13e-05** | **2.79** | **0.005** | **0.011** |
| right entorhinal area | -0.13 | 0.863 | 0.913 | 0.12 | 0.151 | 0.226 | 1.62 | 0.106 | 0.168 |
| right fusiform area | **0.43** | **1.03e-04** | **3.01e-04** | **0.76** | **6.27e-15** | **7.50e-14** | **3.38** | **7.38e-04** | **0.002** |
| right inferior parietal area | **0.49** | **6.16e-06** | **2.28e-05** | **0.74** | **1.08e-13** | **1.13e-12** | **2.88** | **0.004** | **0.009** |
| right inferior temporal area | **0.52** | **1.36e-06** | **5.57e-06** | **0.76** | **5.31e-15** | **6.49e-14** | **2.80** | **0.005** | **0.011** |
| right isthmus cingulate area | 0.17 | 0.072 | 0.121 | **0.72** | **5.67e-13** | **5.40e-12** | **4.78** | **1.80e-06** | **7.19e-06** |
| right lateral occipital area | **0.54** | **6.37e-07** | **2.77e-06** | **0.64** | **1.07e-09** | **6.51e-09** | 1.03 | 0.305 | 0.408 |
| right lateral orbitofrontal area | **0.51** | **3.26e-06** | **1.25e-05** | **0.54** | **5.13e-07** | **2.25e-06** | 0.35 | 0.724 | 0.809 |
| right lingual area | 0.10 | 0.209 | 0.299 | **0.60** | **2.15e-08** | **1.13e-07** | **4.30** | **1.73e-05** | **5.85e-05** |
| right medial orbitofrontal area | **0.43** | **1.05e-04** | **3.09e-04** | **0.44** | **5.48e-05** | **1.70e-04** | 0.13 | 0.897 | 0.940 |
| right middle temporal area | **0.45** | **4.27e-05** | **1.35e-04** | **0.74** | **5.16e-14** | **5.54e-13** | **3.38** | **7.28e-04** | **0.002** |
| right parahippocampal area | 0.02 | 0.433 | 0.545 | **0.28** | **0.009** | **0.018** | 1.57 | 0.117 | 0.184 |
| right paracentral area | **0.36** | **9.93e-04** | **0.002** | **0.61** | **6.80e-09** | **3.77e-08** | **2.41** | **0.016** | **0.031** |
| right pars triangularis area | **0.42** | **1.46e-04** | **4.19e-04** | **0.75** | **2.16e-14** | **2.48e-13** | **3.97** | **7.29e-05** | **2.21e-04** |
| right pars opercularis area | **0.48** | **1.10e-05** | **3.85e-05** | **0.54** | **7.38e-07** | **3.16e-06** | 0.51 | 0.610 | 0.705 |
| right pars orbitalis area | **0.35** | **0.001** | **0.003** | **0.64** | **1.09e-09** | **6.59e-09** | **2.85** | **0.004** | **0.010** |
| right pericalcarine area | **0.44** | **6.86e-05** | **2.09e-04** | **0.68** | **4.82e-11** | **3.43e-10** | **2.55** | **0.011** | **0.022** |
| right postcentral area | **0.43** | **7.50e-05** | **2.26e-04** | **0.68** | **3.47e-11** | **2.50e-10** | **2.76** | **0.006** | **0.013** |
| right posterior cingulate area | **0.32** | **0.003** | **0.007** | **0.69** | **8.85e-12** | **7.14e-11** | **4.14** | **3.50e-05** | **1.12e-04** |
| right precentral area | **0.48** | **1.16e-05** | **4.03e-05** | **0.63** | **2.70e-09** | **1.58e-08** | 1.51 | 0.132 | 0.203 |
| right precuneus area | **0.43** | **8.69e-05** | **2.58e-04** | **0.70** | **5.93e-12** | **4.91e-11** | **2.60** | **0.009** | **0.019** |
| right rostral anterior cingulate area | **0.41** | **1.91e-04** | **5.40e-04** | **0.78** | **4.33e-16** | **6.50e-15** | **4.32** | **1.56e-05** | **5.32e-05** |
| right rostral middle frontal area | **0.40** | **2.73e-04** | **7.50e-04** | **0.64** | **1.18e-09** | **7.13e-09** | **2.42** | **0.016** | **0.030** |
| right superior frontal area | **0.51** | **3.49e-06** | **1.34e-05** | **0.69** | **2.11e-11** | **1.58e-10** | **2.32** | **0.020** | **0.038** |
| right superior parietal area | 0.21 | 0.040 | 0.071 | **0.60** | **2.17e-08** | **1.13e-07** | **3.14** | **0.002** | **0.004** |
| right superior temporal area | **0.48** | **1.18e-05** | **4.10e-05** | **0.77** | **3.04e-15** | **3.96e-14** | **3.52** | **4.36e-04** | **0.001** |
| right supramarginal area | **0.27** | **0.011** | **0.023** | **0.66** | **2.02e-10** | **1.34e-09** | **3.68** | **2.33e-04** | **6.52e-04** |
| right frontal pole area | **0.26** | **0.014** | **0.028** | 0.13 | 0.139 | 0.212 | -0.88 | 0.381 | 0.492 |

| | | | | | | | | | |
|---|---|---|---|---|---|---|---|---|---|
| right temporal pole area | 4.7e-03 | 0.485 | 0.591 | 0.06 | 0.314 | 0.417 | 0.32 | 0.747 | 0.826 |
| right transverse temporal area | **0.25** | **0.016** | **0.031** | **0.58** | **4.47e-08** | **2.23e-07** | **2.64** | **0.008** | **0.017** |
| right insula area | 0.16 | 0.087 | 0.142 | **0.66** | **1.30e-10** | **8.82e-10** | **3.80** | **1.42e-04** | **4.10e-04** |
| left banks of superior temporal sulcus volume | **0.25** | **0.017** | **0.033** | **0.67** | **1.01e-10** | **6.96e-10** | **4.31** | **1.61e-05** | **5.49e-05** |
| left caudal anterior cingulate volume | **0.29** | **0.008** | **0.016** | **0.54** | **6.53e-07** | **2.83e-06** | **3.09** | **0.002** | **0.005** |
| left caudal middle frontal volume | **0.34** | **0.002** | **0.004** | **0.69** | **1.83e-11** | **1.42e-10** | **3.25** | **0.001** | **0.003** |
| left cuneus volume | **0.30** | **0.005** | **0.011** | **0.65** | **3.69e-10** | **2.37e-09** | **2.86** | **0.004** | **0.009** |
| left entorhinal volume | -0.10 | 0.798 | 0.868 | **0.31** | **0.004** | **0.009** | **2.31** | **0.021** | **0.039** |
| left fusiform volume | **0.31** | **0.005** | **0.010** | **0.66** | **2.41e-10** | **1.58e-09** | **2.70** | **0.007** | **0.014** |
| left inferior parietal volume | **0.53** | **1.21e-06** | **5.01e-06** | **0.82** | **9.68e-19** | **2.32e-17** | **3.92** | **8.98e-05** | **2.65e-04** |
| left inferior temporal volume | **0.45** | **4.57e-05** | **1.43e-04** | **0.78** | **9.63e-16** | **1.34e-14** | **3.55** | **3.86e-04** | **0.001** |
| left isthmus cingulate volume | **0.30** | **0.006** | **0.013** | **0.49** | **6.05e-06** | **2.25e-05** | 1.38 | 0.166 | 0.246 |
| left lateral occipital volume | **0.39** | **4.22e-04** | **0.001** | **0.55** | **4.08e-07** | **1.81e-06** | 1.36 | 0.175 | 0.257 |
| left lateral orbitofrontal volume | **0.47** | **2.19e-05** | **7.34e-05** | **0.69** | **2.11e-11** | **1.58e-10** | **2.28** | **0.023** | **0.043** |
| left lingual volume | 0.12 | 0.155 | 0.231 | **0.65** | **4.22e-10** | **2.69e-09** | **4.14** | **3.46e-05** | **1.11e-04** |
| left medial orbitofrontal volume | **0.28** | **0.009** | **0.018** | **0.41** | **1.72e-04** | **4.93e-04** | 1.05 | 0.296 | 0.399 |
| left middle temporal volume | **0.30** | **0.006** | **0.013** | **0.71** | **2.30e-12** | **1.99e-11** | **3.78** | **1.57e-04** | **4.52e-04** |
| left parahippocampal volume | -0.03 | 0.603 | 0.700 | 0.16 | 0.087 | 0.141 | 1.15 | 0.252 | 0.350 |
| left paracentral volume | **0.31** | **0.004** | **0.009** | **0.65** | **4.69e-10** | **2.97e-09** | **2.82** | **0.005** | **0.011** |
| left pars triangularis volume | **0.46** | **3.10e-05** | **1.00e-04** | **0.62** | **4.56e-09** | **2.59e-08** | 1.89 | 0.058 | 0.100 |
| left pars opercularis volume | **0.35** | **0.001** | **0.003** | **0.43** | **8.63e-05** | **2.56e-04** | 0.64 | 0.520 | 0.623 |
| left pars orbitalis volume | **0.53** | **7.98e-07** | **3.40e-06** | **0.58** | **5.73e-08** | **2.82e-07** | 0.46 | 0.646 | 0.740 |
| left pericalcarine volume | **0.31** | **0.004** | **0.009** | **0.56** | **1.67e-07** | **7.79e-07** | 2.06 | 0.039 | 0.070 |
| left postcentral volume | 0.22 | 0.034 | 0.061 | **0.67** | **1.19e-10** | **8.14e-10** | **3.70** | **2.13e-04** | **5.99e-04** |
| left posterior cingulate volume | **0.27** | **0.011** | **0.022** | **0.54** | **4.56e-07** | **2.01e-06** | **2.41** | **0.016** | **0.031** |
| left precentral volume | **0.35** | **0.001** | **0.003** | **0.77** | **2.03e-15** | **2.72e-14** | **5.02** | **5.23e-07** | **2.29e-06** |
| left precuneus volume | **0.27** | **0.011** | **0.022** | **0.69** | **1.22e-11** | **9.68e-11** | **3.65** | **2.62e-04** | **7.27e-04** |
| left rostral anterior cingulate volume | **0.32** | **0.003** | **0.007** | **0.51** | **2.28e-06** | **8.97e-06** | 1.79 | 0.073 | 0.122 |
| left rostral middle frontal volume | **0.46** | **2.40e-05** | **8.00e-05** | **0.56** | **1.50e-07** | **7.07e-07** | 1.18 | 0.240 | 0.337 |
| left superior frontal volume | **0.36** | **0.001** | **0.003** | **0.69** | **1.90e-11** | **1.46e-10** | **4.11** | **3.93e-05** | **1.24e-04** |
| left superior parietal volume | **0.46** | **2.27e-05** | **7.58e-05** | **0.62** | **3.38e-09** | **1.95e-08** | 1.38 | 0.166 | 0.246 |
| left superior temporal volume | 0.19 | 0.055 | 0.094 | **0.80** | **5.70e-17** | **9.85e-16** | **6.15** | **7.76e-10** | **4.83e-09** |
| left supramarginal volume | **0.38** | **5.15e-04** | **0.001** | **0.69** | **2.04e-11** | **1.54e-10** | **3.10** | **0.002** | **0.005** |
| left frontal pole volume | -0.04 | 0.624 | 0.718 | -0.10 | 0.808 | 0.876 | -0.45 | 0.652 | 0.745 |
| left temporal pole volume | -0.07 | 0.707 | 0.792 | 0.07 | 0.282 | 0.384 | 0.72 | 0.469 | 0.579 |

| | | | | | | | | | |
|---|---|---|---|---|---|---|---|---|---|
| left transverse temporal volume | 0.08 | 0.243 | 0.340 | **0.68** | **3.37e-11** | **2.43e-10** | **4.44** | **9.08e-06** | **3.26e-05** |
| left insula volume | **0.41** | **1.80e-04** | **5.12e-04** | **0.73** | **3.20e-13** | **3.09e-12** | **3.31** | **9.27e-04** | **0.002** |
| right banks of superior temporal sulcus volume | **0.39** | **3.36e-04** | **9.07e-04** | **0.60** | **2.11e-08** | **1.11e-07** | 1.78 | 0.075 | 0.124 |
| right caudal anterior cingulate volume | **0.58** | **6.08e-08** | **2.99e-07** | **0.71** | **2.04e-12** | **1.77e-11** | 1.75 | 0.080 | 0.131 |
| right caudal middle frontal volume | **0.40** | **3.11e-04** | **8.47e-04** | **0.53** | **8.58e-07** | **3.65e-06** | 1.28 | 0.200 | 0.288 |
| right cuneus volume | 0.16 | 0.093 | 0.150 | **0.51** | **3.26e-06** | **1.25e-05** | **2.79** | **0.005** | **0.011** |
| right entorhinal volume | -0.21 | 0.961 | 0.982 | 0.18 | 0.067 | 0.113 | **2.41** | **0.016** | **0.031** |
| right fusiform volume | **0.31** | **0.005** | **0.011** | **0.71** | **2.11e-12** | **1.83e-11** | **3.58** | **3.47e-04** | **9.35e-04** |
| right inferior parietal volume | **0.42** | **1.33e-04** | **3.85e-04** | **0.69** | **1.57e-11** | **1.23e-10** | **3.20** | **0.001** | **0.003** |
| right inferior temporal volume | **0.47** | **1.43e-05** | **4.92e-05** | **0.76** | **5.40e-15** | **6.57e-14** | **3.26** | **0.001** | **0.003** |
| right isthmus cingulate volume | 0.18 | 0.072 | 0.120 | **0.69** | **1.35e-11** | **1.06e-10** | **4.40** | **1.08e-05** | **3.80e-05** |
| right lateral occipital volume | **0.56** | **1.46e-07** | **6.91e-07** | **0.59** | **2.82e-08** | **1.46e-07** | 0.29 | 0.768 | 0.846 |
| right lateral orbitofrontal volume | **0.49** | **9.01e-06** | **3.24e-05** | **0.60** | **1.80e-08** | **9.56e-08** | 1.15 | 0.251 | 0.350 |
| right lingual volume | 0.04 | 0.381 | 0.492 | **0.64** | **8.36e-10** | **5.19e-09** | **5.25** | **1.49e-07** | **7.03e-07** |
| right medial orbitofrontal volume | **0.27** | **0.010** | **0.021** | **0.31** | **0.005** | **0.010** | 0.24 | 0.809 | 0.877 |
| right middle temporal volume | **0.38** | **5.09e-04** | **0.001** | **0.68** | **3.21e-11** | **2.34e-10** | **3.13** | **0.002** | **0.004** |
| right parahippocampal volume | 0.09 | 0.233 | 0.329 | 0.03 | 0.393 | 0.504 | -0.33 | 0.741 | 0.822 |
| right paracentral volume | **0.45** | **3.43e-05** | **1.10e-04** | **0.54** | **6.99e-07** | **3.01e-06** | 0.80 | 0.423 | 0.535 |
| right pars triangularis volume | **0.43** | **8.39e-05** | **2.50e-04** | **0.61** | **1.12e-08** | **6.07e-08** | 1.89 | 0.059 | 0.100 |
| right pars opercularis volume | **0.48** | **1.40e-05** | **4.80e-05** | **0.55** | **3.09e-07** | **1.38e-06** | 0.74 | 0.459 | 0.569 |
| right pars orbitalis volume | **0.36** | **0.001** | **0.003** | **0.59** | **3.77e-08** | **1.90e-07** | **2.38** | **0.017** | **0.033** |
| right pericalcarine volume | **0.32** | **0.003** | **0.007** | **0.61** | **6.49e-09** | **3.61e-08** | **2.73** | **0.006** | **0.014** |
| right postcentral volume | **0.48** | **9.43e-06** | **3.37e-05** | **0.67** | **5.49e-11** | **3.89e-10** | **2.26** | **0.024** | **0.045** |
| right posterior cingulate volume | **0.37** | **8.09e-04** | **0.002** | **0.67** | **8.77e-11** | **6.14e-10** | **3.29** | **0.001** | **0.002** |
| right precentral volume | **0.48** | **1.06e-05** | **3.74e-05** | **0.57** | **7.96e-08** | **3.87e-07** | 1.04 | 0.299 | 0.402 |
| right precuneus volume | **0.31** | **0.004** | **0.010** | **0.66** | **2.65e-10** | **1.73e-09** | **3.16** | **0.002** | **0.004** |
| right rostral anterior cingulate volume | **0.40** | **2.69e-04** | **7.42e-04** | **0.69** | **1.10e-11** | **8.75e-11** | **3.00** | **0.003** | **0.006** |
| right rostral middle frontal volume | **0.26** | **0.015** | **0.030** | **0.50** | **5.66e-06** | **2.11e-05** | **2.58** | **0.010** | **0.020** |
| right superior frontal volume | **0.43** | **8.13e-05** | **2.44e-04** | **0.60** | **1.26e-08** | **6.74e-08** | 2.11 | 0.035 | 0.062 |
| right superior parietal volume | **0.25** | **0.017** | **0.033** | **0.53** | **8.90e-07** | **3.78e-06** | **2.36** | **0.018** | **0.035** |
| right superior temporal volume | **0.53** | **1.04e-06** | **4.37e-06** | **0.78** | **2.76e-16** | **4.29e-15** | **3.45** | **5.68e-04** | **0.001** |
| right supramarginal volume | **0.34** | **0.002** | **0.004** | **0.71** | **1.85e-12** | **1.62e-11** | **4.20** | **2.62e-05** | **8.66e-05** |
| right frontal pole volume | 0.03 | 0.396 | 0.506 | -0.05 | 0.666 | 0.757 | -0.52 | 0.606 | 0.701 |
| right temporal pole volume | 0.08 | 0.242 | 0.340 | 0.08 | 0.262 | 0.362 | -0.04 | 0.965 | 0.985 |

| | | | | | | | | | |
|---|---|---|---|---|---|---|---|---|---|
| right transverse temporal volume | 0.23 | 0.027 | 0.050 | **0.51** | **2.65e-06** | **1.04e-05** | **2.31** | **0.021** | **0.039** |
| right insula volume | 0.20 | 0.049 | 0.085 | **0.71** | **2.38e-12** | **2.05e-11** | **4.14** | **3.47e-05** | **1.11e-04** |
| left lateral ventricle subcortical volume | **0.95** | **2.89e-36** | **7.86e-34** | **0.97** | **5.38e-43** | **3.90e-40** | **2.34** | **0.019** | **0.037** |
| left inf lat vent subcortical volume | **0.53** | **9.80e-07** | **4.14e-06** | **0.52** | **1.82e-06** | **7.26e-06** | -0.11 | 0.915 | 0.953 |
| left cerebellum white matter subcortical volume | **0.44** | **6.51e-05** | **1.99e-04** | **0.39** | **4.40e-04** | **0.001** | -0.39 | 0.699 | 0.785 |
| left cerebellum cortex subcortical volume | **0.73** | **2.57e-13** | **2.51e-12** | **0.86** | **3.24e-22** | **1.76e-20** | **2.95** | **0.003** | **0.007** |
| left thalamus proper subcortical volume | **0.39** | **4.05e-04** | **0.001** | **0.78** | **4.59e-16** | **6.84e-15** | **3.90** | **9.50e-05** | **2.80e-04** |
| left caudate subcortical volume | **0.47** | **1.52e-05** | **5.19e-05** | **0.70** | **8.54e-12** | **6.94e-11** | **2.90** | **0.004** | **0.008** |
| left putamen subcortical volume | **0.38** | **5.65e-04** | **0.001** | **0.56** | **1.82e-07** | **8.46e-07** | 1.88 | 0.060 | 0.102 |
| left pallidum subcortical volume | **0.26** | **0.016** | **0.030** | **0.51** | **2.91e-06** | **1.13e-05** | 1.99 | 0.047 | 0.081 |
| 3rd ventricle subcortical volume | **0.69** | **2.00e-11** | **1.53e-10** | **0.90** | **1.59e-26** | **1.73e-24** | **4.64** | **3.51e-06** | **1.35e-05** |
| 4th ventricle subcortical volume | **0.60** | **1.65e-08** | **8.77e-08** | **0.85** | **4.68e-21** | **2.00e-19** | **3.95** | **7.89e-05** | **2.37e-04** |
| brain stem subcortical volume | **0.78** | **2.74e-16** | **4.29e-15** | **0.88** | **5.19e-24** | **3.90e-22** | **2.49** | **0.013** | **0.026** |
| left hippocampus subcortical volume | 0.17 | 0.073 | 0.121 | **0.46** | **2.59e-05** | **8.59e-05** | 1.98 | 0.047 | 0.082 |
| left amygdala subcortical volume | **0.27** | **0.011** | **0.023** | **0.46** | **3.03e-05** | **9.89e-05** | 1.52 | 0.128 | 0.198 |
| csf subcortical volume | **0.24** | **0.020** | **0.038** | **0.65** | **4.88e-10** | **3.08e-09** | **4.17** | **3.04e-05** | **9.90e-05** |
| left accumbens area subcortical volume | 7.6e-03 | 0.475 | 0.584 | **0.24** | **0.022** | **0.041** | 1.39 | 0.165 | 0.245 |
| left ventral diencephalon subcortical volume | **0.48** | **1.26e-05** | **4.35e-05** | **0.68** | **2.66e-11** | **1.95e-10** | 2.20 | 0.028 | 0.051 |
| left vessel subcortical volume | 8.6e-03 | 0.472 | 0.581 | 0.20 | 0.046 | 0.081 | 1.08 | 0.279 | 0.381 |
| left choroid plexus subcortical volume | **0.39** | **3.28e-04** | **8.89e-04** | **0.60** | **1.95e-08** | **1.03e-07** | **2.78** | **0.005** | **0.012** |
| right lateral ventricle subcortical volume | **0.92** | **1.53e-29** | **2.79e-27** | **0.96** | **1.34e-38** | **5.83e-36** | **2.62** | **0.009** | **0.018** |
| right inf lat vent subcortical volume | **0.45** | **4.81e-05** | **1.50e-04** | **0.47** | **1.90e-05** | **6.42e-05** | 0.17 | 0.867 | 0.915 |
| cerebellum white matter subcortical volume | **0.62** | **4.73e-09** | **2.67e-08** | **0.52** | **1.54e-06** | **6.27e-06** | -0.99 | 0.321 | 0.425 |
| cerebellum cortex subcortical volume | **0.68** | **2.26e-11** | **1.67e-10** | **0.84** | **6.18e-20** | **1.89e-18** | **2.72** | **0.006** | **0.014** |
| right thalamus proper subcortical volume | **0.29** | **0.006** | **0.014** | **0.71** | **1.35e-12** | **1.21e-11** | **3.93** | **8.39e-05** | **2.50e-04** |
| right caudate subcortical volume | 0.19 | 0.052 | 0.089 | **0.48** | **1.04e-05** | **3.69e-05** | **2.61** | **0.009** | **0.018** |
| right putamen subcortical volume | **0.29** | **0.007** | **0.014** | **0.70** | **5.13e-12** | **4.30e-11** | **3.55** | **3.90e-04** | **0.001** |
| right pallidum subcortical volume | **0.24** | **0.022** | **0.041** | **0.62** | **5.15e-09** | **2.88e-08** | **2.87** | **0.004** | **0.009** |
| right hippocampus subcortical volume | 0.14 | 0.117 | 0.184 | **0.55** | **2.79e-07** | **1.26e-06** | **3.12** | **0.002** | **0.004** |
| right amygdala subcortical volume | 0.19 | 0.052 | 0.089 | **0.58** | **4.45e-08** | **2.22e-07** | **3.27** | **0.001** | **0.003** |
| right accumbens area subcortical volume | 0.17 | 0.079 | 0.131 | **0.40** | **2.50e-04** | **6.95e-04** | 1.50 | 0.134 | 0.205 |
| right ventral diencephalon subcortical volume | **0.58** | **5.45e-08** | **2.70e-07** | **0.76** | **6.17e-15** | **7.42e-14** | **2.42** | **0.015** | **0.030** |
| right vessel subcortical volume | -7.9e-03 | 0.526 | 0.628 | 0.02 | 0.432 | 0.544 | 0.17 | 0.865 | 0.915 |
| right choroid plexus subcortical volume | **0.41** | **1.61e-04** | **4.63e-04** | **0.48** | **9.72e-06** | **3.46e-05** | 0.72 | 0.470 | 0.579 |
| optic chiasm subcortical volume | 0.02 | 0.446 | 0.558 | 0.20 | 0.045 | 0.078 | 1.10 | 0.270 | 0.371 |

| | | | | | | | | | |
|---|---|---|---|---|---|---|---|---|---|
| corpus callosum posterior subcortical volume | **0.34** | **0.002** | **0.004** | **0.75** | **2.91e-14** | **3.26e-13** | **4.15** | **3.34e-05** | **1.07e-04** |
| corpus callosum mid posterior subcortical volume | 0.13 | 0.133 | 0.205 | **0.52** | **1.33e-06** | **5.48e-06** | **3.14** | **0.002** | **0.004** |
| corpus callosum central subcortical volume | **0.29** | **0.007** | **0.014** | **0.42** | **1.31e-04** | **3.80e-04** | 1.01 | 0.313 | 0.417 |
| corpus callosum mid anterior subcortical volume | **0.33** | **0.002** | **0.005** | **0.45** | **3.79e-05** | **1.20e-04** | 1.08 | 0.279 | 0.381 |
| corpus callosum anterior subcortical volume | **0.43** | **9.05e-05** | **2.67e-04** | **0.82** | **9.41e-19** | **2.30e-17** | **4.66** | **3.09e-06** | **1.19e-05** |

**STable 4.** Pearson correlations of regional measurements from standard axial versus standard multi-orientation 64mT scans with 3T scans. Differences between correlation strengths were tested using Steiger's Z. A positive Z-value indicates that multi-orientation regions were more strongly correlated to 3T scans than axial-only regions. Analyses that are statistically significant after correction for multiple comparisons are in bold.

| Measurement | Standard Axial 64mT Correlations with 3T | | | Standard Multi-Orientation 64mT Correlations with 3T | | | Steiger | | |
|---|---|---|---|---|---|---|---|---|---|
| | ***r*** | ***p*** | ***q*** | ***r*** | ***p*** | ***q*** | ***z*** | ***p*** | ***q*** |
| left banks of superior temporal sulcus thickness | **-0.32** | **0.007** | **0.015** | -0.25 | 0.037 | 0.066 | 0.55 | 0.581 | 0.668 |
| left caudal anterior cingulate thickness | 0.01 | 0.914 | 0.939 | -0.10 | 0.404 | 0.503 | -0.83 | 0.407 | 0.504 |
| left caudal middle frontal thickness | 0.20 | 0.090 | 0.140 | -0.05 | 0.666 | 0.740 | -1.78 | 0.075 | 0.119 |
| left cuneus thickness | 0.13 | 0.276 | 0.365 | -0.02 | 0.878 | 0.913 | -1.08 | 0.281 | 0.370 |
| left entorhinal thickness | -0.02 | 0.896 | 0.926 | -0.20 | 0.096 | 0.148 | -1.09 | 0.276 | 0.365 |
| left fusiform thickness | -0.17 | 0.168 | 0.239 | -0.21 | 0.077 | 0.123 | -0.38 | 0.706 | 0.772 |
| left inferior parietal thickness | -0.19 | 0.123 | 0.183 | -0.23 | 0.055 | 0.091 | -0.39 | 0.694 | 0.763 |
| left inferior temporal thickness | 0.09 | 0.481 | 0.574 | -0.18 | 0.137 | 0.201 | -2.07 | 0.038 | 0.067 |
| left isthmus cingulate thickness | -0.03 | 0.829 | 0.878 | -0.21 | 0.079 | 0.125 | -1.18 | 0.240 | 0.326 |
| left lateral occipital thickness | 0.01 | 0.929 | 0.949 | -0.01 | 0.931 | 0.949 | -0.19 | 0.851 | 0.893 |
| left lateral orbitofrontal thickness | -0.04 | 0.771 | 0.830 | 0.02 | 0.879 | 0.913 | 0.43 | 0.670 | 0.742 |
| left lingual thickness | 0.09 | 0.449 | 0.542 | -0.15 | 0.226 | 0.308 | -1.58 | 0.115 | 0.172 |
| left medial orbitofrontal thickness | **-0.34** | **0.004** | **0.009** | **-0.26** | **0.027** | **0.049** | 0.64 | 0.525 | 0.618 |
| left middle temporal thickness | **-0.27** | **0.022** | **0.041** | **-0.40** | **6.11e-04** | **0.002** | -1.16 | 0.248 | 0.335 |
| left parahippocampal thickness | 0.03 | 0.776 | 0.836 | 0.23 | 0.053 | 0.087 | 1.26 | 0.207 | 0.285 |
| left paracentral thickness | 0.06 | 0.642 | 0.720 | 0.14 | 0.251 | 0.339 | 0.51 | 0.607 | 0.690 |
| left pars triangularis thickness | **0.33** | **0.005** | **0.011** | -0.03 | 0.785 | 0.843 | **-2.32** | **0.020** | **0.038** |
| left pars opercularis thickness | -0.13 | 0.298 | 0.389 | -0.10 | 0.427 | 0.522 | 0.23 | 0.821 | 0.872 |
| left pars orbitalis thickness | -0.21 | 0.081 | 0.129 | **-0.28** | **0.021** | **0.039** | -0.45 | 0.656 | 0.732 |
| left pericalcarine thickness | 0.09 | 0.447 | 0.542 | 0.03 | 0.824 | 0.873 | -0.46 | 0.646 | 0.723 |
| left postcentral thickness | 0.13 | 0.299 | 0.390 | 0.14 | 0.241 | 0.328 | 0.09 | 0.926 | 0.948 |
| left posterior cingulate thickness | -0.18 | 0.133 | 0.196 | -0.20 | 0.102 | 0.155 | -0.11 | 0.909 | 0.935 |
| left precentral thickness | 0.09 | 0.484 | 0.577 | 0.09 | 0.472 | 0.566 | 0.01 | 0.990 | 0.992 |
| left precuneus thickness | 0.07 | 0.550 | 0.642 | -0.03 | 0.825 | 0.874 | -0.79 | 0.431 | 0.525 |
| left rostral anterior cingulate thickness | **-0.30** | **0.010** | **0.021** | -0.19 | 0.118 | 0.177 | 0.88 | 0.378 | 0.477 |
| left rostral middle frontal thickness | -0.01 | 0.915 | 0.939 | **-0.29** | **0.014** | **0.027** | **-2.30** | **0.021** | **0.040** |
| left superior frontal thickness | 0.02 | 0.843 | 0.888 | -0.07 | 0.571 | 0.659 | -0.57 | 0.572 | 0.660 |
| left superior parietal thickness | 0.04 | 0.722 | 0.787 | -0.03 | 0.808 | 0.861 | -0.44 | 0.663 | 0.737 |
| left superior temporal thickness | -0.24 | 0.049 | 0.082 | -0.26 | 0.033 | 0.059 | -0.15 | 0.881 | 0.914 |

| | | | | | | | | | |
|---|---|---|---|---|---|---|---|---|---|
| left supramarginal thickness | 0.02 | 0.859 | 0.898 | **-0.36** | **0.002** | **0.005** | **-2.69** | **0.007** | **0.015** |
| left frontal pole thickness | -0.23 | 0.058 | 0.095 | -0.10 | 0.434 | 0.528 | 0.92 | 0.356 | 0.454 |
| left temporal pole thickness | -0.24 | 0.048 | 0.081 | 0.07 | 0.563 | 0.653 | 1.83 | 0.067 | 0.109 |
| left transverse temporal thickness | **-0.30** | **0.012** | **0.023** | **-0.32** | **0.008** | **0.016** | -0.12 | 0.907 | 0.933 |
| left insula thickness | -0.07 | 0.539 | 0.631 | -0.09 | 0.448 | 0.542 | -0.12 | 0.907 | 0.933 |
| right banks of superior temporal sulcus thickness | -0.13 | 0.290 | 0.381 | -0.21 | 0.087 | 0.137 | -0.52 | 0.600 | 0.684 |
| right caudal anterior cingulate thickness | -0.16 | 0.174 | 0.247 | **-0.32** | **0.007** | **0.014** | -1.25 | 0.210 | 0.289 |
| right caudal middle frontal thickness | 0.05 | 0.656 | 0.732 | -0.16 | 0.194 | 0.269 | -1.40 | 0.163 | 0.233 |
| right cuneus thickness | 0.10 | 0.428 | 0.523 | -0.03 | 0.802 | 0.856 | -0.85 | 0.394 | 0.492 |
| right entorhinal thickness | -0.23 | 0.057 | 0.094 | -0.23 | 0.053 | 0.088 | -0.02 | 0.981 | 0.986 |
| right fusiform thickness | -0.21 | 0.088 | 0.137 | -0.19 | 0.111 | 0.168 | 0.10 | 0.921 | 0.944 |
| right inferior parietal thickness | -0.21 | 0.077 | 0.123 | -0.16 | 0.188 | 0.262 | 0.58 | 0.561 | 0.651 |
| right inferior temporal thickness | -0.14 | 0.256 | 0.344 | -0.19 | 0.124 | 0.184 | -0.35 | 0.727 | 0.792 |
| right isthmus cingulate thickness | -0.02 | 0.871 | 0.907 | -0.22 | 0.062 | 0.101 | -1.36 | 0.175 | 0.247 |
| right lateral occipital thickness | 0.18 | 0.128 | 0.189 | 0.10 | 0.423 | 0.518 | -0.79 | 0.429 | 0.523 |
| right lateral orbitofrontal thickness | -0.05 | 0.710 | 0.776 | -0.12 | 0.307 | 0.398 | -0.62 | 0.534 | 0.626 |
| right lingual thickness | -0.04 | 0.756 | 0.817 | **-0.28** | **0.019** | **0.036** | -1.72 | 0.086 | 0.135 |
| right medial orbitofrontal thickness | **-0.35** | **0.003** | **0.006** | -0.26 | 0.029 | 0.052 | 0.80 | 0.425 | 0.520 |
| right middle temporal thickness | **-0.37** | **0.002** | **0.004** | **-0.33** | **0.005** | **0.011** | 0.32 | 0.746 | 0.809 |
| right parahippocampal thickness | 0.21 | 0.077 | 0.122 | 0.17 | 0.163 | 0.233 | -0.25 | 0.803 | 0.856 |
| right paracentral thickness | 0.17 | 0.171 | 0.243 | 0.14 | 0.256 | 0.344 | -0.21 | 0.836 | 0.882 |
| right pars triangularis thickness | 0.21 | 0.087 | 0.136 | -0.01 | 0.925 | 0.947 | -1.29 | 0.196 | 0.271 |
| right pars opercularis thickness | -0.05 | 0.683 | 0.754 | 0.05 | 0.658 | 0.734 | 0.84 | 0.402 | 0.501 |
| right pars orbitalis thickness | 3.2e-03 | 0.979 | 0.986 | -0.14 | 0.263 | 0.351 | -1.05 | 0.296 | 0.387 |
| right pericalcarine thickness | -0.03 | 0.808 | 0.861 | -0.09 | 0.443 | 0.537 | -0.40 | 0.692 | 0.761 |
| right postcentral thickness | 0.15 | 0.229 | 0.313 | -0.13 | 0.288 | 0.378 | -1.71 | 0.087 | 0.136 |
| right posterior cingulate thickness | -0.11 | 0.362 | 0.460 | **-0.34** | **0.004** | **0.008** | -1.98 | 0.048 | 0.081 |
| right precentral thickness | 0.16 | 0.187 | 0.261 | 5.5e-03 | 0.964 | 0.975 | -1.15 | 0.252 | 0.340 |
| right precuneus thickness | 0.07 | 0.539 | 0.631 | -0.14 | 0.251 | 0.339 | -1.71 | 0.087 | 0.136 |
| right rostral anterior cingulate thickness | **-0.37** | **0.001** | **0.003** | -0.15 | 0.225 | 0.307 | 1.71 | 0.088 | 0.137 |
| right rostral middle frontal thickness | -0.19 | 0.113 | 0.170 | -0.21 | 0.079 | 0.125 | -0.18 | 0.855 | 0.895 |
| right superior frontal thickness | 0.16 | 0.193 | 0.268 | 3.0e-03 | 0.980 | 0.986 | -1.20 | 0.231 | 0.315 |
| right superior parietal thickness | 0.12 | 0.314 | 0.405 | 0.18 | 0.134 | 0.196 | 0.48 | 0.633 | 0.712 |
| right superior temporal thickness | -0.18 | 0.144 | 0.209 | **-0.30** | **0.012** | **0.023** | -0.91 | 0.364 | 0.463 |
| right supramarginal thickness | 0.19 | 0.106 | 0.161 | **-0.32** | **0.006** | **0.013** | **-3.57** | **3.63e-04** | **9.60e-04** |

| | | | | | | | | | |
|---|---|---|---|---|---|---|---|---|---|
| right frontal pole thickness | -0.10 | 0.406 | 0.503 | -0.13 | 0.300 | 0.390 | -0.17 | 0.867 | 0.904 |
| right temporal pole thickness | 0.17 | 0.168 | 0.239 | -0.15 | 0.226 | 0.308 | -1.88 | 0.060 | 0.098 |
| right transverse temporal thickness | -0.03 | 0.814 | 0.866 | -0.26 | 0.031 | 0.056 | -1.58 | 0.115 | 0.172 |
| right insula thickness | -0.07 | 0.589 | 0.674 | -0.06 | 0.595 | 0.679 | 6.9e-03 | 0.995 | 0.996 |
| left banks of superior temporal sulcus area | **0.34** | **0.003** | **0.008** | **0.59** | **6.05e-08** | **2.90e-07** | **2.25** | **0.025** | **0.045** |
| left caudal anterior cingulate area | **0.35** | **0.003** | **0.007** | 0.22 | 0.066 | 0.107 | -1.61 | 0.108 | 0.163 |
| left caudal middle frontal area | **0.31** | **0.009** | **0.018** | **0.46** | **5.69e-05** | **1.70e-04** | 1.27 | 0.205 | 0.284 |
| left cuneus area | **0.41** | **4.66e-04** | **0.001** | **0.45** | **1.05e-04** | **3.05e-04** | 0.28 | 0.780 | 0.839 |
| left entorhinal area | -0.06 | 0.607 | 0.690 | 0.06 | 0.641 | 0.718 | 0.72 | 0.470 | 0.565 |
| left fusiform area | **0.31** | **0.009** | **0.018** | **0.39** | **8.89e-04** | **0.002** | 0.60 | 0.550 | 0.641 |
| left inferior parietal area | **0.59** | **1.04e-07** | **4.85e-07** | **0.69** | **2.58e-11** | **1.86e-10** | 1.36 | 0.175 | 0.247 |
| left inferior temporal area | **0.43** | **1.98e-04** | **5.47e-04** | **0.59** | **1.04e-07** | **4.84e-07** | 1.56 | 0.119 | 0.177 |
| left isthmus cingulate area | **0.34** | **0.004** | **0.009** | 0.18 | 0.146 | 0.212 | -1.10 | 0.270 | 0.359 |
| left lateral occipital area | **0.33** | **0.005** | **0.011** | **0.56** | **5.99e-07** | **2.47e-06** | 1.68 | 0.093 | 0.144 |
| left lateral orbitofrontal area | **0.51** | **6.58e-06** | **2.33e-05** | **0.65** | **8.07e-10** | **4.94e-09** | 1.75 | 0.080 | 0.128 |
| left lingual area | 0.24 | 0.046 | 0.078 | **0.40** | **5.38e-04** | **0.001** | 1.05 | 0.296 | 0.387 |
| left medial orbitofrontal area | **0.49** | **1.68e-05** | **5.53e-05** | **0.53** | **2.45e-06** | **9.23e-06** | 0.42 | 0.678 | 0.748 |
| left middle temporal area | **0.46** | **6.52e-05** | **1.93e-04** | **0.39** | **8.63e-04** | **0.002** | -0.63 | 0.532 | 0.623 |
| left parahippocampal area | -0.23 | 0.060 | 0.098 | 0.05 | 0.660 | 0.735 | 1.71 | 0.088 | 0.137 |
| left paracentral area | **0.34** | **0.003** | **0.008** | **0.71** | **5.36e-12** | **4.29e-11** | **3.52** | **4.26e-04** | **0.001** |
| left pars triangularis area | **0.40** | **6.91e-04** | **0.002** | **0.40** | **5.61e-04** | **0.001** | 0.06 | 0.954 | 0.967 |
| left pars opercularis area | **0.45** | **8.10e-05** | **2.37e-04** | **0.45** | **8.42e-05** | **2.46e-04** | -7.7e-03 | 0.994 | 0.996 |
| left pars orbitalis area | **0.60** | **4.50e-08** | **2.22e-07** | **0.53** | **2.36e-06** | **8.91e-06** | -0.70 | 0.484 | 0.578 |
| left pericalcarine area | **0.33** | **0.005** | **0.011** | **0.42** | **3.43e-04** | **9.12e-04** | 0.63 | 0.530 | 0.622 |
| left postcentral area | 0.21 | 0.088 | 0.137 | 0.23 | 0.053 | 0.088 | 0.19 | 0.848 | 0.892 |
| left posterior cingulate area | **0.30** | **0.011** | **0.022** | **0.38** | **0.001** | **0.003** | 0.86 | 0.390 | 0.489 |
| left precentral area | **0.36** | **0.002** | **0.006** | **0.46** | **5.30e-05** | **1.60e-04** | 0.94 | 0.349 | 0.446 |
| left precuneus area | **0.32** | **0.007** | **0.015** | **0.45** | **1.09e-04** | **3.16e-04** | 0.98 | 0.327 | 0.422 |
| left rostral anterior cingulate area | **0.44** | **1.35e-04** | **3.84e-04** | **0.41** | **4.25e-04** | **0.001** | -0.32 | 0.753 | 0.815 |
| left rostral middle frontal area | **0.61** | **2.61e-08** | **1.31e-07** | **0.65** | **7.92e-10** | **4.88e-09** | 0.63 | 0.526 | 0.618 |
| left superior frontal area | **0.53** | **1.85e-06** | **7.08e-06** | **0.64** | **1.72e-09** | **1.01e-08** | 1.34 | 0.180 | 0.252 |
| left superior parietal area | **0.37** | **0.002** | **0.004** | **0.50** | **9.86e-06** | **3.38e-05** | 1.08 | 0.279 | 0.369 |
| left superior temporal area | **0.29** | **0.015** | **0.029** | **0.49** | **2.00e-05** | **6.47e-05** | 1.65 | 0.099 | 0.152 |
| left supramarginal area | **0.52** | **3.70e-06** | **1.36e-05** | **0.50** | **1.25e-05** | **4.21e-05** | -0.26 | 0.797 | 0.851 |

| | | | | | | | | | |
|---|---|---|---|---|---|---|---|---|---|
| left frontal pole area | 0.25 | 0.038 | 0.066 | 0.12 | 0.304 | 0.394 | -0.76 | 0.448 | 0.542 |
| left temporal pole area | -0.10 | 0.428 | 0.523 | 0.10 | 0.397 | 0.495 | 1.11 | 0.267 | 0.356 |
| left transverse temporal area | 0.16 | 0.192 | 0.267 | **0.46** | **7.44e-05** | **2.19e-04** | 2.15 | 0.032 | 0.056 |
| left insula area | **0.28** | **0.018** | **0.033** | **0.27** | **0.022** | **0.041** | -0.09 | 0.929 | 0.949 |
| right banks of superior temporal sulcus area | **0.41** | **3.90e-04** | **0.001** | **0.32** | **0.006** | **0.013** | -0.79 | 0.427 | 0.522 |
| right caudal anterior cingulate area | **0.62** | **9.78e-09** | **5.24e-08** | **0.70** | **1.57e-11** | **1.19e-10** | 1.27 | 0.202 | 0.279 |
| right caudal middle frontal area | **0.44** | **1.14e-04** | **3.28e-04** | **0.51** | **6.09e-06** | **2.17e-05** | 0.64 | 0.519 | 0.613 |
| right cuneus area | 0.18 | 0.141 | 0.206 | **0.33** | **0.005** | **0.011** | 1.16 | 0.247 | 0.335 |
| right entorhinal area | -0.16 | 0.183 | 0.257 | 0.14 | 0.265 | 0.353 | 2.07 | 0.039 | 0.068 |
| right fusiform area | **0.44** | **1.17e-04** | **3.36e-04** | **0.61** | **2.00e-08** | **1.02e-07** | 1.52 | 0.129 | 0.190 |
| right inferior parietal area | **0.53** | **2.26e-06** | **8.58e-06** | **0.58** | **1.33e-07** | **6.06e-07** | 0.58 | 0.564 | 0.654 |
| right inferior temporal area | **0.55** | **8.35e-07** | **3.39e-06** | **0.60** | **3.37e-08** | **1.68e-07** | 0.63 | 0.532 | 0.623 |
| right isthmus cingulate area | 0.19 | 0.122 | 0.181 | **0.31** | **0.008** | **0.017** | 1.10 | 0.272 | 0.361 |
| right lateral occipital area | **0.54** | **1.44e-06** | **5.62e-06** | **0.69** | **2.48e-11** | **1.80e-10** | 1.68 | 0.093 | 0.144 |
| right lateral orbitofrontal area | **0.51** | **7.31e-06** | **2.56e-05** | **0.55** | **8.48e-07** | **3.44e-06** | 0.40 | 0.690 | 0.760 |
| right lingual area | 0.10 | 0.409 | 0.505 | **0.49** | **1.96e-05** | **6.34e-05** | **2.99** | **0.003** | **0.006** |
| right medial orbitofrontal area | **0.43** | **2.03e-04** | **5.58e-04** | **0.42** | **3.06e-04** | **8.22e-04** | -0.10 | 0.921 | 0.944 |
| right middle temporal area | **0.45** | **9.29e-05** | **2.69e-04** | 0.22 | 0.072 | 0.116 | **-2.26** | **0.024** | **0.044** |
| right parahippocampal area | 0.03 | 0.814 | 0.866 | 0.16 | 0.190 | 0.265 | 0.82 | 0.409 | 0.506 |
| right paracentral area | **0.36** | **0.002** | **0.005** | **0.49** | **2.08e-05** | **6.70e-05** | 1.17 | 0.244 | 0.331 |
| right pars triangularis area | **0.42** | **3.09e-04** | **8.29e-04** | **0.55** | **9.97e-07** | **4.00e-06** | 1.41 | 0.159 | 0.229 |
| right pars opercularis area | **0.48** | **2.23e-05** | **7.13e-05** | **0.39** | **9.43e-04** | **0.002** | -0.83 | 0.406 | 0.503 |
| right pars orbitalis area | **0.37** | **0.001** | **0.003** | **0.52** | **3.86e-06** | **1.42e-05** | 1.42 | 0.156 | 0.225 |
| right pericalcarine area | **0.44** | **1.53e-04** | **4.31e-04** | **0.28** | **0.018** | **0.033** | -1.45 | 0.146 | 0.212 |
| right postcentral area | **0.44** | **1.27e-04** | **3.64e-04** | **0.46** | **5.85e-05** | **1.74e-04** | 0.20 | 0.838 | 0.885 |
| right posterior cingulate area | **0.32** | **0.007** | **0.014** | **0.36** | **0.002** | **0.005** | 0.48 | 0.634 | 0.713 |
| right precentral area | **0.48** | **2.62e-05** | **8.27e-05** | **0.48** | **2.24e-05** | **7.17e-05** | 0.04 | 0.970 | 0.978 |
| right precuneus area | **0.43** | **1.77e-04** | **4.92e-04** | **0.44** | **1.33e-04** | **3.80e-04** | 0.06 | 0.951 | 0.965 |
| right rostral anterior cingulate area | **0.57** | **2.66e-07** | **1.17e-06** | **0.60** | **4.84e-08** | **2.37e-07** | 0.39 | 0.694 | 0.763 |
| right rostral middle frontal area | **0.49** | **1.97e-05** | **6.38e-05** | **0.57** | **2.56e-07** | **1.13e-06** | 0.92 | 0.356 | 0.454 |
| right superior frontal area | **0.56** | **4.54e-07** | **1.92e-06** | **0.69** | **5.19e-11** | **3.58e-10** | 1.74 | 0.083 | 0.131 |
| right superior parietal area | 0.22 | 0.062 | 0.102 | 0.25 | 0.038 | 0.067 | 0.18 | 0.855 | 0.895 |
| right superior temporal area | **0.50** | **8.61e-06** | **2.98e-05** | **0.36** | **0.002** | **0.005** | -1.20 | 0.230 | 0.314 |
| right supramarginal area | **0.28** | **0.021** | **0.039** | **0.28** | **0.018** | **0.034** | 0.06 | 0.955 | 0.967 |
| right frontal pole area | **0.26** | **0.027** | **0.049** | 0.20 | 0.102 | 0.156 | -0.52 | 0.602 | 0.686 |

| | | | | | | | | | |
|---|---|---|---|---|---|---|---|---|---|
| right temporal pole area | 7.4e-03 | 0.951 | 0.965 | 0.20 | 0.102 | 0.155 | 1.04 | 0.300 | 0.390 |
| right transverse temporal area | 0.26 | 0.032 | 0.057 | **0.48** | **2.36e-05** | **7.50e-05** | 1.75 | 0.080 | 0.127 |
| right insula area | 0.22 | 0.066 | 0.107 | 0.21 | 0.083 | 0.131 | -0.12 | 0.901 | 0.929 |
| left banks of superior temporal sulcus volume | 0.26 | 0.033 | 0.059 | **0.54** | **1.26e-06** | **4.97e-06** | **2.65** | **0.008** | **0.016** |
| left caudal anterior cingulate volume | **0.30** | **0.011** | **0.021** | 0.13 | 0.301 | 0.392 | -2.00 | 0.046 | 0.078 |
| left caudal middle frontal volume | **0.40** | **6.72e-04** | **0.002** | **0.52** | **3.15e-06** | **1.17e-05** | 1.06 | 0.289 | 0.380 |
| left cuneus volume | **0.31** | **0.010** | **0.020** | **0.45** | **8.19e-05** | **2.39e-04** | 1.05 | 0.292 | 0.382 |
| left entorhinal volume | -0.10 | 0.405 | 0.503 | 0.04 | 0.756 | 0.817 | 0.83 | 0.407 | 0.504 |
| left fusiform volume | **0.31** | **0.009** | **0.018** | **0.42** | **2.65e-04** | **7.20e-04** | 0.99 | 0.325 | 0.419 |
| left inferior parietal volume | **0.57** | **3.03e-07** | **1.31e-06** | **0.64** | **2.13e-09** | **1.21e-08** | 0.83 | 0.406 | 0.503 |
| left inferior temporal volume | **0.46** | **6.67e-05** | **1.97e-04** | **0.59** | **8.32e-08** | **3.92e-07** | 1.31 | 0.191 | 0.266 |
| left isthmus cingulate volume | **0.31** | **0.010** | **0.020** | 0.16 | 0.179 | 0.252 | -1.04 | 0.299 | 0.389 |
| left lateral occipital volume | **0.39** | **7.49e-04** | **0.002** | **0.49** | **1.88e-05** | **6.12e-05** | 0.72 | 0.471 | 0.565 |
| left lateral orbitofrontal volume | **0.48** | **2.98e-05** | **9.31e-05** | **0.56** | **4.92e-07** | **2.06e-06** | 0.80 | 0.423 | 0.518 |
| left lingual volume | 0.12 | 0.312 | 0.403 | **0.44** | **1.31e-04** | **3.72e-04** | 2.06 | 0.039 | 0.068 |
| left medial orbitofrontal volume | **0.30** | **0.012** | **0.024** | 0.21 | 0.088 | 0.138 | -0.78 | 0.434 | 0.528 |
| left middle temporal volume | **0.31** | **0.010** | **0.020** | **0.38** | **0.001** | **0.003** | 0.62 | 0.536 | 0.627 |
| left parahippocampal volume | -0.04 | 0.721 | 0.787 | 0.08 | 0.521 | 0.613 | 0.78 | 0.436 | 0.530 |
| left paracentral volume | **0.32** | **0.006** | **0.013** | **0.55** | **1.03e-06** | **4.10e-06** | 1.94 | 0.053 | 0.088 |
| left pars triangularis volume | **0.47** | **4.51e-05** | **1.37e-04** | **0.44** | **1.52e-04** | **4.30e-04** | -0.34 | 0.733 | 0.798 |
| left pars opercularis volume | **0.35** | **0.003** | **0.006** | **0.46** | **7.18e-05** | **2.12e-04** | 0.85 | 0.396 | 0.495 |
| left pars orbitalis volume | **0.54** | **1.27e-06** | **4.99e-06** | **0.59** | **7.73e-08** | **3.66e-07** | 0.58 | 0.561 | 0.651 |
| left pericalcarine volume | **0.32** | **0.007** | **0.014** | **0.40** | **7.13e-04** | **0.002** | 0.51 | 0.610 | 0.692 |
| left postcentral volume | 0.23 | 0.053 | 0.088 | **0.49** | **1.74e-05** | **5.70e-05** | 2.04 | 0.042 | 0.072 |
| left posterior cingulate volume | **0.27** | **0.023** | **0.042** | 0.24 | 0.041 | 0.071 | -0.27 | 0.787 | 0.844 |
| left precentral volume | **0.36** | **0.002** | **0.006** | **0.53** | **3.04e-06** | **1.14e-05** | 1.58 | 0.113 | 0.170 |
| left precuneus volume | **0.29** | **0.015** | **0.029** | **0.44** | **1.66e-04** | **4.63e-04** | 1.07 | 0.285 | 0.375 |
| left rostral anterior cingulate volume | **0.39** | **8.49e-04** | **0.002** | 0.26 | 0.028 | 0.050 | -1.21 | 0.224 | 0.307 |
| left rostral middle frontal volume | **0.51** | **7.43e-06** | **2.59e-05** | **0.41** | **4.46e-04** | **0.001** | -1.12 | 0.263 | 0.351 |
| left superior frontal volume | **0.38** | **0.001** | **0.003** | **0.39** | **9.43e-04** | **0.002** | 0.09 | 0.925 | 0.947 |
| left superior parietal volume | **0.47** | **4.29e-05** | **1.31e-04** | **0.65** | **1.50e-09** | **8.87e-09** | 1.87 | 0.061 | 0.100 |
| left superior temporal volume | 0.21 | 0.087 | 0.136 | **0.53** | **2.29e-06** | **8.68e-06** | **2.85** | **0.004** | **0.010** |
| left supramarginal volume | **0.42** | **2.70e-04** | **7.31e-04** | **0.58** | **1.10e-07** | **5.10e-07** | 1.88 | 0.059 | 0.098 |
| left frontal pole volume | -0.04 | 0.754 | 0.816 | -0.08 | 0.502 | 0.596 | -0.26 | 0.794 | 0.849 |

| | | | | | | | | | |
|---|---|---|---|---|---|---|---|---|---|
| left temporal pole volume | -0.08 | 0.495 | 0.589 | 0.10 | 0.397 | 0.495 | 1.04 | 0.299 | 0.389 |
| left transverse temporal volume | 0.11 | 0.380 | 0.479 | **0.35** | **0.003** | **0.006** | 1.85 | 0.064 | 0.105 |
| left insula volume | **0.42** | **2.65e-04** | **7.20e-04** | **0.55** | **7.20e-07** | **2.95e-06** | 1.31 | 0.192 | 0.266 |
| right banks of superior temporal sulcus volume | **0.43** | **1.89e-04** | **5.23e-04** | **0.43** | **2.20e-04** | **6.03e-04** | -0.04 | 0.968 | 0.977 |
| right caudal anterior cingulate volume | **0.64** | **2.45e-09** | **1.39e-08** | **0.53** | **2.17e-06** | **8.26e-06** | -1.51 | 0.131 | 0.192 |
| right caudal middle frontal volume | **0.47** | **4.19e-05** | **1.28e-04** | **0.47** | **3.35e-05** | **1.04e-04** | 0.05 | 0.962 | 0.974 |
| right cuneus volume | 0.16 | 0.179 | 0.252 | **0.28** | **0.018** | **0.035** | 0.81 | 0.417 | 0.512 |
| right entorhinal volume | -0.24 | 0.042 | 0.072 | 0.06 | 0.610 | 0.692 | 2.19 | 0.028 | 0.051 |
| right fusiform volume | **0.32** | **0.007** | **0.014** | **0.46** | **5.43e-05** | **1.63e-04** | 1.12 | 0.263 | 0.351 |
| right inferior parietal volume | **0.48** | **2.84e-05** | **8.93e-05** | **0.58** | **1.78e-07** | **7.99e-07** | 1.11 | 0.266 | 0.355 |
| right inferior temporal volume | **0.53** | **2.75e-06** | **1.03e-05** | **0.58** | **1.65e-07** | **7.44e-07** | 0.51 | 0.613 | 0.694 |
| right isthmus cingulate volume | 0.19 | 0.112 | 0.169 | 0.22 | 0.072 | 0.116 | 0.21 | 0.831 | 0.879 |
| right lateral occipital volume | **0.57** | **3.13e-07** | **1.36e-06** | **0.65** | **1.14e-09** | **6.80e-09** | 0.93 | 0.350 | 0.447 |
| right lateral orbitofrontal volume | **0.49** | **1.86e-05** | **6.06e-05** | **0.30** | **0.013** | **0.024** | -1.51 | 0.131 | 0.192 |
| right lingual volume | 0.04 | 0.753 | 0.815 | **0.39** | **8.34e-04** | **0.002** | **2.53** | **0.012** | **0.023** |
| right medial orbitofrontal volume | **0.28** | **0.020** | **0.038** | 0.25 | 0.036 | 0.063 | -0.21 | 0.830 | 0.878 |
| right middle temporal volume | **0.38** | **0.001** | **0.003** | 0.17 | 0.168 | 0.239 | **-2.23** | **0.026** | **0.047** |
| right parahippocampal volume | 0.12 | 0.309 | 0.400 | 0.14 | 0.245 | 0.332 | 0.10 | 0.918 | 0.941 |
| right paracentral volume | **0.49** | **1.46e-05** | **4.86e-05** | **0.45** | **9.06e-05** | **2.63e-04** | -0.43 | 0.667 | 0.740 |
| right pars triangularis volume | **0.44** | **1.41e-04** | **3.98e-04** | **0.60** | **4.16e-08** | **2.07e-07** | 1.94 | 0.052 | 0.087 |
| right pars opercularis volume | **0.48** | **3.10e-05** | **9.63e-05** | **0.32** | **0.007** | **0.014** | -1.39 | 0.166 | 0.237 |
| right pars orbitalis volume | **0.38** | **0.001** | **0.003** | **0.42** | **3.32e-04** | **8.88e-04** | 0.39 | 0.694 | 0.763 |
| right pericalcarine volume | **0.32** | **0.006** | **0.013** | **0.36** | **0.002** | **0.005** | 0.28 | 0.781 | 0.839 |
| right postcentral volume | **0.50** | **1.04e-05** | **3.53e-05** | **0.50** | **9.54e-06** | **3.28e-05** | 0.02 | 0.984 | 0.989 |
| right posterior cingulate volume | **0.37** | **0.002** | **0.004** | **0.33** | **0.005** | **0.011** | -0.43 | 0.664 | 0.739 |
| right precentral volume | **0.50** | **1.12e-05** | **3.80e-05** | **0.52** | **4.93e-06** | **1.78e-05** | 0.18 | 0.858 | 0.898 |
| right precuneus volume | **0.31** | **0.008** | **0.017** | **0.37** | **0.002** | **0.004** | 0.47 | 0.637 | 0.715 |
| right rostral anterior cingulate volume | **0.53** | **2.12e-06** | **8.07e-06** | **0.58** | **1.85e-07** | **8.26e-07** | 0.55 | 0.580 | 0.667 |
| right rostral middle frontal volume | **0.31** | **0.008** | **0.016** | **0.27** | **0.026** | **0.047** | -0.47 | 0.636 | 0.715 |
| right superior frontal volume | **0.47** | **3.63e-05** | **1.12e-04** | **0.40** | **5.72e-04** | **0.001** | -0.80 | 0.422 | 0.517 |
| right superior parietal volume | **0.28** | **0.020** | **0.037** | **0.34** | **0.004** | **0.009** | 0.55 | 0.583 | 0.670 |
| right superior temporal volume | **0.53** | **2.03e-06** | **7.76e-06** | **0.39** | **9.15e-04** | **0.002** | -1.47 | 0.142 | 0.207 |
| right supramarginal volume | **0.39** | **9.04e-04** | **0.002** | **0.40** | **7.07e-04** | **0.002** | 0.07 | 0.942 | 0.957 |
| right frontal pole volume | 0.03 | 0.791 | 0.847 | 0.08 | 0.500 | 0.595 | 0.35 | 0.727 | 0.792 |
| right temporal pole volume | 0.11 | 0.365 | 0.463 | 0.12 | 0.333 | 0.427 | 0.04 | 0.967 | 0.977 |

| | | | | | | | | | |
|---|---|---|---|---|---|---|---|---|---|
| right transverse temporal volume | 0.23 | 0.055 | 0.091 | **0.56** | **4.74e-07** | **2.00e-06** | **2.65** | **0.008** | **0.016** |
| right insula volume | 0.23 | 0.052 | 0.087 | **0.32** | **0.006** | **0.013** | 0.88 | 0.378 | 0.477 |
| left lateral ventricle subcortical volume | **0.96** | **9.47e-38** | **2.58e-35** | **0.96** | **3.70e-38** | **1.15e-35** | 0.23 | 0.816 | 0.867 |
| left inf lat vent subcortical volume | **0.61** | **1.48e-08** | **7.71e-08** | **0.56** | **4.09e-07** | **1.75e-06** | -0.53 | 0.594 | 0.679 |
| left cerebellum white matter subcortical volume | **0.50** | **9.03e-06** | **3.12e-05** | **0.47** | **3.52e-05** | **1.09e-04** | -0.33 | 0.738 | 0.802 |
| left cerebellum cortex subcortical volume | **0.74** | **3.75e-13** | **3.53e-12** | **0.77** | **5.39e-15** | **6.56e-14** | 0.69 | 0.491 | 0.585 |
| left thalamus proper subcortical volume | **0.41** | **5.04e-04** | **0.001** | **0.55** | **8.70e-07** | **3.52e-06** | 1.57 | 0.115 | 0.173 |
| left caudate subcortical volume | **0.49** | **1.43e-05** | **4.79e-05** | **0.44** | **1.61e-04** | **4.52e-04** | -0.67 | 0.505 | 0.598 |
| left putamen subcortical volume | **0.38** | **0.001** | **0.003** | **0.30** | **0.011** | **0.021** | -0.71 | 0.476 | 0.570 |
| left pallidum subcortical volume | **0.27** | **0.025** | **0.046** | **0.33** | **0.005** | **0.011** | 0.47 | 0.638 | 0.716 |
| 3rd ventricle subcortical volume | **0.72** | **1.65e-12** | **1.43e-11** | **0.70** | **1.44e-11** | **1.10e-10** | -0.52 | 0.600 | 0.683 |
| 4th ventricle subcortical volume | **0.62** | **7.64e-09** | **4.15e-08** | **0.74** | **1.52e-13** | **1.49e-12** | 1.98 | 0.048 | 0.081 |
| brain stem subcortical volume | **0.79** | **3.29e-16** | **4.97e-15** | **0.82** | **1.84e-18** | **4.26e-17** | 0.98 | 0.326 | 0.420 |
| left hippocampus subcortical volume | 0.25 | 0.038 | 0.067 | **0.56** | **5.50e-07** | **2.29e-06** | **2.68** | **0.007** | **0.015** |
| left amygdala subcortical volume | **0.34** | **0.004** | **0.009** | **0.28** | **0.018** | **0.033** | -0.49 | 0.621 | 0.702 |
| csf subcortical volume | **0.59** | **7.99e-08** | **3.78e-07** | **0.62** | **8.13e-09** | **4.37e-08** | 0.82 | 0.415 | 0.511 |
| left accumbens area subcortical volume | 0.01 | 0.932 | 0.950 | -0.02 | 0.856 | 0.896 | -0.26 | 0.799 | 0.853 |
| left ventral diencephalon subcortical volume | **0.49** | **1.68e-05** | **5.52e-05** | **0.66** | **3.62e-10** | **2.30e-09** | 2.14 | 0.032 | 0.058 |
| left vessel subcortical volume | 0.02 | 0.853 | 0.895 | 0.02 | 0.839 | 0.885 | 0.01 | 0.989 | 0.992 |
| left choroid plexus subcortical volume | **0.52** | **3.10e-06** | **1.16e-05** | **0.51** | **6.21e-06** | **2.21e-05** | -0.19 | 0.847 | 0.891 |
| right lateral ventricle subcortical volume | **0.92** | **3.67e-29** | **6.67e-27** | **0.95** | **1.34e-35** | **3.25e-33** | **3.35** | **7.95e-04** | **0.002** |
| right inf lat vent subcortical volume | **0.57** | **3.16e-07** | **1.37e-06** | **0.73** | **1.19e-12** | **1.05e-11** | 2.13 | 0.033 | 0.058 |
| cerebellum white matter subcortical volume | **0.67** | **1.51e-10** | **1.01e-09** | **0.57** | **2.57e-07** | **1.13e-06** | -1.54 | 0.125 | 0.185 |
| cerebellum cortex subcortical volume | **0.69** | **4.46e-11** | **3.11e-10** | **0.81** | **2.86e-17** | **5.23e-16** | **2.84** | **0.005** | **0.010** |
| right thalamus proper subcortical volume | **0.30** | **0.013** | **0.025** | **0.44** | **1.38e-04** | **3.91e-04** | 1.49 | 0.136 | 0.199 |
| right caudate subcortical volume | 0.25 | 0.040 | 0.070 | **0.40** | **5.52e-04** | **0.001** | 1.49 | 0.136 | 0.199 |
| right putamen subcortical volume | **0.29** | **0.014** | **0.027** | **0.50** | **1.22e-05** | **4.13e-05** | 1.70 | 0.089 | 0.138 |
| right pallidum subcortical volume | 0.24 | 0.043 | 0.074 | 0.14 | 0.243 | 0.330 | -0.64 | 0.520 | 0.613 |
| right hippocampus subcortical volume | 0.18 | 0.128 | 0.189 | **0.48** | **2.55e-05** | **8.06e-05** | **2.40** | **0.016** | **0.031** |
| right amygdala subcortical volume | 0.24 | 0.048 | 0.081 | **0.28** | **0.021** | **0.039** | 0.27 | 0.790 | 0.847 |
| right accumbens area subcortical volume | 0.19 | 0.122 | 0.181 | 0.12 | 0.336 | 0.430 | -0.48 | 0.633 | 0.712 |
| right ventral diencephalon subcortical volume | **0.59** | **1.05e-07** | **4.86e-07** | **0.52** | **3.93e-06** | **1.44e-05** | -0.87 | 0.383 | 0.482 |
| right vessel subcortical volume | -0.05 | 0.666 | 0.740 | **0.81** | **2.35e-17** | **4.42e-16** | **6.44** | **1.17e-10** | **7.82e-10** |
| right choroid plexus subcortical volume | **0.50** | **1.01e-05** | **3.45e-05** | **0.54** | **1.12e-06** | **4.46e-06** | 0.51 | 0.607 | 0.690 |

| | | | | | | | | | |
|---|---|---|---|---|---|---|---|---|---|
| optic chiasm subcortical volume | 0.02 | 0.891 | 0.921 | 0.06 | 0.627 | 0.707 | 0.32 | 0.752 | 0.815 |
| corpus callosum posterior subcortical volume | **0.44** | **1.58e-04** | **4.42e-04** | **0.51** | **6.46e-06** | **2.30e-05** | 1.03 | 0.301 | 0.392 |
| corpus callosum mid posterior subcortical volume | 0.17 | 0.152 | 0.219 | 0.18 | 0.128 | 0.189 | 0.15 | 0.884 | 0.916 |
| corpus callosum central subcortical volume | **0.31** | **0.009** | **0.018** | 0.19 | 0.118 | 0.177 | -1.59 | 0.112 | 0.169 |
| corpus callosum mid anterior subcortical volume | **0.34** | **0.004** | **0.010** | 0.21 | 0.087 | 0.136 | -1.65 | 0.100 | 0.153 |
| corpus callosum anterior subcortical volume | **0.47** | **4.15e-05** | **1.27e-04** | **0.75** | **1.30e-13** | **1.29e-12** | **4.04** | **5.46e-05** | **1.63e-04** |

**STable 5.** Intra-class correlations (ICCs) of regional measurements from standard axial versus standard multi-orientation 64mT scans with 3T scans. Differences between ICC strengths were tested using Steiger's Z. A positive Z-value indicates that multi-orientation regions were more strongly correlated to 3T scans than axial-only regions. Analyses that are statistically significant after correction for multiple comparisons are in bold.

| Measurement | Standard Axial 64mT ICC with 3T | | | Standard Multi-Orientation 64mT ICC with 3T | | | Steiger | | |
|---|---|---|---|---|---|---|---|---|---|
| | ***ICC*** | ***p*** | ***q*** | ***ICC*** | ***p*** | ***q*** | ***z*** | ***p*** | ***q*** |
| left banks of superior temporal sulcus thickness | -0.20 | 0.952 | 0.977 | -0.19 | 0.944 | 0.973 | 0.07 | 0.947 | 0.975 |
| left caudal anterior cingulate thickness | 0.01 | 0.462 | 0.572 | -0.07 | 0.724 | 0.809 | -0.59 | 0.554 | 0.656 |
| left caudal middle frontal thickness | 0.17 | 0.079 | 0.130 | -0.04 | 0.638 | 0.732 | -1.47 | 0.142 | 0.215 |
| left cuneus thickness | 0.09 | 0.239 | 0.336 | -0.01 | 0.546 | 0.647 | -0.71 | 0.480 | 0.587 |
| left entorhinal thickness | -5.2e-03 | 0.517 | 0.621 | -0.16 | 0.908 | 0.948 | -0.90 | 0.366 | 0.476 |
| left fusiform thickness | -0.12 | 0.847 | 0.907 | -0.16 | 0.904 | 0.946 | -0.27 | 0.788 | 0.862 |
| left inferior parietal thickness | -0.15 | 0.898 | 0.941 | -0.19 | 0.941 | 0.972 | -0.30 | 0.763 | 0.842 |
| left inferior temporal thickness | 0.07 | 0.293 | 0.396 | -0.11 | 0.812 | 0.879 | -1.31 | 0.190 | 0.275 |
| left isthmus cingulate thickness | -0.02 | 0.577 | 0.678 | -0.18 | 0.931 | 0.966 | -0.98 | 0.329 | 0.434 |
| left lateral occipital thickness | 7.3e-03 | 0.476 | 0.584 | -7.0e-03 | 0.523 | 0.626 | -0.13 | 0.900 | 0.943 |
| left lateral orbitofrontal thickness | -0.03 | 0.609 | 0.705 | 0.02 | 0.444 | 0.556 | 0.40 | 0.690 | 0.780 |
| left lingual thickness | 0.07 | 0.274 | 0.376 | -0.13 | 0.860 | 0.912 | -1.33 | 0.183 | 0.266 |
| left medial orbitofrontal thickness | -0.29 | 0.992 | 0.996 | -0.19 | 0.948 | 0.975 | 0.76 | 0.447 | 0.559 |
| left middle temporal thickness | -0.19 | 0.940 | 0.972 | -0.27 | 0.989 | 0.995 | -0.75 | 0.453 | 0.564 |
| left parahippocampal thickness | 0.01 | 0.454 | 0.565 | 0.20 | 0.051 | 0.087 | 1.12 | 0.261 | 0.361 |
| left paracentral thickness | 0.05 | 0.346 | 0.454 | 0.11 | 0.179 | 0.261 | 0.39 | 0.697 | 0.783 |
| left pars triangularis thickness | **0.29** | **0.007** | **0.016** | -0.03 | 0.598 | 0.696 | -2.01 | 0.044 | 0.078 |
| left pars opercularis thickness | -0.12 | 0.842 | 0.903 | -0.09 | 0.773 | 0.850 | 0.23 | 0.816 | 0.883 |
| left pars orbitalis thickness | -0.19 | 0.943 | 0.973 | -0.26 | 0.984 | 0.994 | -0.45 | 0.654 | 0.746 |
| left pericalcarine thickness | 0.07 | 0.274 | 0.376 | 0.02 | 0.419 | 0.531 | -0.33 | 0.738 | 0.820 |
| left postcentral thickness | 0.12 | 0.151 | 0.226 | 0.14 | 0.119 | 0.186 | 0.10 | 0.920 | 0.958 |
| left posterior cingulate thickness | -0.13 | 0.863 | 0.913 | -0.13 | 0.857 | 0.911 | 0.02 | 0.981 | 0.993 |
| left precentral thickness | 0.08 | 0.244 | 0.342 | 0.08 | 0.246 | 0.344 | -4.3e-03 | 0.997 | 0.997 |
| left precuneus thickness | 0.06 | 0.305 | 0.408 | -0.02 | 0.574 | 0.676 | -0.67 | 0.506 | 0.611 |
| left rostral anterior cingulate thickness | -0.24 | 0.978 | 0.991 | -0.14 | 0.878 | 0.922 | 0.74 | 0.461 | 0.571 |
| left rostral middle frontal thickness | -0.01 | 0.542 | 0.643 | -0.25 | 0.982 | 0.993 | -1.90 | 0.058 | 0.099 |
| left superior frontal thickness | 0.02 | 0.421 | 0.534 | -0.06 | 0.695 | 0.782 | -0.52 | 0.604 | 0.701 |
| left superior parietal thickness | 0.04 | 0.373 | 0.485 | -0.03 | 0.586 | 0.684 | -0.39 | 0.696 | 0.783 |
| left superior temporal thickness | -0.20 | 0.952 | 0.977 | -0.23 | 0.973 | 0.989 | -0.25 | 0.804 | 0.873 |
| left supramarginal thickness | 0.02 | 0.438 | 0.550 | -0.31 | 0.995 | 0.997 | **-2.27** | **0.023** | **0.043** |

| | | | | | | | | | |
|---|---|---|---|---|---|---|---|---|---|
| left frontal pole thickness | -0.22 | 0.968 | 0.986 | -0.09 | 0.784 | 0.859 | 0.87 | 0.384 | 0.495 |
| left temporal pole thickness | -0.10 | 0.799 | 0.869 | 0.07 | 0.287 | 0.390 | 0.99 | 0.324 | 0.428 |
| left transverse temporal thickness | -0.25 | 0.980 | 0.993 | -0.27 | 0.990 | 0.995 | -0.21 | 0.831 | 0.894 |
| left insula thickness | -0.06 | 0.699 | 0.785 | -0.06 | 0.692 | 0.781 | 0.02 | 0.986 | 0.995 |
| right banks of superior temporal sulcus thickness | -0.10 | 0.789 | 0.862 | -0.18 | 0.928 | 0.963 | -0.53 | 0.599 | 0.697 |
| right caudal anterior cingulate thickness | -0.13 | 0.859 | 0.912 | -0.22 | 0.966 | 0.985 | -0.68 | 0.495 | 0.601 |
| right caudal middle frontal thickness | 0.05 | 0.344 | 0.452 | -0.14 | 0.878 | 0.922 | -1.24 | 0.214 | 0.306 |
| right cuneus thickness | 0.07 | 0.277 | 0.379 | -0.02 | 0.581 | 0.680 | -0.65 | 0.518 | 0.622 |
| right entorhinal thickness | -0.10 | 0.803 | 0.872 | -0.21 | 0.962 | 0.982 | -0.66 | 0.512 | 0.617 |
| right fusiform thickness | -0.14 | 0.882 | 0.925 | -0.13 | 0.868 | 0.916 | 0.06 | 0.954 | 0.977 |
| right inferior parietal thickness | -0.17 | 0.923 | 0.960 | -0.12 | 0.850 | 0.907 | 0.50 | 0.617 | 0.711 |
| right inferior temporal thickness | -0.10 | 0.801 | 0.871 | -0.13 | 0.854 | 0.910 | -0.18 | 0.855 | 0.911 |
| right isthmus cingulate thickness | -0.01 | 0.537 | 0.639 | -0.11 | 0.824 | 0.889 | -0.66 | 0.511 | 0.615 |
| right lateral occipital thickness | 0.13 | 0.139 | 0.212 | 0.07 | 0.285 | 0.387 | -0.56 | 0.574 | 0.676 |
| right lateral orbitofrontal thickness | -0.04 | 0.637 | 0.731 | -0.10 | 0.794 | 0.866 | -0.44 | 0.663 | 0.755 |
| right lingual thickness | -0.03 | 0.610 | 0.705 | -0.23 | 0.971 | 0.988 | -1.35 | 0.178 | 0.259 |
| right medial orbitofrontal thickness | -0.31 | 0.996 | 0.997 | -0.21 | 0.961 | 0.982 | 0.87 | 0.386 | 0.496 |
| right middle temporal thickness | -0.27 | 0.988 | 0.995 | -0.27 | 0.987 | 0.995 | 0.02 | 0.981 | 0.993 |
| right parahippocampal thickness | 0.08 | 0.252 | 0.350 | 0.13 | 0.140 | 0.213 | 0.28 | 0.783 | 0.858 |
| right paracentral thickness | 0.14 | 0.125 | 0.194 | 0.13 | 0.146 | 0.220 | -0.08 | 0.933 | 0.966 |
| right pars triangularis thickness | 0.19 | 0.059 | 0.100 | -0.01 | 0.535 | 0.637 | -1.17 | 0.240 | 0.337 |
| right pars opercularis thickness | -0.04 | 0.638 | 0.732 | 0.04 | 0.359 | 0.469 | 0.70 | 0.486 | 0.592 |
| right pars orbitalis thickness | 2.8e-03 | 0.491 | 0.597 | -0.10 | 0.795 | 0.867 | -0.76 | 0.449 | 0.561 |
| right pericalcarine thickness | -0.02 | 0.572 | 0.674 | -0.08 | 0.736 | 0.818 | -0.34 | 0.735 | 0.817 |
| right postcentral thickness | 0.15 | 0.113 | 0.177 | -0.13 | 0.858 | 0.911 | -1.71 | 0.087 | 0.141 |
| right posterior cingulate thickness | -0.07 | 0.731 | 0.815 | -0.19 | 0.942 | 0.973 | -0.94 | 0.347 | 0.455 |
| right precentral thickness | 0.16 | 0.092 | 0.148 | 5.5e-03 | 0.482 | 0.589 | -1.15 | 0.252 | 0.350 |
| right precuneus thickness | 0.06 | 0.299 | 0.402 | -0.12 | 0.836 | 0.897 | -1.45 | 0.147 | 0.221 |
| right rostral anterior cingulate thickness | -0.30 | 0.994 | 0.997 | -0.10 | 0.806 | 0.874 | 1.43 | 0.154 | 0.229 |
| right rostral middle frontal thickness | -0.18 | 0.932 | 0.966 | -0.17 | 0.926 | 0.962 | 0.04 | 0.964 | 0.984 |
| right superior frontal thickness | 0.16 | 0.098 | 0.157 | 2.6e-03 | 0.491 | 0.597 | -1.16 | 0.247 | 0.345 |
| right superior parietal thickness | 0.10 | 0.200 | 0.288 | 0.15 | 0.103 | 0.164 | 0.41 | 0.685 | 0.775 |
| right superior temporal thickness | -0.17 | 0.922 | 0.959 | -0.28 | 0.991 | 0.996 | -0.80 | 0.422 | 0.535 |
| right supramarginal thickness | 0.18 | 0.062 | 0.105 | -0.29 | 0.993 | 0.996 | **-3.23** | **0.001** | **0.003** |

| | | | | | | | | | |
|---|---|---|---|---|---|---|---|---|---|
| right frontal pole thickness | -0.09 | 0.780 | 0.856 | -0.12 | 0.843 | 0.903 | -0.19 | 0.852 | 0.908 |
| right temporal pole thickness | 0.10 | 0.213 | 0.304 | -0.13 | 0.858 | 0.911 | -1.34 | 0.180 | 0.263 |
| right transverse temporal thickness | -0.02 | 0.572 | 0.674 | -0.20 | 0.953 | 0.977 | -1.22 | 0.222 | 0.315 |
| right insula thickness | -0.06 | 0.678 | 0.768 | -0.05 | 0.652 | 0.745 | 0.05 | 0.957 | 0.979 |
| left banks of superior temporal sulcus area | **0.34** | **0.002** | **0.004** | **0.59** | **3.30e-08** | **1.68e-07** | **2.24** | **0.025** | **0.047** |
| left caudal anterior cingulate area | **0.28** | **0.008** | **0.017** | 0.17 | 0.075 | 0.124 | -1.35 | 0.177 | 0.259 |
| left caudal middle frontal area | **0.29** | **0.007** | **0.015** | **0.45** | **4.33e-05** | **1.37e-04** | 1.32 | 0.188 | 0.273 |
| left cuneus area | **0.41** | **2.15e-04** | **6.02e-04** | **0.44** | **5.45e-05** | **1.69e-04** | 0.25 | 0.799 | 0.869 |
| left entorhinal area | -0.06 | 0.692 | 0.781 | 0.05 | 0.331 | 0.436 | 0.69 | 0.493 | 0.599 |
| left fusiform area | **0.31** | **0.005** | **0.010** | **0.38** | **5.97e-04** | **0.002** | 0.53 | 0.593 | 0.691 |
| left inferior parietal area | **0.57** | **1.03e-07** | **4.99e-07** | **0.68** | **3.24e-11** | **2.35e-10** | 1.34 | 0.181 | 0.263 |
| left inferior temporal area | **0.42** | **1.25e-04** | **3.64e-04** | **0.56** | **2.09e-07** | **9.67e-07** | 1.35 | 0.176 | 0.258 |
| left isthmus cingulate area | **0.33** | **0.002** | **0.005** | 0.17 | 0.078 | 0.129 | -1.08 | 0.279 | 0.381 |
| left lateral occipital area | **0.32** | **0.003** | **0.007** | **0.53** | **1.25e-06** | **5.14e-06** | 1.47 | 0.141 | 0.214 |
| left lateral orbitofrontal area | **0.51** | **2.90e-06** | **1.13e-05** | **0.61** | **7.14e-09** | **3.95e-08** | 1.17 | 0.243 | 0.340 |
| left lingual area | **0.24** | **0.023** | **0.042** | **0.40** | **2.89e-04** | **7.88e-04** | 1.02 | 0.309 | 0.412 |
| left medial orbitofrontal area | **0.48** | **1.09e-05** | **3.84e-05** | **0.52** | **1.43e-06** | **5.84e-06** | 0.43 | 0.670 | 0.760 |
| left middle temporal area | **0.45** | **4.19e-05** | **1.32e-04** | **0.39** | **4.21e-04** | **0.001** | -0.55 | 0.583 | 0.682 |
| left parahippocampal area | -0.16 | 0.906 | 0.947 | 0.05 | 0.353 | 0.462 | 1.23 | 0.218 | 0.311 |
| left paracentral area | **0.34** | **0.002** | **0.004** | **0.68** | **2.52e-11** | **1.86e-10** | **3.16** | **0.002** | **0.004** |
| left pars triangularis area | **0.40** | **3.23e-04** | **8.75e-04** | **0.39** | **3.39e-04** | **9.16e-04** | -0.01 | 0.989 | 0.995 |
| left pars opercularis area | **0.45** | **4.43e-05** | **1.39e-04** | **0.44** | **6.32e-05** | **1.93e-04** | -0.07 | 0.944 | 0.973 |
| left pars orbitalis area | **0.59** | **4.06e-08** | **2.04e-07** | **0.51** | **3.00e-06** | **1.16e-05** | -0.78 | 0.438 | 0.550 |
| left pericalcarine area | **0.33** | **0.002** | **0.006** | **0.38** | **5.62e-04** | **0.001** | 0.34 | 0.731 | 0.815 |
| left postcentral area | 0.20 | 0.044 | 0.078 | **0.23** | **0.026** | **0.047** | 0.20 | 0.839 | 0.900 |
| left posterior cingulate area | **0.30** | **0.006** | **0.013** | **0.37** | **8.03e-04** | **0.002** | 0.77 | 0.442 | 0.554 |
| left precentral area | **0.35** | **0.001** | **0.003** | **0.46** | **3.05e-05** | **9.90e-05** | 0.91 | 0.365 | 0.475 |
| left precuneus area | **0.31** | **0.004** | **0.010** | **0.42** | **1.21e-04** | **3.54e-04** | 0.86 | 0.392 | 0.503 |
| left rostral anterior cingulate area | **0.33** | **0.003** | **0.006** | **0.26** | **0.016** | **0.030** | -0.68 | 0.498 | 0.604 |
| left rostral middle frontal area | **0.56** | **2.02e-07** | **9.38e-07** | **0.63** | **2.14e-09** | **1.27e-08** | 0.88 | 0.381 | 0.492 |
| left superior frontal area | **0.52** | **1.89e-06** | **7.54e-06** | **0.63** | **2.76e-09** | **1.61e-08** | 1.29 | 0.198 | 0.285 |
| left superior parietal area | **0.37** | **7.83e-04** | **0.002** | **0.48** | **1.03e-05** | **3.67e-05** | 0.93 | 0.350 | 0.459 |
| left superior temporal area | **0.29** | **0.008** | **0.016** | **0.49** | **8.71e-06** | **3.14e-05** | 1.66 | 0.097 | 0.155 |
| left supramarginal area | **0.50** | **3.88e-06** | **1.47e-05** | **0.48** | **1.12e-05** | **3.92e-05** | -0.23 | 0.818 | 0.884 |
| left frontal pole area | **0.24** | **0.020** | **0.038** | 0.12 | 0.151 | 0.226 | -0.73 | 0.467 | 0.577 |

| | | | | | | | | | |
|---|---|---|---|---|---|---|---|---|---|
| left temporal pole area | -0.06 | 0.693 | 0.781 | 0.07 | 0.274 | 0.376 | 0.74 | 0.459 | 0.569 |
| left transverse temporal area | 0.13 | 0.145 | 0.219 | **0.44** | **7.45e-05** | **2.25e-04** | 2.17 | 0.030 | 0.054 |
| left insula area | **0.27** | **0.012** | **0.024** | **0.27** | **0.013** | **0.025** | -0.02 | 0.981 | 0.993 |
| right banks of superior temporal sulcus area | **0.39** | **3.81e-04** | **0.001** | **0.32** | **0.003** | **0.007** | -0.61 | 0.539 | 0.640 |
| right caudal anterior cingulate area | **0.49** | **8.19e-06** | **2.97e-05** | **0.53** | **1.04e-06** | **4.35e-06** | 0.57 | 0.567 | 0.671 |
| right caudal middle frontal area | **0.40** | **2.76e-04** | **7.58e-04** | **0.46** | **3.32e-05** | **1.07e-04** | 0.52 | 0.605 | 0.701 |
| right cuneus area | 0.17 | 0.073 | 0.122 | **0.33** | **0.003** | **0.006** | 1.11 | 0.266 | 0.366 |
| right entorhinal area | -0.13 | 0.863 | 0.913 | 0.12 | 0.169 | 0.250 | 1.72 | 0.086 | 0.140 |
| right fusiform area | **0.43** | **1.03e-04** | **3.01e-04** | **0.58** | **6.15e-08** | **3.02e-07** | 1.35 | 0.177 | 0.259 |
| right inferior parietal area | **0.49** | **6.16e-06** | **2.28e-05** | **0.57** | **1.42e-07** | **6.76e-07** | 0.79 | 0.427 | 0.538 |
| right inferior temporal area | **0.52** | **1.36e-06** | **5.57e-06** | **0.59** | **3.37e-08** | **1.71e-07** | 0.74 | 0.459 | 0.569 |
| right isthmus cingulate area | 0.17 | 0.072 | 0.121 | **0.31** | **0.004** | **0.009** | 1.17 | 0.244 | 0.341 |
| right lateral occipital area | **0.54** | **6.37e-07** | **2.77e-06** | **0.67** | **9.85e-11** | **6.81e-10** | 1.35 | 0.175 | 0.258 |
| right lateral orbitofrontal area | **0.51** | **3.26e-06** | **1.25e-05** | **0.48** | **1.25e-05** | **4.34e-05** | -0.26 | 0.797 | 0.868 |
| right lingual area | 0.10 | 0.209 | 0.299 | **0.49** | **9.01e-06** | **3.24e-05** | **2.96** | **0.003** | **0.007** |
| right medial orbitofrontal area | **0.43** | **1.05e-04** | **3.09e-04** | **0.41** | **2.07e-04** | **5.82e-04** | -0.16 | 0.870 | 0.917 |
| right middle temporal area | **0.45** | **4.27e-05** | **1.35e-04** | 0.22 | 0.036 | 0.064 | **-2.25** | **0.024** | **0.045** |
| right parahippocampal area | 0.02 | 0.433 | 0.545 | 0.15 | 0.112 | 0.176 | 0.79 | 0.429 | 0.541 |
| right paracentral area | **0.36** | **9.93e-04** | **0.002** | **0.47** | **1.71e-05** | **5.82e-05** | 1.03 | 0.301 | 0.403 |
| right pars triangularis area | **0.42** | **1.46e-04** | **4.19e-04** | **0.54** | **7.53e-07** | **3.22e-06** | 1.26 | 0.208 | 0.298 |
| right pars opercularis area | **0.48** | **1.10e-05** | **3.85e-05** | **0.38** | **4.63e-04** | **0.001** | -0.82 | 0.413 | 0.525 |
| right pars orbitalis area | **0.35** | **0.001** | **0.003** | **0.49** | **5.83e-06** | **2.17e-05** | 1.35 | 0.175 | 0.258 |
| right pericalcarine area | **0.44** | **6.86e-05** | **2.09e-04** | **0.28** | **0.009** | **0.019** | -1.47 | 0.141 | 0.214 |
| right postcentral area | **0.43** | **7.50e-05** | **2.26e-04** | **0.45** | **3.66e-05** | **1.17e-04** | 0.19 | 0.850 | 0.907 |
| right posterior cingulate area | **0.32** | **0.003** | **0.007** | **0.35** | **0.001** | **0.003** | 0.37 | 0.714 | 0.799 |
| right precentral area | **0.48** | **1.16e-05** | **4.03e-05** | **0.48** | **1.06e-05** | **3.75e-05** | 0.02 | 0.984 | 0.994 |
| right precuneus area | **0.43** | **8.69e-05** | **2.58e-04** | **0.43** | **9.91e-05** | **2.92e-04** | -0.03 | 0.977 | 0.991 |
| right rostral anterior cingulate area | **0.41** | **1.91e-04** | **5.40e-04** | **0.40** | **2.67e-04** | **7.38e-04** | -0.12 | 0.907 | 0.948 |
| right rostral middle frontal area | **0.40** | **2.73e-04** | **7.50e-04** | **0.51** | **3.31e-06** | **1.27e-05** | 1.09 | 0.276 | 0.378 |
| right superior frontal area | **0.51** | **3.49e-06** | **1.34e-05** | **0.62** | **5.09e-09** | **2.86e-08** | 1.42 | 0.157 | 0.233 |
| right superior parietal area | 0.21 | 0.040 | 0.071 | 0.23 | 0.027 | 0.050 | 0.15 | 0.881 | 0.925 |
| right superior temporal area | **0.48** | **1.18e-05** | **4.10e-05** | **0.35** | **0.001** | **0.003** | -1.07 | 0.286 | 0.388 |
| right supramarginal area | **0.27** | **0.011** | **0.023** | **0.28** | **0.009** | **0.019** | 0.07 | 0.948 | 0.975 |
| right frontal pole area | **0.26** | **0.014** | **0.028** | 0.20 | 0.050 | 0.086 | -0.49 | 0.625 | 0.720 |

| | | | | | | | | | |
|---|---|---|---|---|---|---|---|---|---|
| right temporal pole area | 4.7e-03 | 0.485 | 0.591 | 0.14 | 0.129 | 0.199 | 0.71 | 0.475 | 0.584 |
| right transverse temporal area | **0.25** | **0.016** | **0.031** | **0.48** | **1.03e-05** | **3.67e-05** | 1.76 | 0.079 | 0.130 |
| right insula area | 0.16 | 0.087 | 0.142 | 0.19 | 0.060 | 0.101 | 0.23 | 0.818 | 0.884 |
| left banks of superior temporal sulcus volume | **0.25** | **0.017** | **0.033** | **0.53** | **9.84e-07** | **4.15e-06** | **2.55** | **0.011** | **0.022** |
| left caudal anterior cingulate volume | **0.29** | **0.008** | **0.016** | 0.12 | 0.160 | 0.238 | -1.87 | 0.062 | 0.105 |
| left caudal middle frontal volume | **0.34** | **0.002** | **0.004** | **0.48** | **9.63e-06** | **3.43e-05** | 1.14 | 0.254 | 0.352 |
| left cuneus volume | **0.30** | **0.005** | **0.011** | **0.45** | **3.77e-05** | **1.20e-04** | 1.05 | 0.292 | 0.395 |
| left entorhinal volume | -0.10 | 0.798 | 0.868 | 0.04 | 0.379 | 0.490 | 0.82 | 0.412 | 0.524 |
| left fusiform volume | **0.31** | **0.005** | **0.010** | **0.41** | **1.90e-04** | **5.39e-04** | 0.89 | 0.375 | 0.486 |
| left inferior parietal volume | **0.53** | **1.21e-06** | **5.01e-06** | **0.59** | **2.84e-08** | **1.46e-07** | 0.68 | 0.494 | 0.600 |
| left inferior temporal volume | **0.45** | **4.57e-05** | **1.43e-04** | **0.56** | **2.36e-07** | **1.08e-06** | 1.06 | 0.288 | 0.390 |
| left isthmus cingulate volume | **0.30** | **0.006** | **0.013** | 0.16 | 0.089 | 0.145 | -0.96 | 0.337 | 0.444 |
| left lateral occipital volume | **0.39** | **4.22e-04** | **0.001** | **0.49** | **8.54e-06** | **3.09e-05** | 0.76 | 0.450 | 0.561 |
| left lateral orbitofrontal volume | **0.47** | **2.19e-05** | **7.34e-05** | **0.50** | **4.89e-06** | **1.84e-05** | 0.31 | 0.757 | 0.836 |
| left lingual volume | 0.12 | 0.155 | 0.231 | **0.44** | **5.97e-05** | **1.84e-04** | 2.06 | 0.039 | 0.070 |
| left medial orbitofrontal volume | **0.28** | **0.009** | **0.018** | 0.20 | 0.044 | 0.078 | -0.65 | 0.513 | 0.617 |
| left middle temporal volume | **0.30** | **0.006** | **0.013** | **0.36** | **0.001** | **0.003** | 0.56 | 0.578 | 0.678 |
| left parahippocampal volume | -0.03 | 0.603 | 0.700 | 0.07 | 0.286 | 0.388 | 0.64 | 0.524 | 0.627 |
| left paracentral volume | **0.31** | **0.004** | **0.009** | **0.52** | **2.09e-06** | **8.29e-06** | 1.74 | 0.082 | 0.135 |
| left pars triangularis volume | **0.46** | **3.10e-05** | **1.00e-04** | **0.37** | **6.37e-04** | **0.002** | -0.86 | 0.388 | 0.499 |
| left pars opercularis volume | **0.35** | **0.001** | **0.003** | **0.45** | **3.75e-05** | **1.20e-04** | 0.83 | 0.408 | 0.521 |
| left pars orbitalis volume | **0.53** | **7.98e-07** | **3.40e-06** | **0.53** | **1.13e-06** | **4.70e-06** | -0.07 | 0.940 | 0.972 |
| left pericalcarine volume | **0.31** | **0.004** | **0.009** | **0.39** | **3.43e-04** | **9.26e-04** | 0.54 | 0.586 | 0.684 |
| left postcentral volume | 0.22 | 0.034 | 0.061 | **0.46** | **2.29e-05** | **7.65e-05** | 1.93 | 0.054 | 0.093 |
| left posterior cingulate volume | **0.27** | **0.011** | **0.022** | **0.24** | **0.020** | **0.038** | -0.27 | 0.789 | 0.862 |
| left precentral volume | **0.35** | **0.001** | **0.003** | **0.51** | **2.25e-06** | **8.87e-06** | 1.49 | 0.135 | 0.206 |
| left precuneus volume | **0.27** | **0.011** | **0.022** | **0.39** | **4.24e-04** | **0.001** | 0.82 | 0.412 | 0.525 |
| left rostral anterior cingulate volume | **0.32** | **0.003** | **0.007** | 0.21 | 0.041 | 0.072 | -1.08 | 0.280 | 0.381 |
| left rostral middle frontal volume | **0.46** | **2.40e-05** | **8.00e-05** | **0.36** | **8.86e-04** | **0.002** | -1.08 | 0.279 | 0.381 |
| left superior frontal volume | **0.36** | **0.001** | **0.003** | **0.35** | **0.001** | **0.003** | -0.03 | 0.979 | 0.992 |
| left superior parietal volume | **0.46** | **2.27e-05** | **7.58e-05** | **0.58** | **7.06e-08** | **3.45e-07** | 1.10 | 0.270 | 0.372 |
| left superior temporal volume | 0.19 | 0.055 | 0.094 | **0.48** | **1.12e-05** | **3.91e-05** | **2.48** | **0.013** | **0.026** |
| left supramarginal volume | **0.38** | **5.15e-04** | **0.001** | **0.52** | **1.52e-06** | **6.18e-06** | 1.57 | 0.117 | 0.183 |
| left frontal pole volume | -0.04 | 0.624 | 0.718 | -0.08 | 0.744 | 0.825 | -0.25 | 0.805 | 0.874 |
| left temporal pole volume | -0.07 | 0.707 | 0.792 | 0.09 | 0.234 | 0.330 | 0.86 | 0.392 | 0.503 |

| | | | | | | | | | |
|---|---|---|---|---|---|---|---|---|---|
| left transverse temporal volume | 0.08 | 0.243 | 0.340 | **0.35** | **0.001** | **0.003** | 1.91 | 0.056 | 0.096 |
| left insula volume | **0.41** | **1.80e-04** | **5.12e-04** | **0.55** | **3.06e-07** | **1.37e-06** | 1.40 | 0.163 | 0.242 |
| right banks of superior temporal sulcus volume | **0.39** | **3.36e-04** | **9.07e-04** | **0.40** | **2.73e-04** | **7.50e-04** | 0.06 | 0.953 | 0.977 |
| right caudal anterior cingulate volume | **0.58** | **6.08e-08** | **2.99e-07** | **0.47** | **1.48e-05** | **5.06e-05** | -1.38 | 0.168 | 0.248 |
| right caudal middle frontal volume | **0.40** | **3.11e-04** | **8.47e-04** | **0.38** | **6.01e-04** | **0.002** | -0.17 | 0.866 | 0.915 |
| right cuneus volume | 0.16 | 0.093 | 0.150 | **0.28** | **0.009** | **0.018** | 0.83 | 0.408 | 0.521 |
| right entorhinal volume | -0.21 | 0.961 | 0.982 | 0.06 | 0.314 | 0.417 | 1.91 | 0.056 | 0.095 |
| right fusiform volume | **0.31** | **0.005** | **0.011** | **0.42** | **1.31e-04** | **3.80e-04** | 0.89 | 0.373 | 0.485 |
| right inferior parietal volume | **0.42** | **1.33e-04** | **3.85e-04** | **0.52** | **1.96e-06** | **7.80e-06** | 1.04 | 0.297 | 0.400 |
| right inferior temporal volume | **0.47** | **1.43e-05** | **4.92e-05** | **0.55** | **2.89e-07** | **1.30e-06** | 0.75 | 0.455 | 0.565 |
| right isthmus cingulate volume | 0.18 | 0.072 | 0.120 | 0.21 | 0.037 | 0.066 | 0.33 | 0.745 | 0.825 |
| right lateral occipital volume | **0.56** | **1.46e-07** | **6.91e-07** | **0.64** | **1.09e-09** | **6.59e-09** | 0.81 | 0.416 | 0.528 |
| right lateral orbitofrontal volume | **0.49** | **9.01e-06** | **3.24e-05** | **0.28** | **0.010** | **0.020** | -1.63 | 0.103 | 0.164 |
| right lingual volume | 0.04 | 0.381 | 0.492 | **0.39** | **4.26e-04** | **0.001** | **2.50** | **0.012** | **0.025** |
| right medial orbitofrontal volume | **0.27** | **0.010** | **0.021** | **0.25** | **0.017** | **0.033** | -0.19 | 0.849 | 0.907 |
| right middle temporal volume | **0.38** | **5.09e-04** | **0.001** | 0.16 | 0.088 | 0.143 | **-2.24** | **0.025** | **0.047** |
| right parahippocampal volume | 0.09 | 0.233 | 0.329 | 0.13 | 0.132 | 0.203 | 0.28 | 0.783 | 0.858 |
| right paracentral volume | **0.45** | **3.43e-05** | **1.10e-04** | **0.41** | **1.77e-04** | **5.06e-04** | -0.42 | 0.676 | 0.767 |
| right pars triangularis volume | **0.43** | **8.39e-05** | **2.50e-04** | **0.56** | **2.34e-07** | **1.07e-06** | 1.44 | 0.149 | 0.223 |
| right pars opercularis volume | **0.48** | **1.40e-05** | **4.80e-05** | **0.32** | **0.004** | **0.008** | -1.41 | 0.159 | 0.236 |
| right pars orbitalis volume | **0.36** | **0.001** | **0.003** | **0.40** | **2.85e-04** | **7.82e-04** | 0.37 | 0.714 | 0.799 |
| right pericalcarine volume | **0.32** | **0.003** | **0.007** | **0.36** | **0.001** | **0.003** | 0.29 | 0.771 | 0.848 |
| right postcentral volume | **0.48** | **9.43e-06** | **3.37e-05** | **0.46** | **3.01e-05** | **9.82e-05** | -0.29 | 0.773 | 0.850 |
| right posterior cingulate volume | **0.37** | **8.09e-04** | **0.002** | **0.33** | **0.003** | **0.006** | -0.45 | 0.656 | 0.748 |
| right precentral volume | **0.48** | **1.06e-05** | **3.74e-05** | **0.45** | **3.67e-05** | **1.17e-04** | -0.29 | 0.774 | 0.850 |
| right precuneus volume | **0.31** | **0.004** | **0.010** | **0.34** | **0.002** | **0.005** | 0.22 | 0.829 | 0.893 |
| right rostral anterior cingulate volume | **0.40** | **2.69e-04** | **7.42e-04** | **0.46** | **2.79e-05** | **9.17e-05** | 0.68 | 0.499 | 0.604 |
| right rostral middle frontal volume | **0.26** | **0.015** | **0.030** | 0.22 | 0.033 | 0.061 | -0.36 | 0.717 | 0.803 |
| right superior frontal volume | **0.43** | **8.13e-05** | **2.44e-04** | **0.34** | **0.002** | **0.004** | -1.01 | 0.311 | 0.414 |
| right superior parietal volume | **0.25** | **0.017** | **0.033** | **0.27** | **0.011** | **0.022** | 0.17 | 0.863 | 0.913 |
| right superior temporal volume | **0.53** | **1.04e-06** | **4.37e-06** | **0.38** | **5.61e-04** | **0.001** | -1.51 | 0.132 | 0.203 |
| right supramarginal volume | **0.34** | **0.002** | **0.004** | **0.34** | **0.002** | **0.005** | -0.07 | 0.943 | 0.973 |
| right frontal pole volume | 0.03 | 0.396 | 0.506 | 0.08 | 0.250 | 0.348 | 0.35 | 0.730 | 0.814 |
| right temporal pole volume | 0.08 | 0.242 | 0.340 | 0.10 | 0.207 | 0.296 | 0.08 | 0.939 | 0.972 |

| | | | | | | | | | |
|---|---|---|---|---|---|---|---|---|---|
| right transverse temporal volume | 0.23 | 0.027 | 0.050 | **0.56** | **2.02e-07** | **9.38e-07** | **2.64** | **0.008** | **0.017** |
| right insula volume | 0.20 | 0.049 | 0.085 | **0.31** | **0.004** | **0.009** | 1.06 | 0.288 | 0.391 |
| left lateral ventricle subcortical volume | **0.95** | **2.89e-36** | **7.86e-34** | **0.95** | **1.98e-36** | **6.16e-34** | 0.10 | 0.920 | 0.958 |
| left inf lat vent subcortical volume | **0.53** | **9.80e-07** | **4.14e-06** | **0.49** | **7.89e-06** | **2.87e-05** | -0.39 | 0.697 | 0.784 |
| left cerebellum white matter subcortical volume | **0.44** | **6.51e-05** | **1.99e-04** | **0.38** | **4.94e-04** | **0.001** | -0.59 | 0.558 | 0.660 |
| left cerebellum cortex subcortical volume | **0.73** | **2.57e-13** | **2.51e-12** | **0.76** | **1.16e-14** | **1.35e-13** | 0.51 | 0.612 | 0.707 |
| left thalamus proper subcortical volume | **0.39** | **4.05e-04** | **0.001** | **0.53** | **1.03e-06** | **4.32e-06** | 1.52 | 0.128 | 0.198 |
| left caudate subcortical volume | **0.47** | **1.52e-05** | **5.19e-05** | **0.43** | **8.53e-05** | **2.53e-04** | -0.48 | 0.632 | 0.726 |
| left putamen subcortical volume | **0.38** | **5.65e-04** | **0.001** | **0.30** | **0.005** | **0.012** | -0.71 | 0.476 | 0.584 |
| left pallidum subcortical volume | **0.26** | **0.016** | **0.030** | **0.33** | **0.003** | **0.006** | 0.52 | 0.601 | 0.698 |
| 3rd ventricle subcortical volume | **0.69** | **2.00e-11** | **1.53e-10** | **0.67** | **7.56e-11** | **5.34e-10** | -0.34 | 0.731 | 0.815 |
| 4th ventricle subcortical volume | **0.60** | **1.65e-08** | **8.77e-08** | **0.69** | **1.67e-11** | **1.30e-10** | 1.36 | 0.175 | 0.258 |
| brain stem subcortical volume | **0.78** | **2.74e-16** | **4.29e-15** | **0.82** | **6.80e-19** | **1.70e-17** | 1.12 | 0.261 | 0.361 |
| left hippocampus subcortical volume | 0.17 | 0.073 | 0.121 | **0.40** | **2.66e-04** | **7.36e-04** | 1.82 | 0.068 | 0.115 |
| left amygdala subcortical volume | **0.27** | **0.011** | **0.023** | 0.22 | 0.031 | 0.056 | -0.40 | 0.689 | 0.778 |
| csf subcortical volume | **0.24** | **0.020** | **0.038** | **0.24** | **0.023** | **0.042** | -0.12 | 0.907 | 0.948 |
| left accumbens area subcortical volume | 7.6e-03 | 0.475 | 0.584 | -0.02 | 0.572 | 0.674 | -0.21 | 0.830 | 0.893 |
| left ventral diencephalon subcortical volume | **0.48** | **1.26e-05** | **4.35e-05** | **0.66** | **1.86e-10** | **1.25e-09** | 2.21 | 0.027 | 0.050 |
| left vessel subcortical volume | 8.6e-03 | 0.472 | 0.581 | 3.7e-03 | 0.488 | 0.594 | -0.03 | 0.977 | 0.991 |
| left choroid plexus subcortical volume | **0.39** | **3.28e-04** | **8.89e-04** | **0.41** | **1.83e-04** | **5.18e-04** | 0.21 | 0.833 | 0.895 |
| right lateral ventricle subcortical volume | **0.92** | **1.53e-29** | **2.79e-27** | **0.93** | **9.24e-33** | **2.24e-30** | 1.57 | 0.116 | 0.182 |
| right inf lat vent subcortical volume | **0.45** | **4.81e-05** | **1.50e-04** | **0.59** | **2.75e-08** | **1.42e-07** | 1.66 | 0.096 | 0.154 |
| cerebellum white matter subcortical volume | **0.62** | **4.73e-09** | **2.67e-08** | **0.50** | **4.15e-06** | **1.57e-05** | -1.58 | 0.114 | 0.178 |
| cerebellum cortex subcortical volume | **0.68** | **2.26e-11** | **1.67e-10** | **0.79** | **1.74e-16** | **2.85e-15** | **2.34** | **0.019** | **0.037** |
| right thalamus proper subcortical volume | **0.29** | **0.006** | **0.014** | **0.42** | **1.31e-04** | **3.79e-04** | 1.28 | 0.202 | 0.290 |
| right caudate subcortical volume | 0.19 | 0.052 | 0.089 | **0.38** | **6.35e-04** | **0.002** | 1.63 | 0.103 | 0.164 |
| right putamen subcortical volume | **0.29** | **0.007** | **0.014** | **0.50** | **5.55e-06** | **2.07e-05** | 1.69 | 0.091 | 0.147 |
| right pallidum subcortical volume | **0.24** | **0.022** | **0.041** | 0.14 | 0.124 | 0.193 | -0.64 | 0.523 | 0.626 |
| right hippocampus subcortical volume | 0.14 | 0.117 | 0.184 | **0.42** | **1.59e-04** | **4.55e-04** | 2.14 | 0.033 | 0.059 |
| right amygdala subcortical volume | 0.19 | 0.052 | 0.089 | 0.21 | 0.038 | 0.067 | 0.12 | 0.902 | 0.944 |
| right accumbens area subcortical volume | 0.17 | 0.079 | 0.131 | 0.11 | 0.176 | 0.258 | -0.39 | 0.699 | 0.785 |
| right ventral diencephalon subcortical volume | **0.58** | **5.45e-08** | **2.70e-07** | **0.52** | **1.72e-06** | **6.94e-06** | -0.82 | 0.411 | 0.524 |
| right vessel subcortical volume | -7.9e-03 | 0.526 | 0.628 | **0.63** | **1.51e-09** | **9.11e-09** | **4.24** | **2.24e-05** | **7.50e-05** |
| right choroid plexus subcortical volume | **0.41** | **1.61e-04** | **4.63e-04** | **0.50** | **5.25e-06** | **1.97e-05** | 0.91 | 0.364 | 0.474 |
| optic chiasm subcortical volume | 0.02 | 0.446 | 0.558 | 0.05 | 0.330 | 0.435 | 0.27 | 0.787 | 0.861 |

| | | | | | | | | | |
|---|---|---|---|---|---|---|---|---|---|
| corpus callosum posterior subcortical volume | **0.34** | **0.002** | **0.004** | **0.47** | **1.88e-05** | **6.37e-05** | 1.59 | 0.112 | 0.176 |
| corpus callosum mid posterior subcortical volume | 0.13 | 0.133 | 0.205 | 0.16 | 0.093 | 0.149 | 0.34 | 0.736 | 0.818 |
| corpus callosum central subcortical volume | **0.29** | **0.007** | **0.014** | 0.19 | 0.058 | 0.099 | -1.20 | 0.230 | 0.325 |
| corpus callosum mid anterior subcortical volume | **0.33** | **0.002** | **0.005** | 0.20 | 0.045 | 0.079 | -1.55 | 0.120 | 0.188 |
| corpus callosum anterior subcortical volume | **0.43** | **9.05e-05** | **2.67e-04** | **0.74** | **4.82e-14** | **5.23e-13** | **4.22** | **2.41e-05** | **8.00e-05** |

**STable 6.** Correlations of global measurements from SynthSR-processed axial 64mT scans versus SynthSR-processed multi-orientation 64mT scans with 3T scans. Differences between correlation strengths were tested using Steiger's Z. A positive Z-value indicates that the multi-orientation scans were more strongly correlated to 3T scans than the axial-only scans. Analyses that are statistically significant after correction for multiple comparisons are in bold.

| Measurement | SynthSR−Processed Axial 64mT Correlations with 3T | | | SynthSR−Processed Multi-Orientation 64mT Correlations with 3T | | | Steiger | | |
|---|---|---|---|---|---|---|---|---|---|
| | *ICC* | *p* | *q* | *ICC* | *p* | *q* | *z* | *p* | *q* |
| Total Surface Area | **0.90** | **9.01e-26** | **5.16e-25** | **0.87** | **1.42e-22** | **6.38e-22** | **-2.16** | **0.031** | **0.042** |
| Mean Cortical Thickness | 0.14 | 0.241 | 0.286 | 0.12 | 0.317 | 0.363 | -0.19 | 0.853 | 0.867 |
| Estimated Intracranial Volume | **0.85** | **1.74e-20** | **6.10e-20** | **0.84** | **1.52e-19** | **5.05e-19** | -0.89 | 0.371 | 0.410 |
| Subcortical Gray Matter Volume | **0.88** | **4.89e-24** | **2.57e-23** | **0.87** | **3.47e-22** | **1.46e-21** | -0.84 | 0.400 | 0.435 |
| Cortical Volume | **0.86** | **1.94e-21** | **7.17e-21** | **0.86** | **5.50e-22** | **2.17e-21** | 0.28 | 0.776 | 0.802 |
| Cerebral White Matter Volume | **0.92** | **1.23e-28** | **7.74e-28** | **0.92** | **1.20e-28** | **7.74e-28** | 5.4e-03 | 0.996 | 0.996 |
| Total Brain Volume | **0.97** | **1.15e-43** | **1.81e-42** | **0.96** | **3.64e-38** | **3.82e-37** | -1.94 | 0.052 | 0.069 |

**STable 7.** Correlations of individual-level differences between low- and high-field scans with motion during low-field scans (framewise displacement). Steiger's Z-tests assessed whether correlations were significantly different between low-field acquisition approaches. Analyses that are statistically significant after correction for multiple comparisons are in bold.

| Brain measure | Standard LF Axial-3T x Motion | | | SynthSR-3T x Motion | | | Steiger's Test: SynthSR-3T x Motion vs. Standard LF Axial-3T x Motion | | | Standard LF Multiple-3T x Motion | | | Steigers Test: Standard LF Multiple-3T x Motion vs. Standard LF Axial-3T x Motion | | | SynthSR Multiple-3T x Motion | | | Steiger's test: SynthSR multiple-3T x Motion vs. Standard LF Axial-3T x Motion | | |
|---|---|---|---|---|---|---|---|---|---|---|---|---|---|---|---|---|---|---|---|---|---|
| | ***r*** | ***p*** | ***q*** | ***r*** | ***p*** | ***q*** | ***Z*** | ***p*** | ***q*** | ***r*** | ***p*** | ***q*** | ***Z*** | ***p*** | ***q*** | ***r*** | ***p*** | ***q*** | ***Z*** | ***p*** | ***q*** |
| Total Surface Area | 0.03 | 0.804 | 0.975 | 0.05 | 0.697 | 0.899 | 0.12 | 0.906 | 0.979 | 1.7e-03 | 0.989 | 0.989 | -0.28 | 0.778 | 0.975 | 0.05 | 0.696 | 0.899 | 0.11 | 0.909 | 0.979 |
| Mean Cortical Thickness | -0.12 | 0.325 | 0.568 | -0.14 | 0.242 | 0.475 | -0.12 | 0.901 | 0.979 | -0.13 | 0.297 | 0.560 | -0.06 | 0.953 | 0.989 | -3.1e-03 | 0.980 | 0.989 | 0.64 | 0.520 | 0.750 |
| Estimated Intracranial Volume | 0.19 | 0.113 | 0.284 | -4.1e-03 | 0.973 | 0.989 | -1.05 | 0.293 | 0.560 | 0.20 | 0.096 | 0.268 | 0.17 | 0.862 | 0.979 | 0.03 | 0.824 | 0.975 | -0.80 | 0.423 | 0.691 |
| Subcortical Gray Matter Volume | 0.26 | 0.029 | 0.125 | 0.28 | 0.020 | 0.087 | 0.14 | 0.887 | 0.979 | 0.20 | 0.102 | 0.274 | -0.72 | 0.473 | 0.705 | 0.25 | 0.036 | 0.140 | -0.07 | 0.947 | 0.989 |
| Cortical Volume | 0.03 | 0.775 | 0.975 | -0.18 | 0.127 | 0.296 | **-2.73** | **0.006** | **0.041** | -0.08 | 0.486 | 0.710 | -1.61 | 0.107 | 0.277 | -0.12 | 0.305 | 0.563 | -1.87 | 0.061 | 0.213 |
| Cerebral White Matter Volume | 0.02 | 0.860 | 0.979 | -0.08 | 0.532 | 0.755 | -0.84 | 0.400 | 0.665 | -0.11 | 0.351 | 0.603 | -2.12 | 0.034 | 0.140 | 0.02 | 0.891 | 0.979 | -0.03 | 0.972 | 0.989 |
| Total Brain Volume | 0.20 | 0.103 | 0.274 | -0.07 | 0.564 | 0.767 | -1.83 | 0.067 | 0.219 | 0.09 | 0.435 | 0.699 | -1.27 | 0.206 | 0.428 | -0.09 | 0.475 | 0.705 | -1.84 | 0.065 | 0.219 |

**Stable 8.** Correlations of individual-level differences between low- and high-field scans with participant age. Steiger's Z-tests assessed whether correlations were significantly different between low-field acquisition approaches.

| Brain measure | Standard LF Axial-3T x Age | | | SynthSR-3T x Age | | | Steiger's Test: SynthSR-3T x Age vs. Standard LF Axial-3T x Age | | | Standard LF Multiple-3T x Age | | | Steigers Test: Standard LF Multiple-3T x Age vs. Standard LF Axial-3T x Age | | | SynthSR Multiple-3T x Age | | | Steiger's test: SynthSR multiple-3T x Age vs. Standard LF Axial-3T x Age | | |
|---|---|---|---|---|---|---|---|---|---|---|---|---|---|---|---|---|---|---|---|---|---|
| | ***r*** | ***p*** | ***q*** | ***r*** | ***p*** | ***q*** | ***Z*** | ***p*** | ***q*** | ***r*** | ***p*** | ***q*** | ***Z*** | ***p*** | ***q*** | ***r*** | ***p*** | ***q*** | ***Z*** | ***p*** | ***q*** |
| Total Surface Area | **-0.43** | **1.74e-04** | **0.002** | -0.09 | 0.473 | 0.705 | 2.56 | 0.011 | 0.061 | -0.29 | 0.015 | 0.080 | 1.55 | 0.121 | 0.290 | -0.09 | 0.470 | 0.705 | 2.42 | 0.016 | 0.080 |
| Mean Cortical Thickness | -0.09 | 0.465 | 0.705 | **-0.59** | **8.36e-08** | **2.73e-06** | **-3.17** | **0.002** | **0.014** | **-0.40** | **5.06e-04** | **0.006** | **-2.77** | **0.006** | **0.040** | **-0.38** | **0.001** | **0.013** | -1.67 | 0.095 | 0.268 |
| Estimated Intracranial Volume | -0.12 | 0.316 | 0.566 | 0.19 | 0.116 | 0.284 | 1.67 | 0.094 | 0.268 | -0.05 | 0.678 | 0.898 | 1.26 | 0.209 | 0.428 | 0.28 | 0.017 | 0.082 | 2.00 | 0.045 | 0.171 |
| Subcortical Gray Matter Volume | 0.07 | 0.543 | 0.759 | -4.2e-03 | 0.972 | 0.989 | -0.60 | 0.550 | 0.759 | -0.05 | 0.666 | 0.895 | -1.38 | 0.167 | 0.365 | -0.11 | 0.382 | 0.645 | -1.26 | 0.207 | 0.428 |
| Cortical Volume | **-0.64** | **2.18e-09** | **2.14e-07** | **-0.58** | **1.44e-07** | **3.53e-06** | 1.00 | 0.318 | 0.566 | **-0.63** | **6.57e-09** | **3.22e-07** | 0.27 | 0.788 | 0.975 | **-0.53** | **2.34e-06** | **4.59e-05** | 1.67 | 0.095 | 0.268 |
| Cerebral White Matter Volume | -0.31 | 0.010 | 0.061 | -0.28 | 0.019 | 0.087 | 0.23 | 0.818 | 0.975 | **-0.39** | **8.45e-04** | **0.009** | -1.41 | 0.158 | 0.352 | **-0.33** | **0.005** | **0.039** | -0.21 | 0.831 | 0.975 |
| Total Brain Volume | -0.03 | 0.835 | 0.975 | -0.24 | 0.047 | 0.171 | -1.47 | 0.141 | 0.321 | **-0.33** | **0.006** | **0.040** | **-3.81** | **1.36e-04** | **0.002** | -0.21 | 0.085 | 0.268 | -1.19 | 0.234 | 0.468 |

*Supplemental Figures*

**SFigure 1.** Flow chart of participant inclusion and exclusion

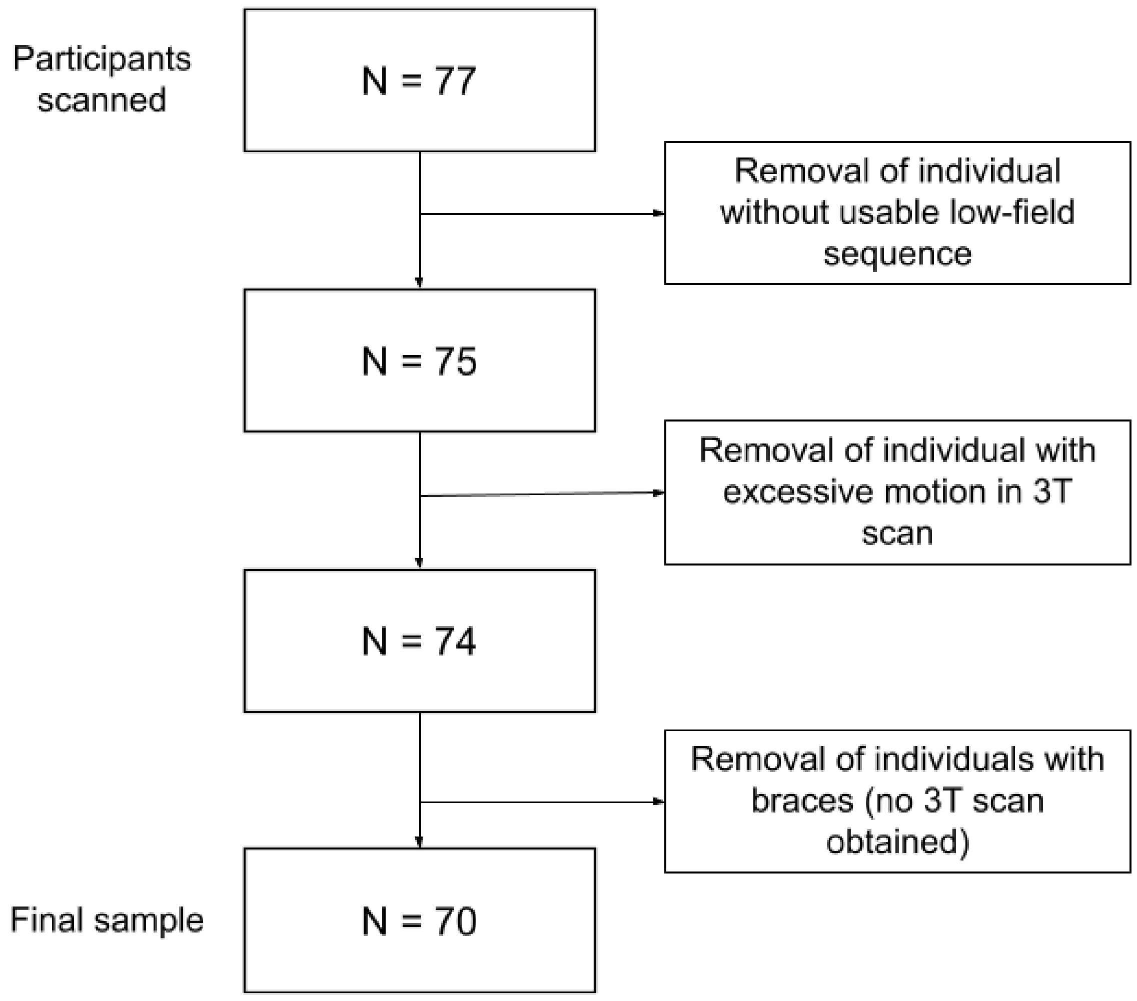

**SFigure 2.** Bland-Altman plots of agreement between 3T and standard 64mT scans. Bland-Altman plots illustrate the agreement between 3T and standard 64mT scans. Bias indicated by blue dotted line, and 95% limits of agreement by the upper and lower red dotted lines. 95% confidence intervals indicated in shaded regions. Correlation between pairwise differences and means shown by line of best fit.

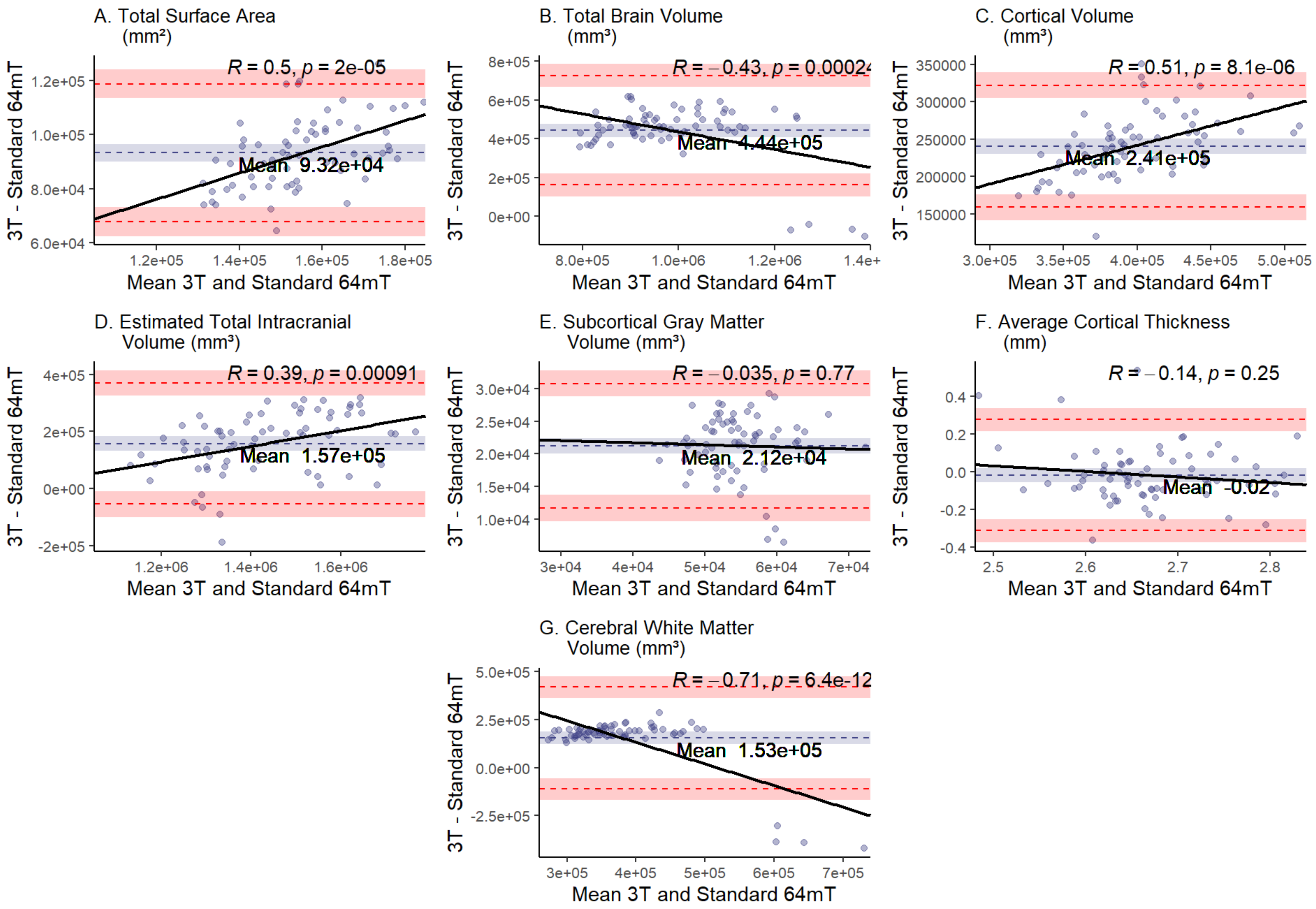

**SFigure 3.** Bland-Altman plots illustrate the agreement between 3T and SynthSR-processed 64mT scans. Bias indicated by blue dotted line, and 95% limits of agreement by the upper and lower red dotted lines. 95% confidence intervals indicated in shaded regions. Correlation between pairwise differences and means shown by line of best fit.

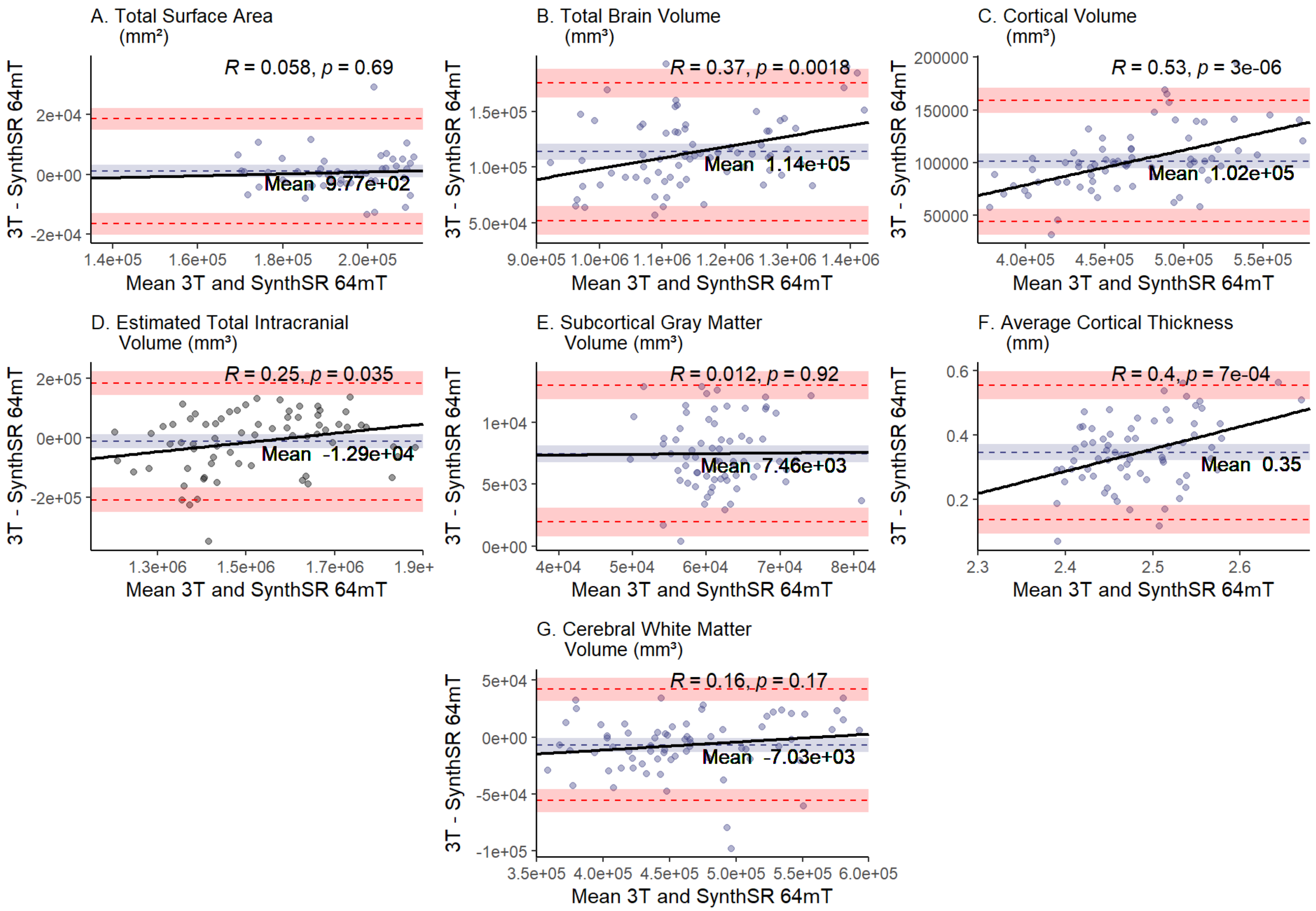

**SFigure 4.** Comparison of individual-level global measurements across standard 64mT axial scans, standard 64mT multi-orientation scans, and traditional 3T scans

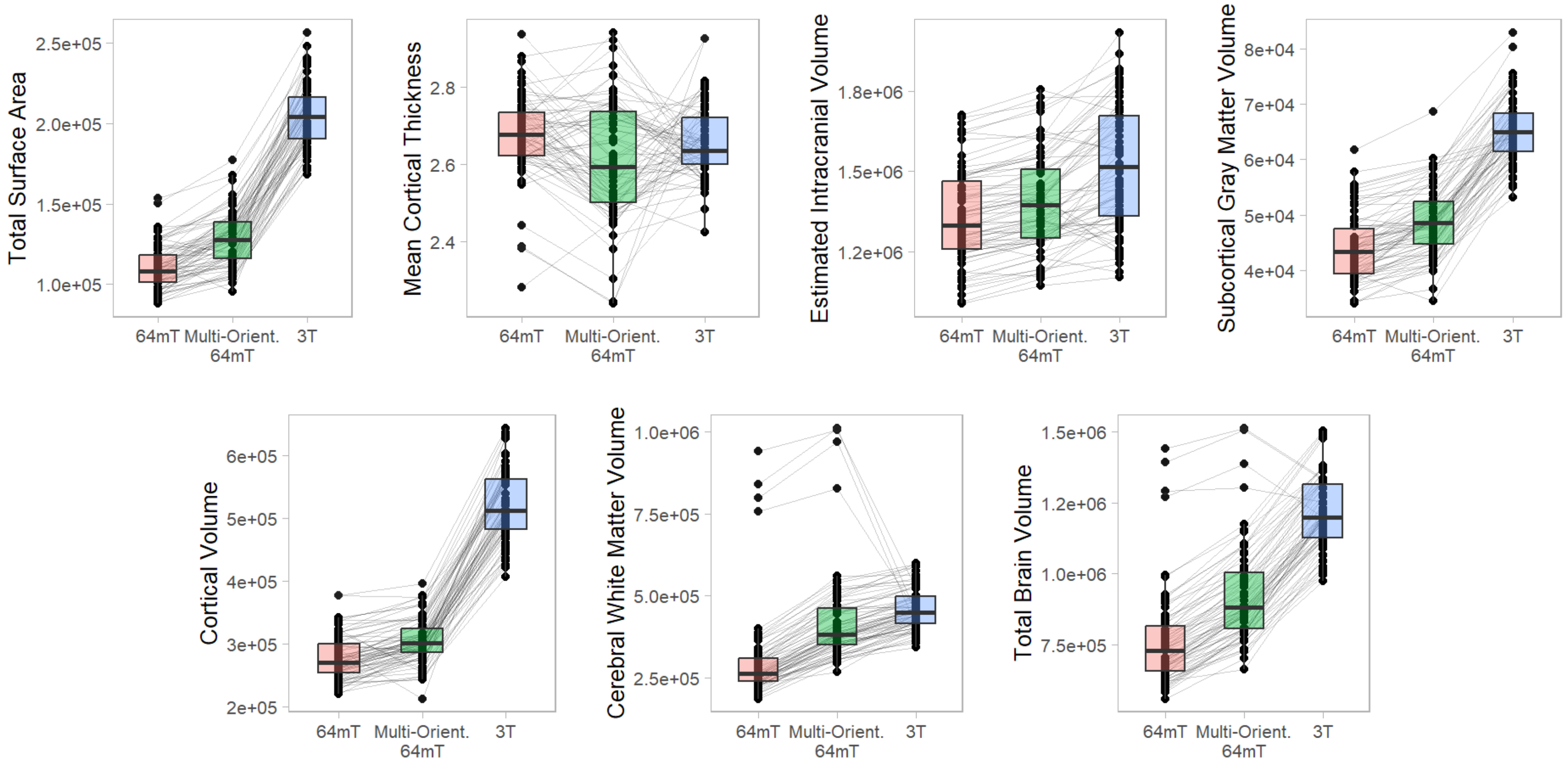

**SFigure 5.** Comparison of individual-level global measurements across standard 64mT axial scans, SynthSR-processed 64mT multi-orientation scans, and traditional 3T scans

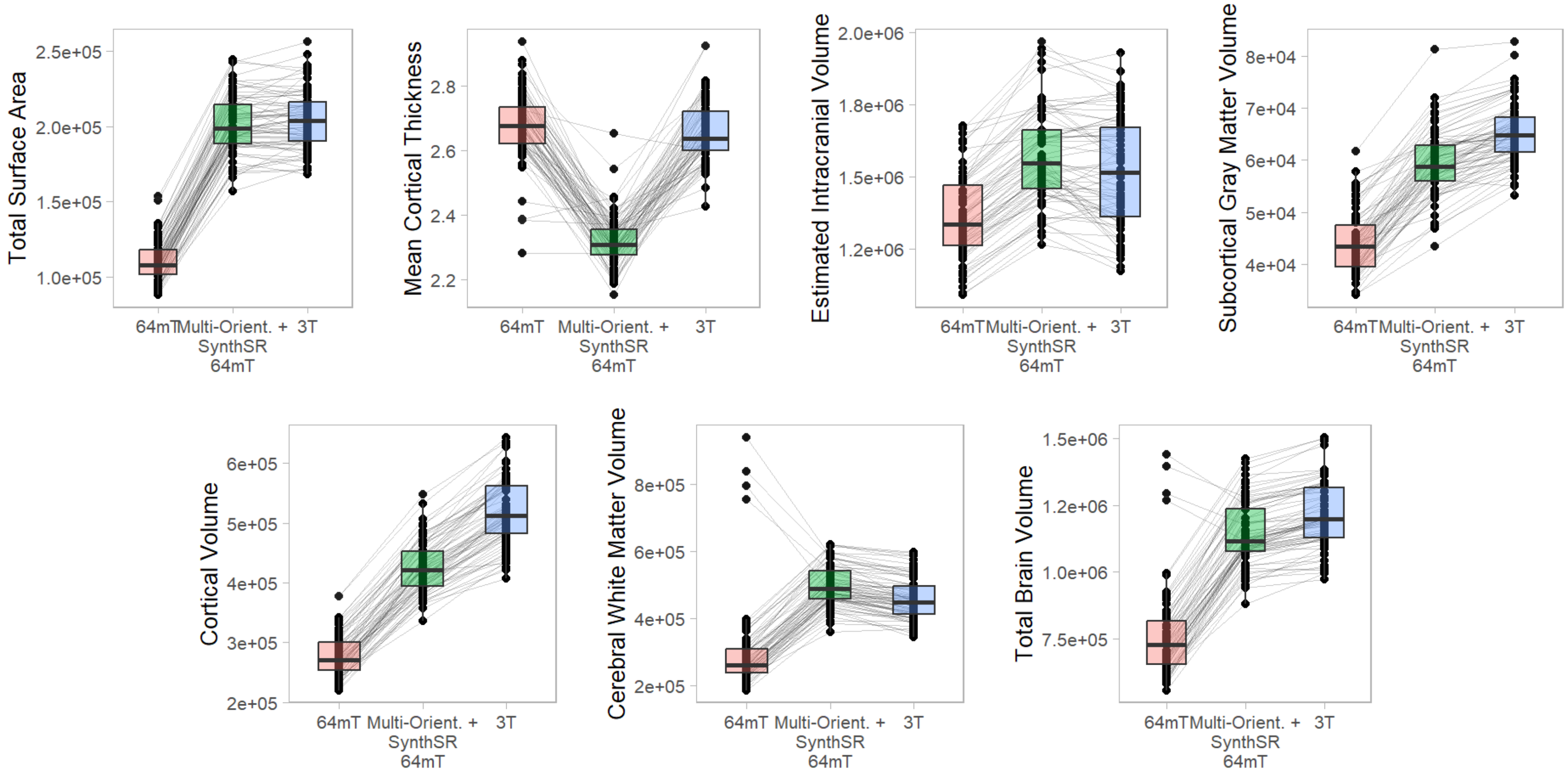

**SFigure 6.** Comparison of super-resolution approaches in improving correspondence between low-field and high-field-acquired MR images. A. Correlation of standard, single-orientation low-field images with high-field images for surface area, cortical volume, cortical thickness and subcortical volume. B. Correlation of SynthSR-processed, multi-orientation low-field images with high-field images. C. Steiger z-test values for change in correspondence between single-orientation, SynthSR-processed images and multi-orientation, SynthSR-processed images.

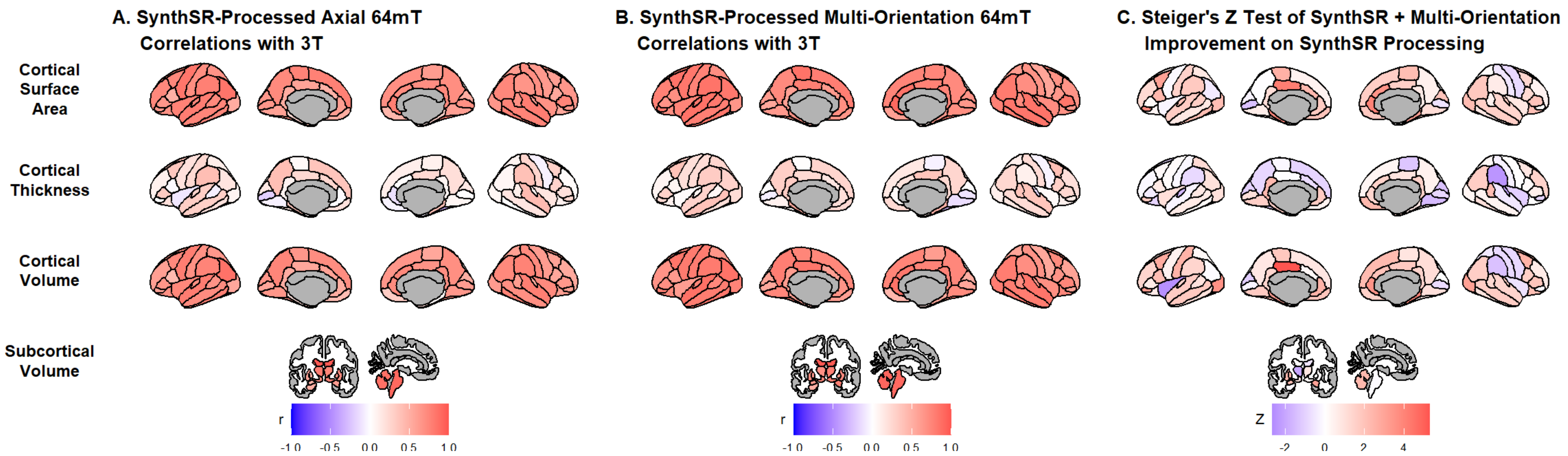
